# Screening for prostate cancer using PSA with and without MRI: systematic reviews with meta-analysis

**DOI:** 10.64898/2026.03.30.26349764

**Authors:** Jennifer Pillay, Lindsay Gaudet, Sholeh Rahman, Roland Grad, Guylène Thériault, Philipp Dahm, Keith J Todd, Gail Macartney, Brett Thombs, Sabrina Saba, Lisa Hartling

**Affiliations:** Alberta Research Centre for Health Evidence, University of Alberta; McGill University; Université de Montréal; University of Minnesota; University of Prince Edward Island

## Abstract

**Background:** Previous recommendations on screening for prostate cancer relied on ongoing trials of screening with prostate-specific antigen (PSA), which may have lacked sufficient follow-up duration to fully examine effects on mortality and overdiagnosis. Findings which consider absolute effects by age and screening intensity, along with newer guidance for assessing evidence certainty, may lead to different interpretations. Adding magnetic resonance imaging (MRI) to PSA-based screening has been raised as a way to reduce false positives (FPs) and overdiagnosis.

**Methods:** We systematically searched MEDLINE, Embase, and Central from 2014 to January 28, 2026, for randomized controlled trials (RCTs) and prospective observational studies of: (i) screening versus no screening and (ii) sequential screening with MRI for those with a positive PSA test versus PSA alone among men not known to be at high risk for prostate cancer. Studies on screening with PSA or digital rectal examination (DRE) published pre-2014 were identified from existing systematic reviews and reference lists. Studies on FPs and complications from biopsies after PSA screening did not require a control group. Paired reviewers screened titles/abstracts (assisted with artificial intelligence) and full texts, assessed risk of bias, and extracted data, by age when available. We pooled data when suitable using random-effects models, investigated heterogeneity, and assessed the certainty of evidence using GRADE with conclusions of effects based on decision thresholds based on absolute effect sizes.

**Results:** Across both questions, we included 15 RCTs (N=856,000; 8 sites of ERSPC considered separate trials) and 8 observational studies (N=56,122). At 20 years, among 1000 men who underwent repeated PSA-based screening every 2-4 years starting from age 55-69 (mean 62), there is likely a reduction in prostate-cancer mortality (≥2 fewer) and metastatic cancer incidence (≥6 fewer), at the expense of prostate-cancer overdiagnosis (≥24 cases) and FPs (≥150 cases) (all moderate certainty). If screening starts at age 50-54 or age 55, the benefits are probably smaller (e.g., 1 vs. 2 fewer prostate-cancer related deaths) with similar harms. Adding DRE or screening with PSA annually does not add benefit. One round of PSA screening or starting screening later at age 70-74 may not offer any important benefit or harm (low to moderate certainty), and any benefit from screening primarily with DRE was not shown. Compared with PSA alone, sequential screening with PSA followed by MRI reduces FPs (≥33 fewer) and overdiagnosis (via ≥10 fewer diagnoses of clinically insignificant [e.g., Gleason 6] cancers without impacting detection of clinically significant cancers) (moderate to high certainty), though findings were limited to one round of screening without long-term follow-up or measurement of mortality.

**Interpretation:** This review provides clinicians and other interest holders with anticipated absolute effects by age, and assessments of certainty across critical and important outcomes and with approximately two decades of follow-up. Findings apply to a general population and may differ for specific groups. Results for most critical outcomes, both benefits and harms, exceeded thresholds for clinically important effect sizes, thereby demonstrating the complexity of guideline developers’ and patients’ decision-making regarding screening trade-offs. Findings about adding MRI for those with a positive PSA test were limited and would require additional consideration of costs, infrastructure, expertise, and equity.

**Protocol registration:** PROSPERO - CRD420250651056.

## Background

One in 9 (12%) men in Canada develop prostate cancer during their lifetime, and 1 in 34 (2.9%) die from it.^1^ Many prostate cancers grow slowly and/or cause no symptoms, thus 10-year survival is relatively high (88%) compared with some other cancers such as colorectal (61%) and lung (15%). Increasing age is a major risk factor, with only 2% and 17% of cases diagnosed among men under age 50 and 60, respectively, and only 1.8% and 14% of cancer deaths occurring in those under age 60 and 70.^1^

Screening for asymptomatic prostate cancer aims to reduce mortality by diagnosing cancers at an earlier, curable stage. Any benefit comes with the cost of some harm,^2^ including physical (e.g., harms from biopsies and/or treatment) and psychological effects from false positives (FPs; positive screening test[s] among those without cancer) and overdiagnosis (diagnosis of a cancer that would never cause harm, never progress, or progress too slowly to ever cause symptoms^3^). Evidence on screening effectiveness has relied on randomized controlled trials (RCTs) using serum prostate-specific antigen (PSA), which at typical thresholds of 3-4 ng/mL does not have high specificity for cancer nor does it adequately discriminate cancers with differing prognoses. Sequential screening, such as PSA with triage of those with positive results sent for magnetic resonance imaging (MRI), may improve the benefits-to-harms profile by reducing FPs and diagnoses of latent low-risk cancers (i.e., overdiagnosis).

The 2014 evidence review and recommendations by the Canadian Task Force on Preventive Health Care (CTFPHC) relied on ongoing PSA screening trials which may have had insufficient follow-up to fully evaluate the effects on mortality and overdiagnosis in the context of slow-growing prostate cancer.^4, 5^ Further, the 2014 analysis did not focus on differential effects by age and screening intensity or use newer guidance to assess the certainty of evidence using absolute effects and thresholds for decision making.^6–8^

## Methods

We followed a protocol,^9^ adhered to methodological^10^ and reporting standards,^11^ and used Grading of Recommendations Assessment, Development and Evaluation (GRADE) guidance for assessing the certainty of evidence. A working group consisting of the clinician authors (RG, GT, PD, KT, GM) and BT informed the development of key questions (KQs) and eligibility criteria, rated potential outcomes for their importance, came to consensus on decision thresholds about which to draw conclusions, and provided input for the analytic approach (all prior to data extraction and analysis). They did not participate in study selection; extraction, risk of bias assessments or analysis of data; or assessment of the certainty of evidence.

### KQs and Eligibility criteria

We examined the benefits and harms of screening versus no screening (KQ1) and sequential screening with MRI after a positive PSA test versus PSA alone (KQ2) among men not known to be at high risk for prostate cancer (e.g., with germline mutations) (**Figure 1**). For KQ1, any systematically offered screening intervention was eligible (e.g., PSA of 3 or 4 ng/mL [+/− digital rectal examination (DRE)], DRE or MRI alone), and comparators could include opportunistic screening initiated during a routine healthcare visit. For KQ2 (MRI following a positive PSA test), we expected a positive screening test with MRI to be a Prostate Imaging Reporting and Data System (PI-RADs, used to grade the likelihood of clinically significant cancer)^12^, score of ≥3 or 4 (/5) though other scoring methods were allowed. For the outcomes of all-cause mortality, prostate-cancer mortality, incidence of metastatic cancer (all considered benefit outcomes), overdiagnosis (excess cancer incidence; KQ1 only), screen-detection of clinically significant and insignificant cancer (KQ2 only), harms from cancer treatment (i.e., incontinence and erectile dysfunction), health-related quality of life (HRQoL), and psychological effects from screening we sought RCTs as well as prospective cohort studies with concurrent controls and adequate control for potential confounders. Because of limited ascertainment for incidence of metastatic cancer (i.e., distant metastases at baseline through follow-up, including cases that progress after diagnosis) we also analyzed the indirect/surrogate outcome of detection of metastatic cancer, and, if necessary, advanced cancer (via screening and clinical presentation). For KQ1, uncontrolled studies were eligible for FPs (cumulative risk over ≥2 rounds of screening) and complications from biopsies after PSA screening because they are attributable to screening. For KQ2, we accepted within-subject comparisons (i.e., everyone receiving both screening strategies) for outcomes (i.e., cancer detection and FPs at first round) not confounded by downstream management of cancer. There were no exclusions based on country, and full texts had to be written in English or French. **Appendix 1** contains the full eligibility criteria.

**Figure 1:**
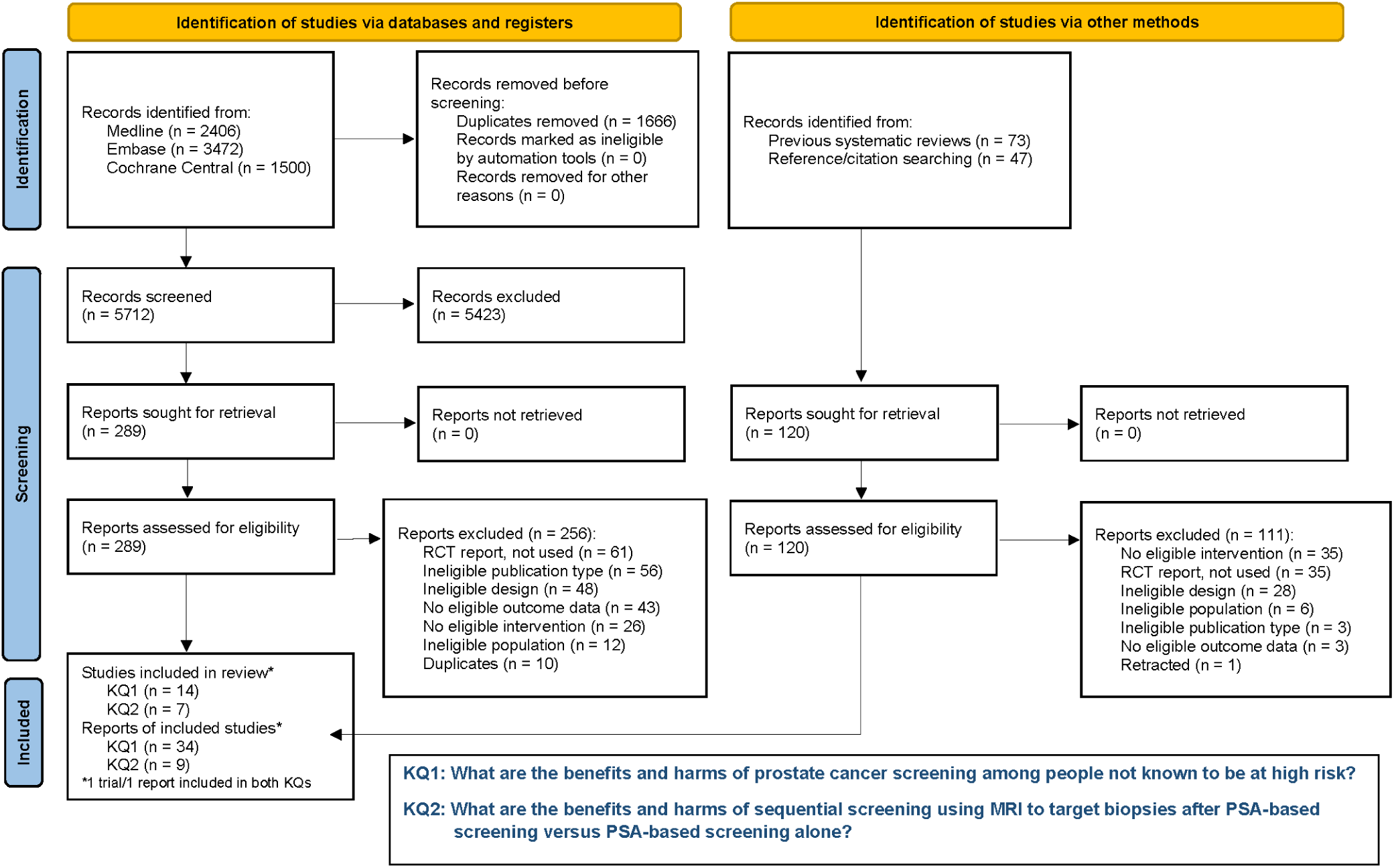
Flow of literature for both key questions (KQs) from screening and selection using the database and registry searches, then adding unique reports from other sources. Notes: RCT: randomized controlled trial.

### Data sources and searches

We located RCTs and (for harms) nonrandomized studies for KQ1 on screening with PSA or DRE published before 2014 from the previous CTFPHC review^4^ and other existing systematic reviews.^13–15^ An information specialist developed and ran de novo searches in MEDLINE, Embase and Central to capture studies relevant to both KQs published between 2014 and January 28, 2026 (**Appendix 1**). The MEDLINE search was peer reviewed.^16^ Based on existing reviews,^13, 17^ all studies relevant to KQ2 and alternative KQ1 interventions (e.g., risk calculation/stratification before PSA, MRI alone) would be captured in our searches. We hand-searched ClincialTrials.gov, reference lists of included studies, and recent systematic reviews located in our searches. Results were imported into EndNote (Clarivate Analytics, Philadelphia, USA) and duplicates removed manually.

### Study selection

Screening and selection for database records were conducted using DistillerSR (Evidence Partners Inc., Ottawa, Canada). After training DistillerAI with 200 records confirmed by reviewers as included/excluded reports, titles and abstracts were screened independently by two reviewers until the program estimated we had identified ≥95% of included papers (and with ≥50% of records screened) at which point we changed to single-reviewer screening.^18^ Selection of full texts used two independent reviewers with consensus or, if necessary, adjudication by a third reviewer. Records not found in our database searches were screened in Excel by one reviewer with another verifying exclusions. Because there are many reports about some of the trials, final inclusion of individual reports was determined after charting data.

### Data charting and extraction

Before data extraction, we charted aspects of each report such as follow-up timepoint, outcome definitions, age-specific data availability, and subgroup analysis by screening intensity, race, ethnicity or family history. There was particular interest in examining screening effects by age and screening intensity (number and frequency of rounds). Due to variations in age eligibility, screening tests, PSA thresholds, and screening intensities across the eight European screening trial (ERSPC) sites, we treated each trial site as a separate RCT when possible. For all studies, we prioritized reports with data by starting age (50-54, 55-69 and 70-74 years) and then longest follow-up. We contacted authors for results by age or for outcomes of interest (e.g., by site) when these were not reported. Our main interest was an intention-to-screen approach though we extracted data for per protocol (i.e., attenders of screening) analyses where reported. **Appendix 1** contains additional details on extraction.

An extraction form was created in Excel and pilot-tested to ensure accuracy and completeness. One reviewer extracted data, another verified all data for accuracy and completeness, and consensus resolved discrepancies.

### Risk of bias

Risk of bias was assessed in duplicate for each finding of interest, and final decisions used consensus. For KQ1 RCTs we applied ROB 2.^19^ Concerns about deviations from interventions/contamination (e.g., <50% contrast in screening between arms, little-to no increase in cancer detection during screening phase) were only rated at high risk if we thought that they would affect the outcome; because of confidence that this bias would drive findings towards the null, and because we were primarily concerned about whether the effect surpassed a threshold, the risk was rated as high if the study results fell below the relevant threshold. For uncontrolled data on harms, we adapted the NIH tool for Observational Cohort Studies primarily by removing questions on comparisons (**Appendix 1**).^20^ For KQ2, we used QUADAS-C^21^ for comparative accuracy; because the reference standard intentionally differed by group (e.g., systematic 10-12 core biopsies for PSA screening versus MRI-targeted biopsies [+/− systematic biopsies] for sequential screening), we did not consider this a potential bias as our focus was on detection rates and FPs, not on true accuracy. Overall risk was designated as either high (i.e., one tool domain at high risk or, for harms, based on an algorithm [**Appendix 1**]), some concerns, or low, with the latter two combined as “lower risk” for subgroup analysis.

### Analysis

When more than one study reported on an outcome, we pooled their data when suitable using random effects models (**Appendix 1** contains additional details on our meta-analysis). The decision to pool did not rely on statistical heterogeneity (I^2^ statistic) but rather on clinical and methodological differences.^22^

We also assessed for potential effect moderation. For KQ1, variables of interest about the population were age, race, ethnicity, and family history of prostate cancer. For these we relied on individual participant/with-in study data reported or provided by authors. Intervention- and study-related variables, for which we applied subgroup/stratified analyses, were screening intensity (i.e., low [∼ 1 round] vs. moderate [2-4 rounds offered to a majority and ≥3-year interval] vs. high [≥5 rounds and annual/biannual screening), screening test (i.e., PSA vs. PSA or DRE), and risk of bias (high vs. lower). For KQ2, subgroup analyses of interest were based on the screening test threshold(s) (e.g., PI-RADs 3 vs. 4) and biopsy approach (e.g., 10-12 core vs. 6-core for systematic biopsies in the PSA alone group, and MRI-targeted vs. MRI-targeted plus systematic biopsies in the MRI + PSA group). We judged subgroup effects for their credibility based on several criteria.^6, 23^

### Certainty of evidence and summary of findings

We assessed the certainty of evidence using GRADE. All outcomes started at high certainty, including harms from uncontrolled studies because of their attribution to screening.^24^ Two reviewers independently assessed the certainty of evidence, with disagreements resolved by consensus or arbitration by a third reviewer.

Before analysis and any availability of results, the working group used a survey and consensus to develop decision thresholds for each outcome^8, 25^ that reflected an absolute magnitude of effect for which a majority (>50%) of patients would likely think of as small but important. Relative effects were converted into absolute effects by multiplying an assumed control event rate (CER) by the relative effect point estimate (and its 95% confidence interval [CI]) from the analysis.^26^ CERs were created using data by age from Canadian statistics (for cancer incidence and all-cause mortality) and other included studies (e.g., trials with lowest contamination rates) (**Appendix 1**). Based on study follow-up timepoints and life expectancy, CERs and absolute effects for KQ1 capture a time horizon of 20 years for starting screening between age 50 and 69 and of 15 years for starting at age 70-74. Because some effects differed substantially from the thresholds, *post hoc* we report magnitudes of effect larger than the thresholds (or smaller for harms) when there was moderate/high-certainty evidence.

We created narrative statements about the findings.^26, 27^ For evidence of low certainty we use “may/possibly” and for moderate we use “probably/likely” as modifiers. Magnitude of effects in the narrative statements reflect our judgment about the best estimate of effect, considering that relying on other magnitudes, such as the point estimate from the analysis, would result in lower certainty due to imprecision and possibly other factors.

### Ethics approval

We did not need to seek ethics approval for this systematic review and meta-analysis since it relied entirely on published data.

## Results

For KQ1 we included, for benefits 13 RCTs (N=780,736; 8 from ERSPC) with results data from 31 reports^28–58^ and, for harms only, 1 RCT only (n=12,629^34^) and 3 uncontrolled cohort studies (N=5593). ^28, 59, 60^ No controlled prospective cohorts were identified that reported on benefits. For KQ2 we included 2 reports of 2 RCTs (N=87,893; one also included for harms in KQ1)^34, 61^ and 5 within-subject comparisons (N=50,529) reported in 7 reports ^62–68^ (**Figure 1**). **Appendices 2 and 3** contain additional study characteristics tables, risk of bias assessments, forest plots and tables including subgroup and per protocol analyses, and summary of findings for uncontrolled data on harms from PSA screening. **Appendix 2** also describes findings on cancer incidence (no benefit outcomes reported) from one RCT (n=9026) in KQ1 evaluating DRE-based screening.^53^

### Benefits and Harms from PSA-Based Screening versus No Screening (KQ1)

Twelve RCTs examined the effects of PSA screening (+/− DRE) versus no screening on mortality outcomes, incidence of metastatic cancer, and overdiagnosis (**Table 1**). A high risk-of-bias trial from Quebec (enrolling men age 45-80 years) was not included in the analysis due to inability to obtain age-specific data.^38^ Only one RCT reported on HRQoL^29^ and none reported treatment harms or psychological effects. Ten RCTs and three observational studies reported cumulative FPs and complications from biopsies due to PSA screening.

**Table 1.**
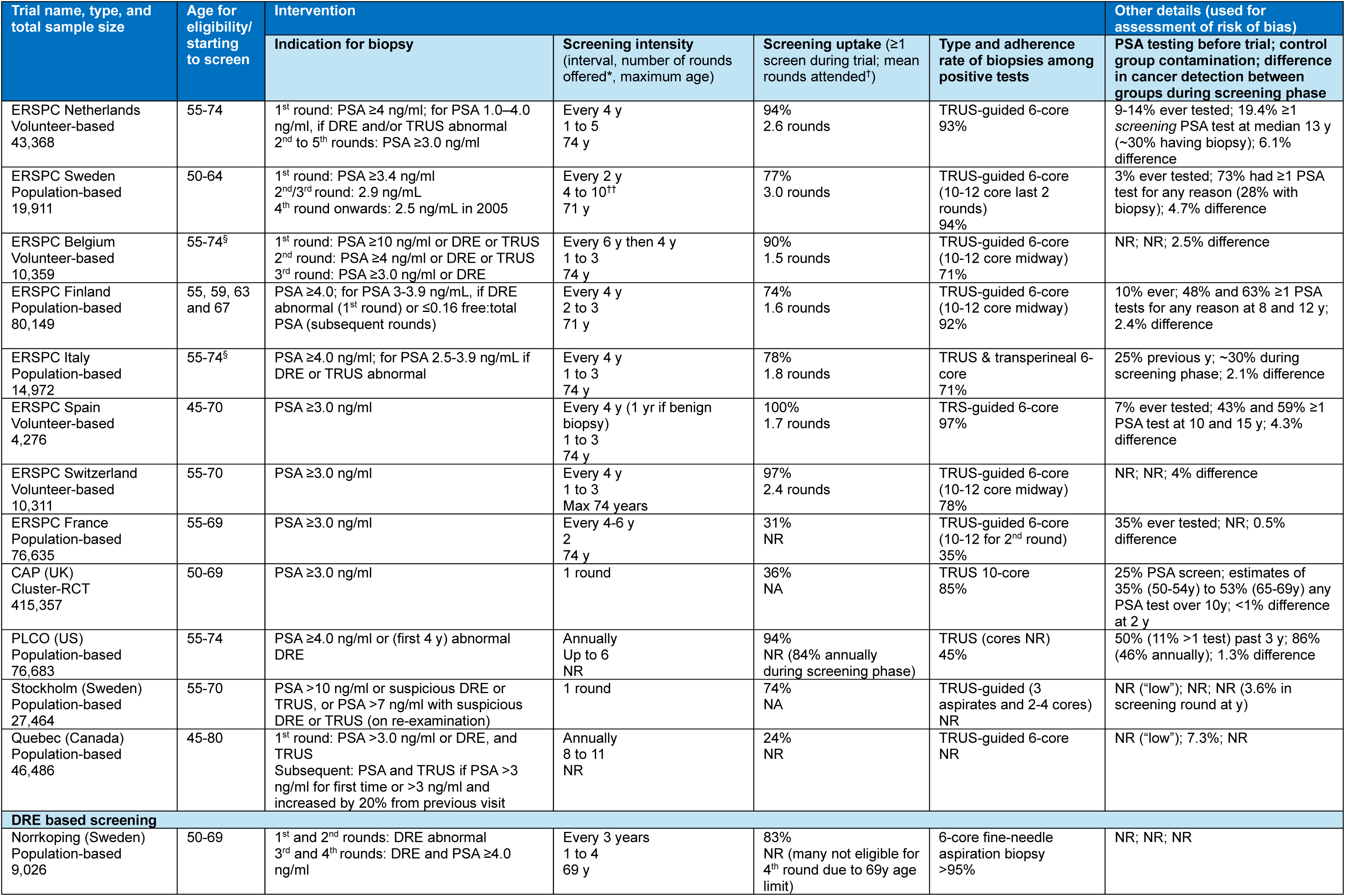
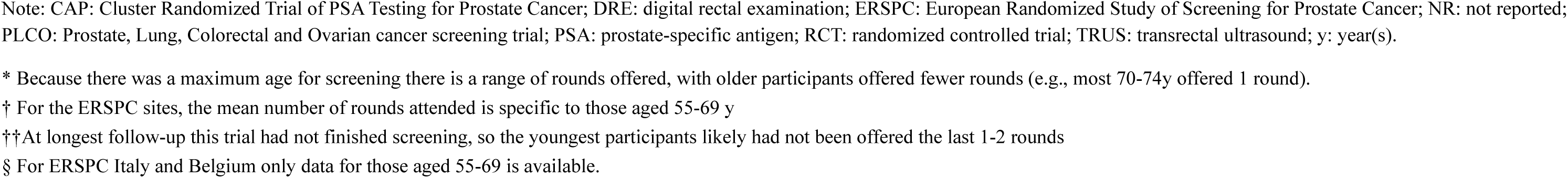
Characteristics of screening trials (KQ1)

| Trial name, type, and total sample size | Age for eligibility/ starting to screen | Intervention |  |  |  | Other details (used for assessment of risk of bias) |
| --- | --- | --- | --- | --- | --- | --- |
|  |  | Indication for biopsy | Screening intensity (interval, number of rounds offered*, maximum age) | Screening uptake (≥1 screen during trial; mean rounds attended†) | Type and adherence rate of biopsies among positive tests | PSA testing before trial; control group contamination; difference in cancer detection between groups during screening phase |
| ERSPC Netherlands Volunteer-based<br>43,368 | 55-74 | 1<sup>st</sup> round: PSA ≥4 ng/ml; for PSA 1.0–4.0 ng/ml, if DRE and/or TRUS abnormal<br>2<sup>nd</sup> to 5<sup>th</sup> rounds: PSA ≥3.0 ng/ml | Every 4 y<br>1 to 5<br>74 y | 94%<br>2.6 rounds | TRUS-guided 6-core<br>93% | 9-14% ever tested; 19.4% ≥1 screening PSA test at median 13 y (~30% having biopsy); 6.1% difference |
| ERSPC Sweden Population-based<br>19,911 | 50-64 | 1<sup>st</sup> round: PSA ≥3.4 ng/ml<br>2<sup>nd</sup>/3<sup>rd</sup> round: 2.9 ng/mL<br>4<sup>th</sup> round onwards: 2.5 ng/mL in 2005 | Every 2 y<br>4 to 10<sup>††</sup><br>71 y | 77%<br>3.0 rounds | TRUS-guided 6-core (10-12 core last 2 rounds)<br>94% | 3% ever tested; 73% had ≥1 PSA test for any reason (28% with biopsy); 4.7% difference |
| ERSPC Belgium Volunteer-based<br>10,359 | 55-74<sup>§</sup> | 1<sup>st</sup> round: PSA ≥10 ng/ml or DRE or TRUS<br>2<sup>nd</sup> round: PSA ≥4 ng/ml or DRE or TRUS<br>3<sup>rd</sup> round: PSA ≥3.0 ng/ml or DRE | Every 6 y then 4 y<br>1 to 3<br>74 y | 90%<br>1.5 rounds | TRUS-guided 6-core (10-12 core midway)<br>71% | NR; NR; 2.5% difference |
| ERSPC Finland Population-based<br>80,149 | 55, 59, 63 and 67 | PSA ≥4.0; for PSA 3-3.9 ng/mL, if DRE abnormal (1<sup>st</sup> round) or ≤0.16 free:total PSA (subsequent rounds) | Every 4 y<br>2 to 3<br>71 y | 74%<br>1.6 rounds | TRUS-guided 6-core (10-12 core midway)<br>92% | 10% ever; 48% and 63% ≥1 PSA tests for any reason at 8 and 12 y; 2.4% difference |
| ERSPC Italy Population-based<br>14,972 | 55-74<sup>§</sup> | PSA ≥4.0 ng/ml; for PSA 2.5-3.9 ng/mL if DRE or TRUS abnormal | Every 4 y<br>1 to 3<br>74 y | 78%<br>1.8 rounds | TRUS & transperineal 6-core<br>71% | 25% previous y; ~30% during screening phase; 2.1% difference |
| ERSPC Spain Volunteer-based<br>4,276 | 45-70 | PSA ≥3.0 ng/ml | Every 4 y (1 yr if benign biopsy)<br>1 to 3<br>74 y | 100%<br>1.7 rounds | TRS-guided 6-core<br>97% | 7% ever tested; 43% and 59% ≥1 PSA test at 10 and 15 y; 4.3% difference |
| ERSPC Switzerland Volunteer-based<br>10,311 | 55-70 | PSA ≥3.0 ng/ml | Every 4 y<br>1 to 3<br>Max 74 years | 97%<br>2.4 rounds | TRUS-guided 6-core (10-12 core midway)<br>78% | NR; NR; 4% difference |
| ERSPC France Population-based<br>76,635 | 55-69 | PSA ≥3.0 ng/ml | Every 4-6 y<br>2<br>74 y | 31%<br>NR | TRUS-guided 6-core (10-12 for 2<sup>nd</sup> round)<br>35% | 35% ever tested; NR; 0.5% difference |
| CAP (UK) Cluster-RCT<br>415,357 | 50-69 | PSA ≥3.0 ng/ml | 1 round | 36%<br>NA | TRUS 10-core<br>85% | 25% PSA screen; estimates of 35% (50-54y) to 53% (65-69y) any PSA test over 10y; <1% difference at 2 y |
| PLCO (US) Population-based<br>76,683 | 55-74 | PSA ≥4.0 ng/ml or (first 4 y) abnormal DRE | Annually<br>Up to 6<br>NR | 94%<br>NR (84% annually during screening phase) | TRUS (cores NR)<br>45% | 50% (11% >1 test) past 3 y; 86% (46% annually); 1.3% difference |
| Stockholm (Sweden) Population-based<br>27,464 | 55-70 | PSA >10 ng/ml or suspicious DRE or TRUS, or PSA >7 ng/ml with suspicious DRE or TRUS (on re-examination) | 1 round | 74%<br>NA | TRUS-guided (3 aspirates and 2-4 cores)<br>NR | NR ("low"); NR; NR (3.6% in screening round at y) |
| Quebec (Canada) Population-based<br>46,486 | 45-80 | 1<sup>st</sup> round: PSA >3.0 ng/ml or DRE, and TRUS<br>Subsequent: PSA and TRUS if PSA >3 ng/ml for first time or >3 ng/ml and increased by 20% from previous visit | Annually<br>8 to 11<br>NR | 24%<br>NR | TRUS-guided 6-core<br>NR | NR ("low"); 7.3%; NR |
| **DRE based screening** |  |  |  |  |  |  |
| Norrköping (Sweden) Population-based<br>9,026 | 50-69 | 1<sup>st</sup> and 2<sup>nd</sup> rounds: DRE abnormal<br>3<sup>rd</sup> and 4<sup>th</sup> rounds: DRE and PSA ≥4.0 ng/ml | Every 3 years<br>1 to 4<br>69 y | 83%<br>NR (many not eligible for 4<sup>th</sup> round due to 69y age limit) | 6-core fine-needle aspiration biopsy<br>>95% | NR; NR; NR |

Note: CAP: Cluster Randomized Trial of PSA Testing for Prostate Cancer; DRE: digital rectal examination; ERSPC: European Randomized Study of Screening for Prostate Cancer; NR: not reported; PLCO: Prostate, Lung, Colorectal and Ovarian cancer screening trial; PSA: prostate-specific antigen; RCT: randomized controlled trial; TRUS: transrectal ultrasound; y: year(s).
\* Because there was a maximum age for screening there is a range of rounds offered, with older participants offered fewer rounds (e.g., most 70-74y offered 1 round).
† For the ERSPC sites, the mean number of rounds attended is specific to those aged 55-69 y
†† At longest follow-up this trial had not finished screening, so the youngest participants likely had not been offered the last 1-2 rounds
§ For ERSPC Italy and Belgium only data for those aged 55-69 is available.

The largest trial of low-intensity screening (CAP) was at high risk of bias across outcomes, as were the mortality outcomes and overdiagnosis in several trials examining multiple screening rounds (**Figure 2**). Multiple sources apart from the main trial reports were used for these assessments (e.g., protocols, reports of contamination and treatment by arm).^69–80^ Apart from concerns about deviations from interventions, there was some concern for CAP and serious concern for ERSPC Spain about lack of allocation concealment. Data by age at enrollment were received from study authors for CAP (50-54 and 55-69 years [adjusted for clustering]), PLCO (55-69 and 70-74 years), and ERSPC Spain (45-50 and 55-70 years). For the ERSPC RCTs, data by age and site were available except for ages 70-74 (missing data on all outcomes from Belgium and Italy) and for ages 55-69 by site for all-cause mortality (where we used multi-site data).

**Figure 2:**
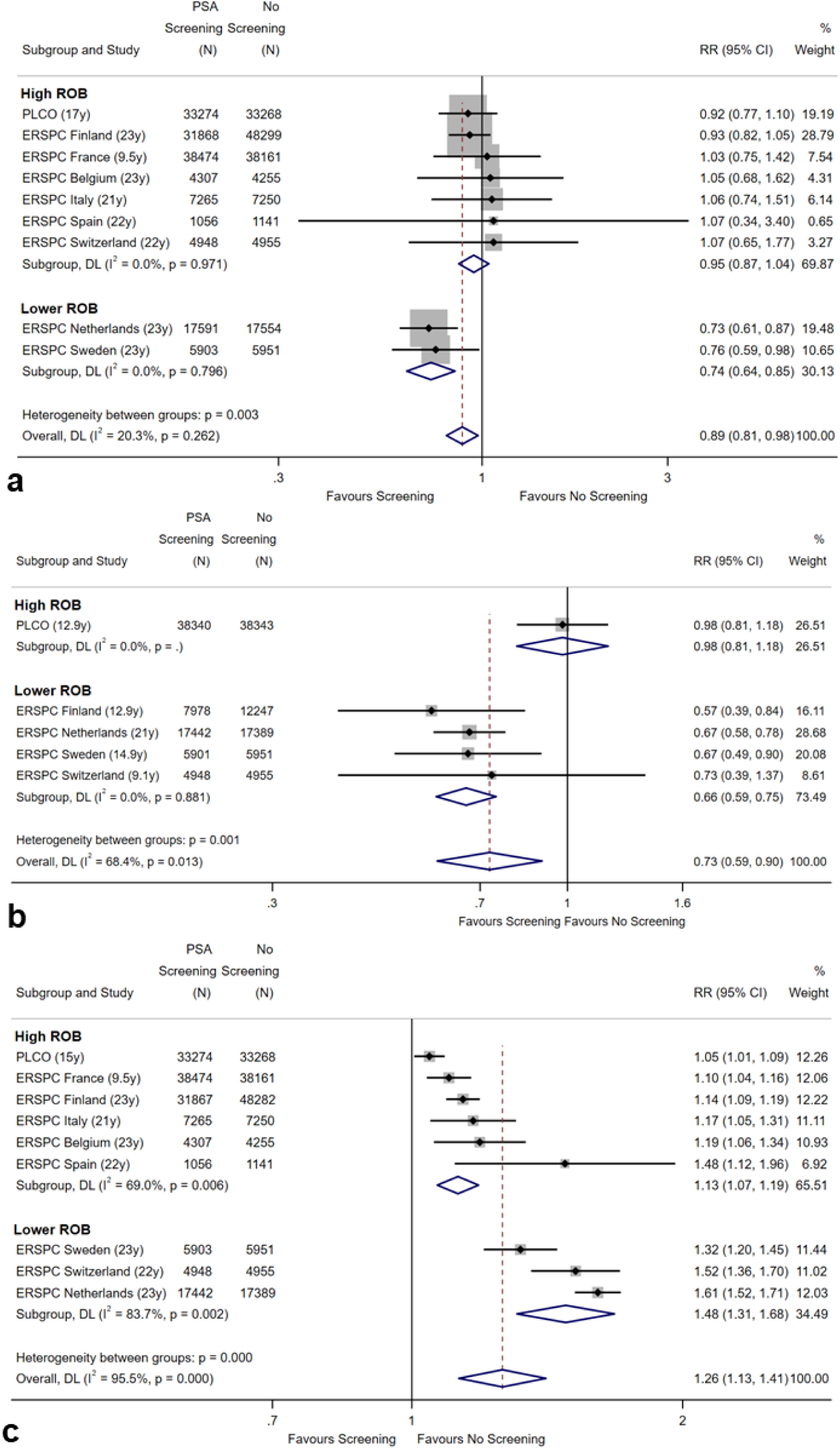
Relative effects from multiple rounds of PSA-based screening versus no screening starting from age 55-69 across all studies (RRs at bottom right of each figure) and for the credible subgroups of lower risk of bias trials on (a) prostate-cancer mortality, (b) incidence of metastatic cancer through follow-up, and (c) overdiagnosis. Absolute effects at 20 years, per 1000 men screened, for the lower risk of bias trials are in Table 2, and from analysis across all trials for those starting to screen at age 55-69 are, by outcome: (a) 1.65 (95% CI 0.3 to 2.85) fewer, (b) 6.75 (2.5 to 10.25) fewer, and (c) 20.2 (10 to 31.57) cases, and for starting screening at age 55 are (a) 0.88 (0.16 to 1.52) fewer, (b) 3.51 (1.3 to 5.33) fewer, and (c) 16.1 (8.1 to 25.4) cases. Effects across all studies for starting to screen at age 55-69 (but not at age 55) surpassed the thresholds of (a) 1 fewer, (5) 5 fewer, and (c) 20 cases, but are imprecise due to serious inconsistency largely from risk of bias. Notes: ERSPC: European Randomized Study of Screening for Prostate Cancer; N: number of participants; PLCO: Prostate, Lung, Colorectal and Ovarian; ROB: risk of bias; RR: rate ratio

**Table 2.**
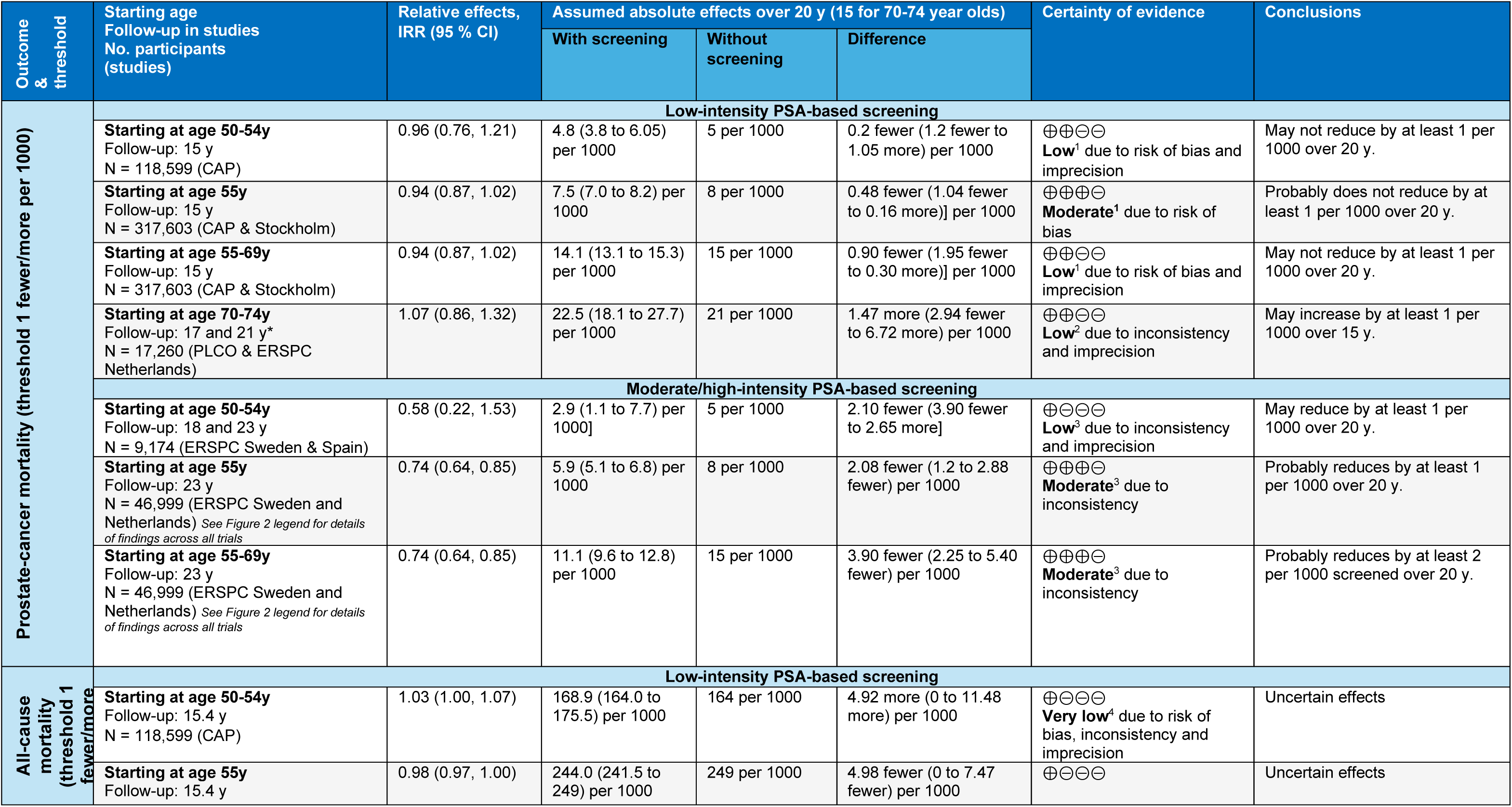
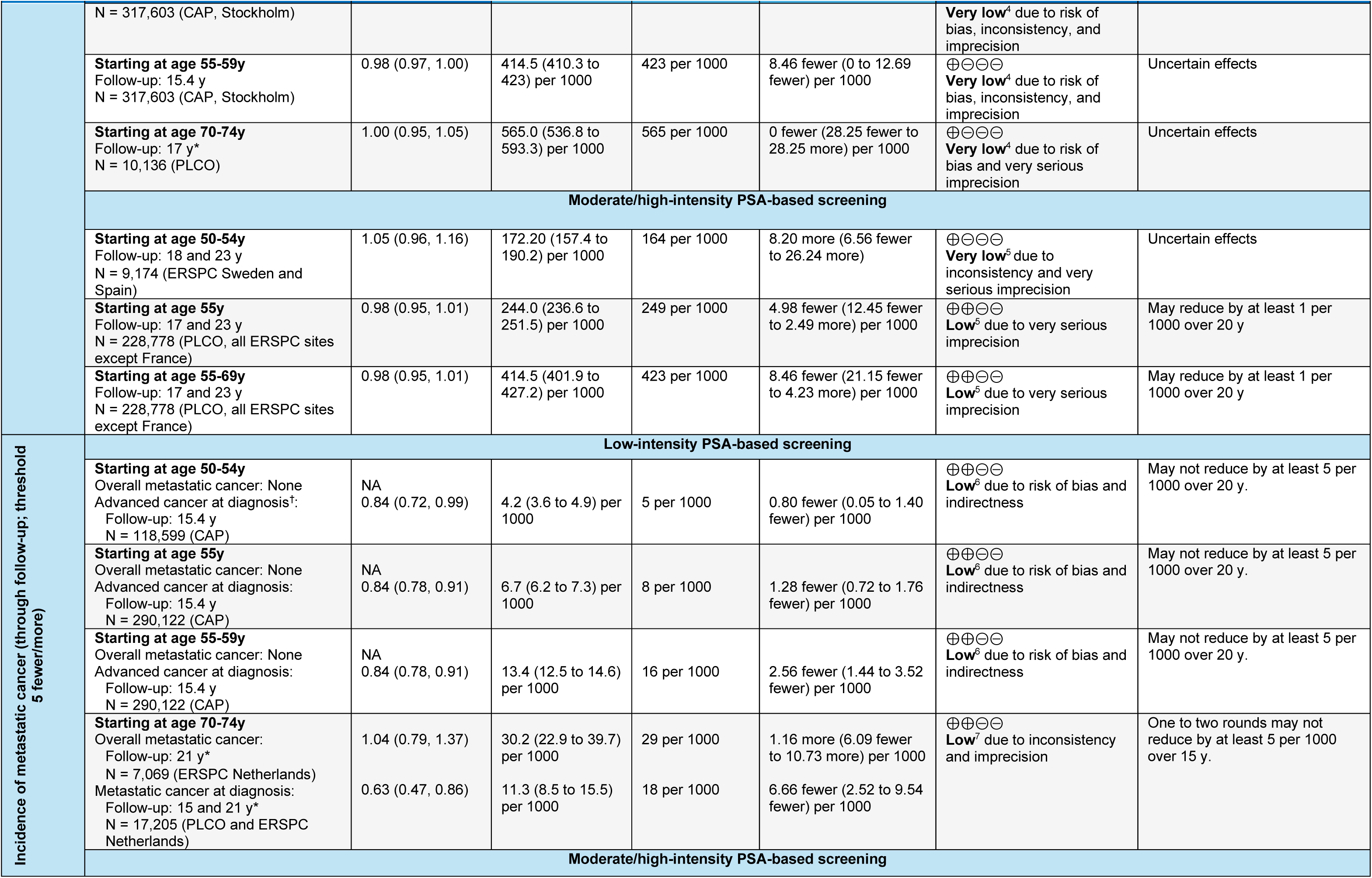
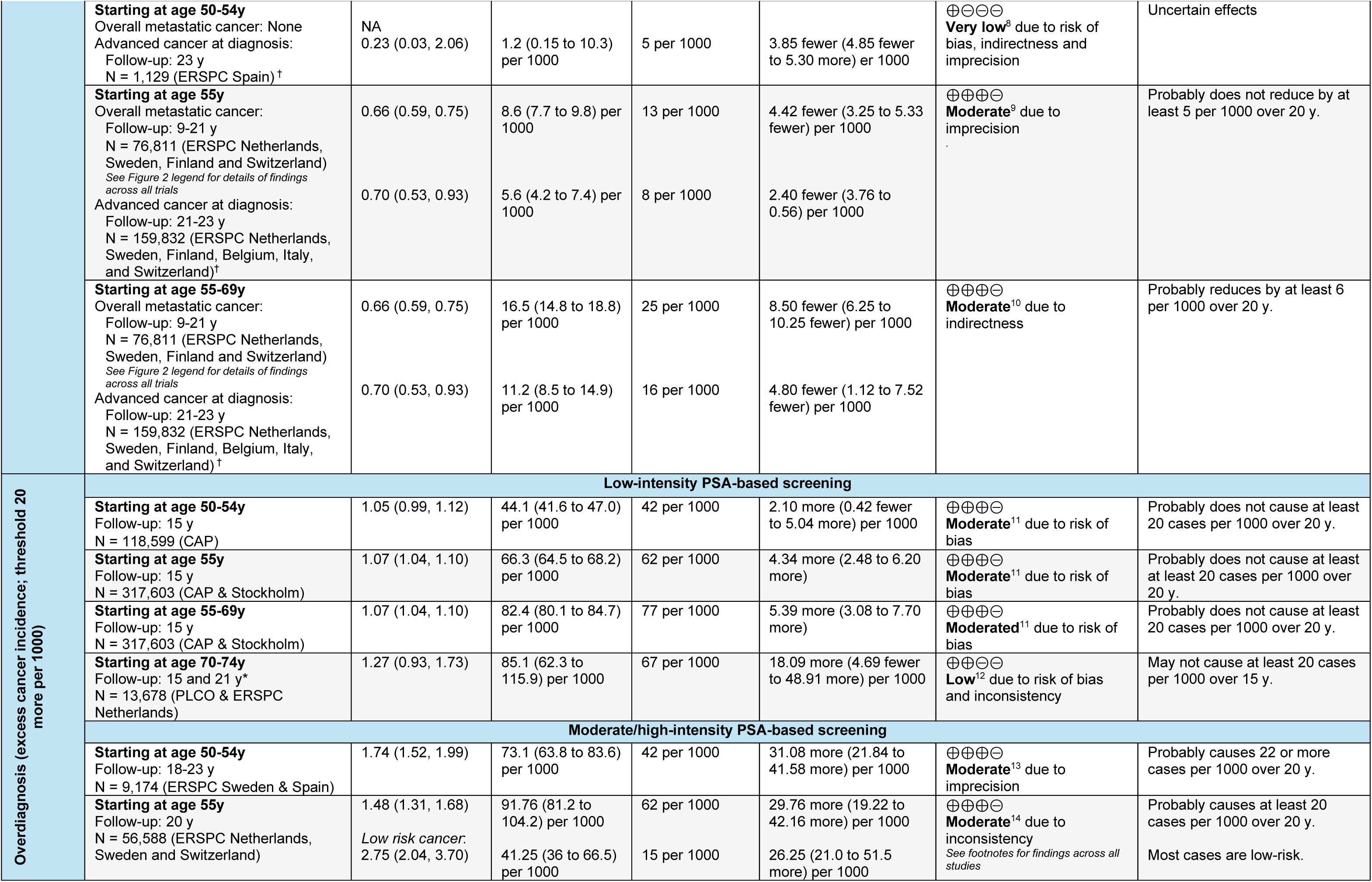
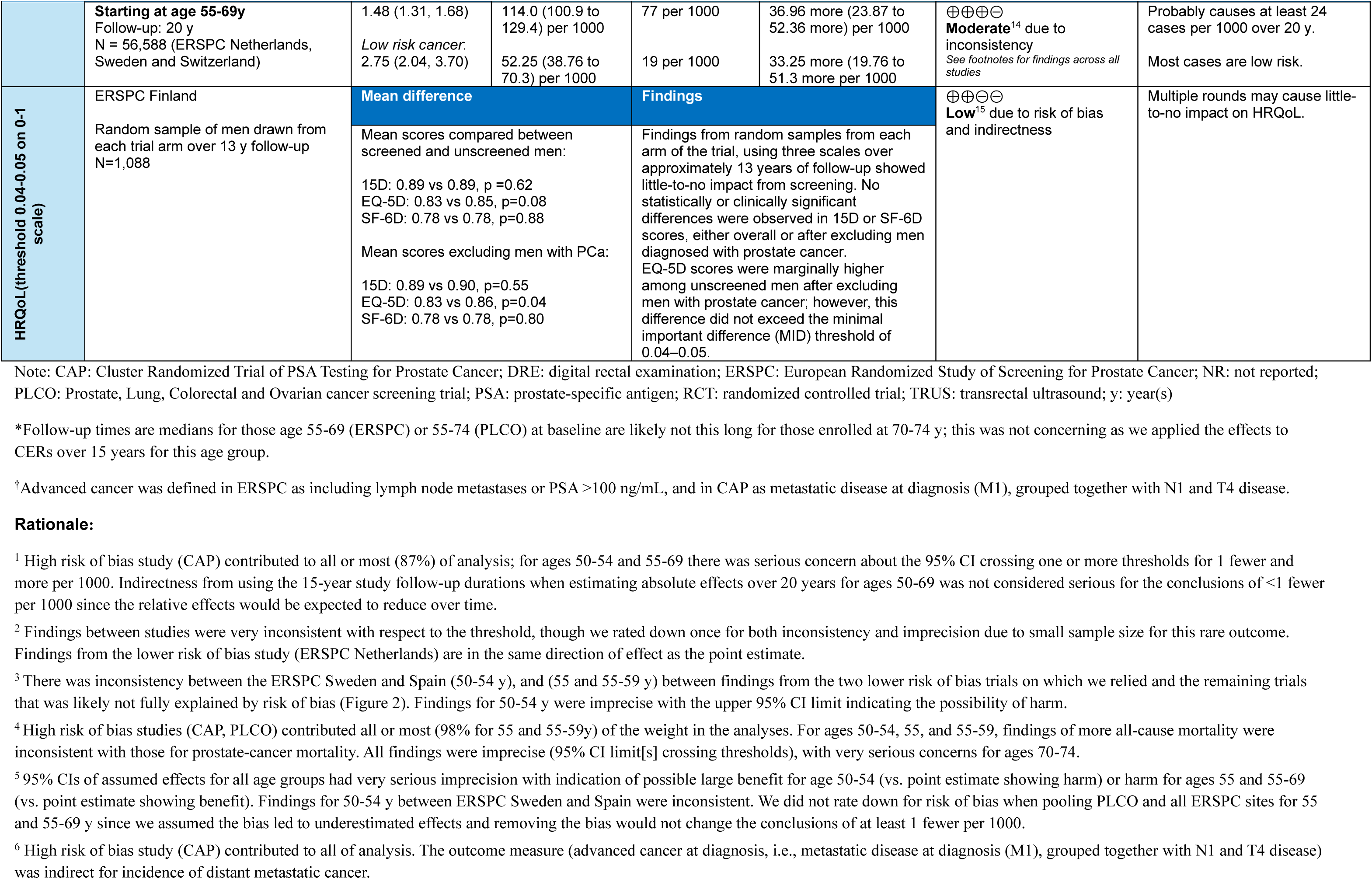
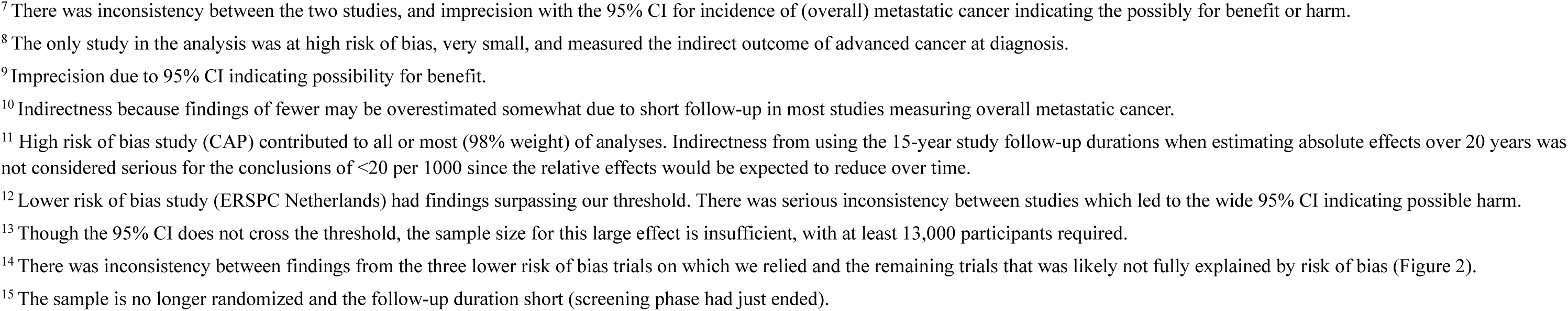
Summary of findings for benefits, overdiagnosis, and HRQoL from PSA-based screening versus no screening (KQ1), by screening intensity and starting age.

**Table 2** (first column), **Figure 3** and **Appendix 1** report the decision thresholds. For all comparative effects, we rated the certainty of evidence for seven circumstances based on screening intensity and age group, namely, for low-intensity screening (mostly one round) starting at ages 50-54, 55, 55-69 or 70-74 and for multiple rounds of screening starting at ages 50-54, 55, or 55-69 (**Table 2**). This categorization was based on several factors: i) moderate-to-high credibility of effect moderation by starting age (e.g., 50-54 vs. 55-69 vs. 70-74 across ERSPC sites at 16 year follow-up^35^ and 55-69 vs. 70-74 in PLCO at 17 years [data received from authors]), ii) no significant differences within age groups between moderate and high-intensity screening or by screening test (i.e. PSA alone vs. PSA or DRE), iii) within-study findings that multiple versus single rounds of screening moderated effects,^35, 45^ and iv) clinically relevant differences between ages (e.g., most 70-74 year olds were offered 1 round in otherwise multi-round interventions).

**Figure 3:**
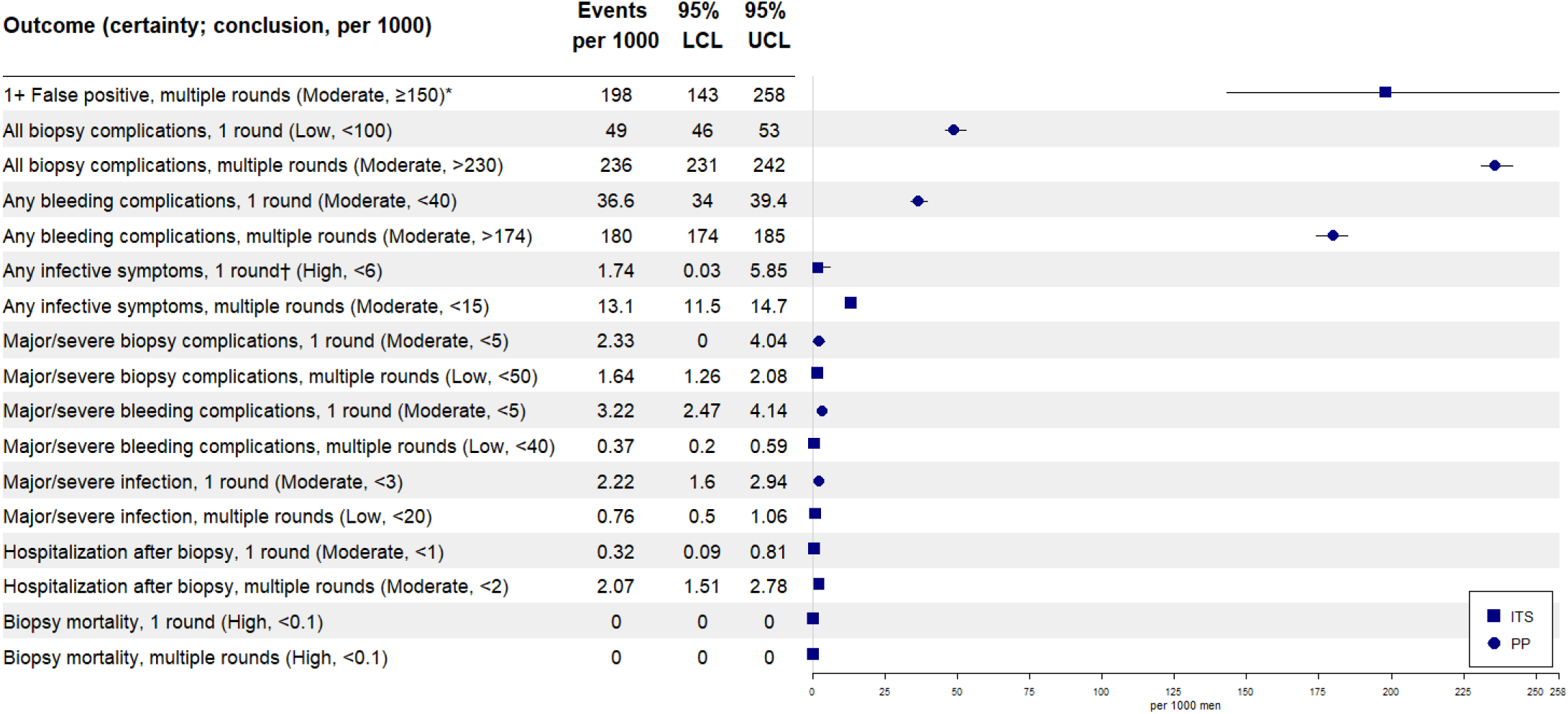
Effects from PSA-based screening on risk for ≥ 1 false positive result (cumulative over multiple rounds) and complications from biopsies after PSA screening (single round and multiple rounds), per 1000 men. Each estimate represents pooled proportions across eligible studies. Certainty levels for the stated conclusions, based on the decision thresholds, are indicated. See Appendix 2 for the summary of findings table with rationale for each certainty assessment. Note: Per protocol (PP) effects were used when intention-to-screen (ITS) data were not available. LCL: lower limit of 95% CI; UCL: upper limit of 95% CI.

For low-intensity screening (including all screening starting at age 70-74 in the trials), there was very low certainty about any benefit for any age, and low certainty for harm from an increase in prostate-cancer mortality for those starting screening at age 70-74. Starting at age 50-54, at 20 years multiple rounds of screening may reduce prostate-cancer mortality by at least 1 per 1000 and probably causes at least 20 cases of overdiagnosis. Findings are uncertain for all-cause mortality and incidence of metastatic cancer. Screening multiple times from age 55 probably reduces prostate-cancer mortality and may reduce all-cause mortality, each by at least 1 per 1000 after 20 years, but probably does not result in at least 5 fewer cases of metastatic cancer and probably causes at least 20 cases of overdiagnosis. If the age to start screening varies between ages 55 and 69 (on average 62), reductions in prostate-cancer mortality (at least 2 fewer per 1000) and incidence of metastatic cancer (at least 6 fewer) are probably larger than when starting at age 50-54 or 55, and screening probably also causes slightly more (24 cases) overdiagnosis. For prostate-cancer mortality, incidence of metastatic cancer, and overdiagnosis, we relied on lower risk of bias findings for starting at age 55 or 55-69. For comparison, absolute effects based on findings across all studies are shown in **Figure 2** (see legend). Though the studies we relied upon were rated as lower risk of bias, it should be noted that these also had higher intensity screening (i.e., offering up to 5-10 rounds of screening, and/or >2 rounds completed on average for ages 55-69) than most other trials (**Table 1**). Findings indicate that most (>80%) cases of overdiagnosis are of low-risk cancer (**Table 2**).

Figure 3 contains pooled proportions and certainty levels for cumulative FPs and biopsy complications after a single and multiple rounds. Age-specific data were not available. The risks for one or more FPs, biopsy complication (any severity), and bleeding from a biopsy are probably at least 150, 230, and 174, respectively, per 1000 men screened multiple times. Findings for major/severe complications, hospitalizations, and mortality from a biopsy were all below the decision thresholds, though there was low certainty in some cases due to risk of bias and reporting bias (i.e., very few RCTs reported these outcomes).

Figure 4 presents findings across most outcomes for men starting to screen multiple times from age 55-69, with follow-up to 20 years. Despite lacking data from eligible studies, we added estimates for long-term treatment harms among the portion of men overdiagnosed (based on treatment studies^81^ and treatments received by men with low-risk disease; see Figure 4 footnotes and **Appendix 4** for details on methods).

**Figure 4.**
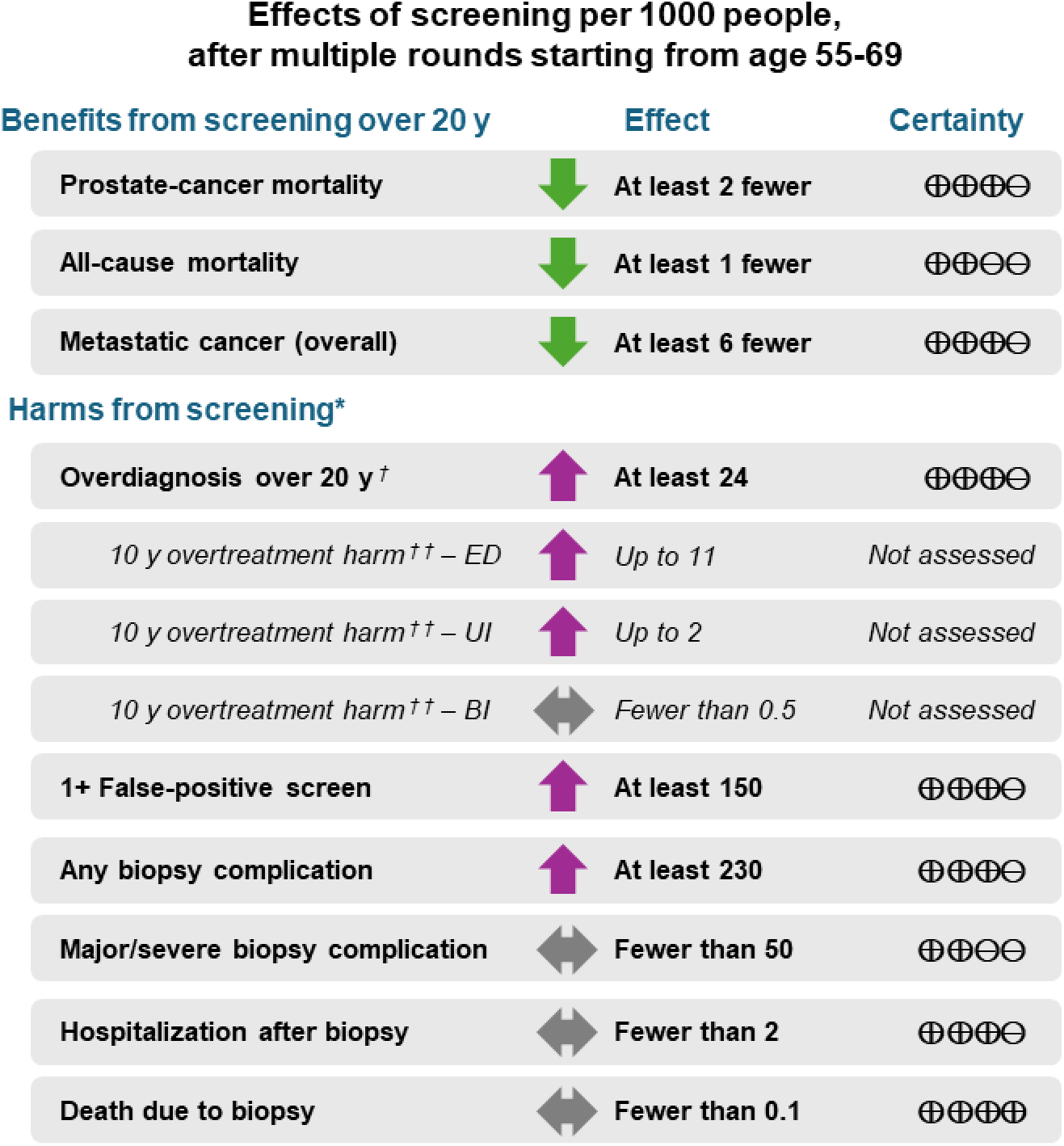
Summary of findings across most outcomes for multiple rounds of PSA screening versus no screening starting at age 55 to 69 and with 20-year follow-up. Certainty assessments are based on the effects stated in the middle of the figure. Of the Any biopsy complication, most are bleeding. Figure 3 presents data on additional harm outcomes. Notes: ⊕⊕⊕⊕=high certainty; ⊕⊕⊕⊝=moderate certainty; ⊕⊕⊝⊝=low certainty. *Harms from screening only occur in the screening group. †As per Table 2, a very large majority of overdiagnosed cases are low-risk disease. ††Because eligible studies did not report data on treatment harms, which are important physical harms from overdiagnosis, these effects were estimated from contemporary cohorts reporting initial treatment choice (active surveillance, radical prostatectomy, radiation therapy) and available long-term (i.e., 10 year) follow-up data from treatment studies (i.e., PROTECT trial83) on erectile dysfunction (ED), urinary incontinence (UI), and bowel incontinence (BI) among men diagnosed with low-risk disease. See Appendix 4 for details and shorter-term harms. Psychological or other impacts (e.g., financial burden) are not estimated.

Scarce data were reported about effects by race, ethnicity or family history (**Appendix 2**). In the PLCO trial which enrolled ∼5% Black participants, prostate-cancer mortality (>2-fold [1.4% vs. 0.64%] at 19 years) and incidence rates (36% higher [14.7% vs. 10.7%] at 13 years) were higher among Black versus White men, but the effects (RRs) from screening were very similar.^44^ Effects from screening on metastatic cancer at diagnosis may be larger for Black men (RR 0.74 vs. 0.97, p=0.43), though findings were not statistically significant. In this trial, Black men had more FPs than did White men (≥1 in 14.5% vs. 12.4%, respectively; p=0.02)^44^ and higher rates of biopsy complications (any complication and infections).^49^ Conversely, in a cohort study (n=2578) in Texas the opposite findings for FPs were found.^28^

### Benefits and Harms from Sequential (PSA+MRI) versus PSA Screening (KQ2)

In all seven eligible studies, a threshold of 3 ng/mL was used for PSA screening in both arms and diagnosis in the PSA alone group relied on systematic 10-12 core biopsies, with one exception (12-18 core used^62^). From all studies, eligible data were only available from one round of screening and across ages (mostly 50-69). Data from five studies allowed comparison of PSA screening to a sequential arm using an MRI threshold of PI-RADS 3 for screening and MRI-targeted (e.g., 2-4 cores per suspicious lesion) biopsies for diagnosis. Findings indicate that replacing PSA screening with sequential screening likely makes little-to-no difference in detection of clinically significant (i.e., Gleason score ≥7) cancer but reduces clinically insignificant cancer detection by at least 10 per 1000 and FPs by at least 33 per 1000 screened (Figure 5, **Table 3**). Two studies were added when exploring subgroups of variations in sequential screening protocols. No subgroup findings had significant interaction effects, though when considering the threshold of 1 more/fewer per 1000 there may be some benefit for clinically significant cancer detection from combing an MRI-targeted and systematic (10-12 core) biopsies compared with only MRI-targeted biopsies (Figure 5, **Table 3**). Findings from per protocol analyses were almost identical.

**Figure 5:**
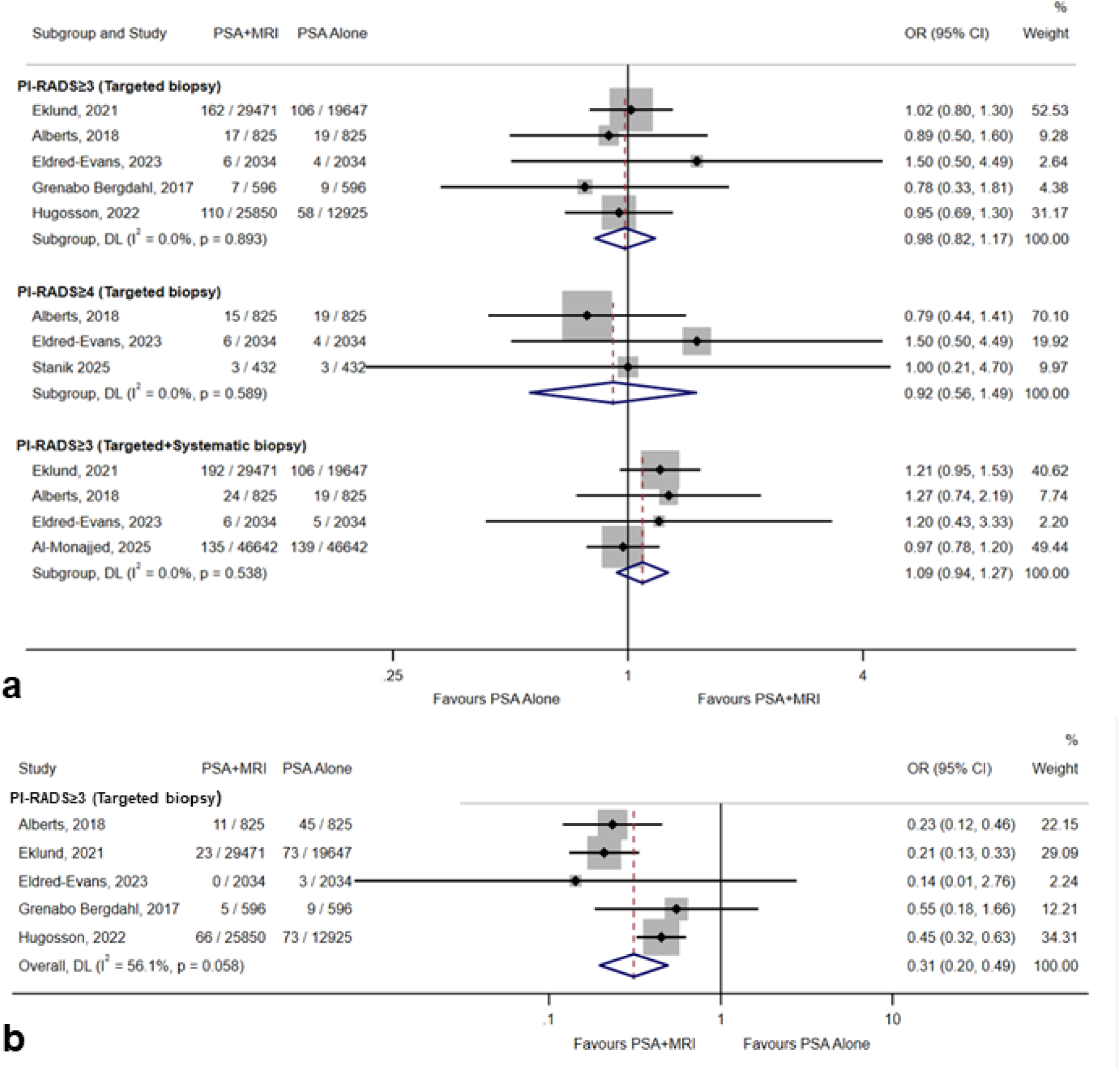
Relative effects from one round of screening with sequential (PSA+MRI) versus PSA alone screening on (a) clinically significant and (b) clinically insignificant cancer detection. For (a), we present data from subgroup analyses based on the MRI screening threshold and biopsy approach in the sequential screening arm because the absolute effects (though without significant interaction effects) differed from the main analysis (PI-RADS ≥3 with targeted biopsy) with respect to our threshold of 1 fewer/more per 1000. For clinically insignificant cancer detection, the effects in all analyses surpassed 10 fewer per 1000 so only the main analysis is shown. See Table 3 for absolute effects and Appendix 3 for all figures.

**Table 3:**
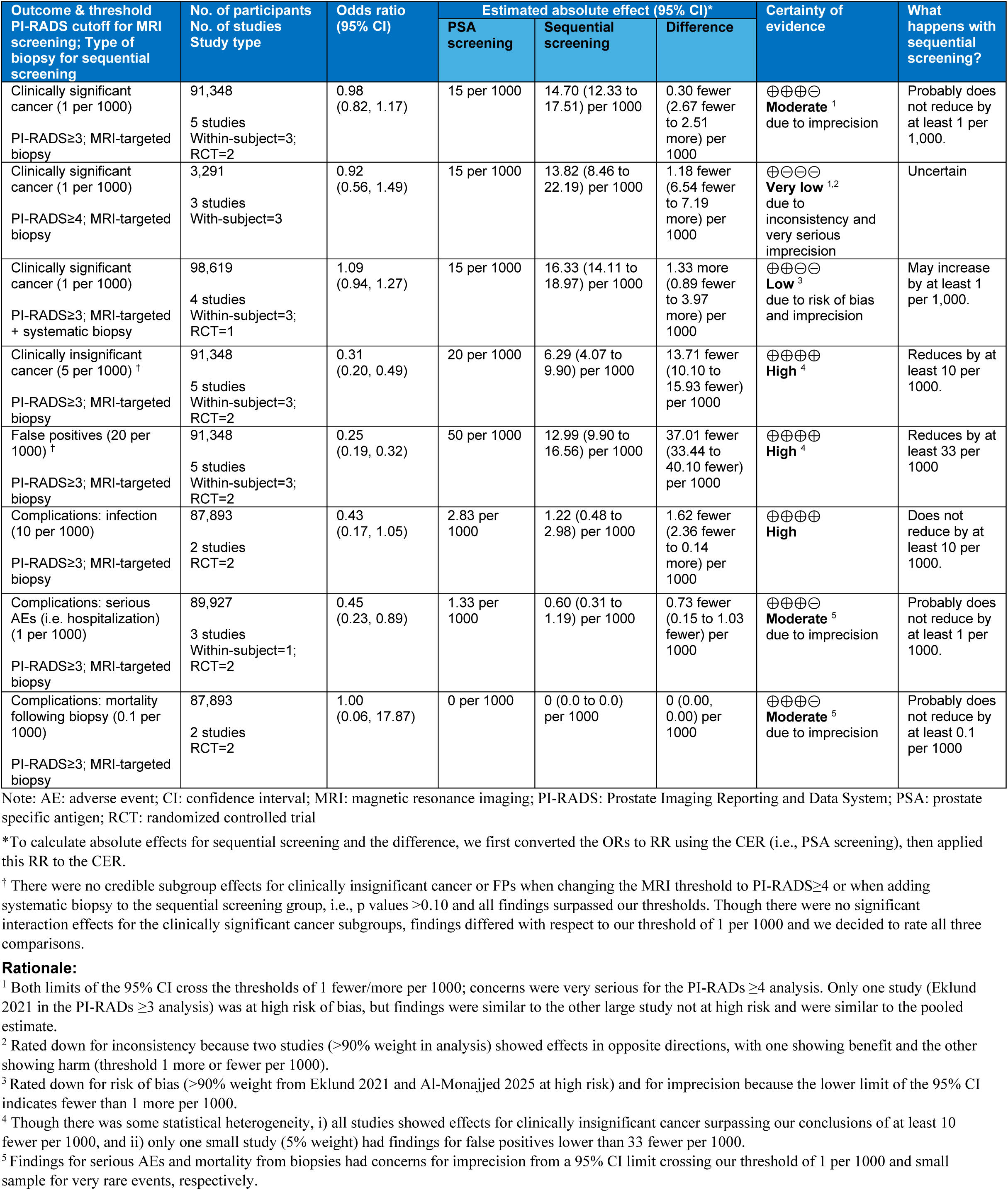
Summary of findings for KQ2 benefits and harms after one round of sequential screening (i.e., MRI for those with positive PSA test) versus PSA screening (PSA≥3 ng/ml threshold and 10-12 core systematic biopsy)

### Interpretation

Trials of prostate-cancer screening that focus on PSA have now all completed screening and provide results after approximately two decades of follow-up. This review provides clinicians and other interest holders with findings using absolute effects, by age, across many critical and important outcomes to inform decisions about screening. The effects on benefits and harms from multiple screening rounds and starting to screen before age 70 should be emphasized. In the trials, screening stopped between 71-74 years and in the trials with the highest certainty evidence the younger men (e.g., 50-60y at enrollment) were offered (with >75% adherence) screening every 2-4 years over a period of 12-20 years. We found no credible evidence showing added benefit from annual screening or adding DRE to PSA testing. Sequential screening with PSA positives triaged to MRI likely reduces (by about half) the number of men experiencing a FP or clinically insignificant cancer (and thus overdiagnosis), though findings are limited to one screening round and did not address long-term effects on mortality; further, costs, infrastructure, accessibility, and technical experience are important considerations.

Our findings differ from those of the 2014 review in several ways. We included additional follow-up data for mortality and overdiagnosis; examined findings by intensity of screening; and applied current guidance for assessing certainty of evidence using absolute findings by age within a Canadian context. We relied on decision thresholds (rather than point estimates) for making conclusions about magnitude of effects. We also examined the potential benefit of adding MRI to PSA screening. Compared with the 2014 recommendations and others (e.g.,^82^) that also favored findings from one or more ERSPC sites, our findings of larger absolute effects will be due to our assessment of risk of bias by site as well as the application of fairly similar RRs to larger CERs due to the longer time horizon we applied.

This review has some limitations. Although comprehensive, our search may have missed observational studies published before 2014 that reported on benefit outcomes from PSA screening. We tried to prevent this by hand searching reference lists. Further, we are aware of two ongoing RCTs relevant to KQ1 on sequential screening versus no screening that have yet to report data from the control group (GÖTEBORG-2)^34^ or benefit outcomes (ProScreen).^83^ We did not formally explore patient preferences. Gathering patient input or reviewing evidence on patient preferences would help validate the applied thresholds, elucidate how patients weigh the benefits and harms, and provide information for shared decision-making. The included trials began many years ago, and screening protocols (e.g., repeat PSA testing, 10-12 core and transperineal biopsies) and treatment approaches (e.g., more active surveillance) have since evolved.^84, 85^ Despite this, clinical input indicated no serious concern about indirectness. Some screening was received by men in both arms (to a similar degree) in the trials during follow-up and this may have diluted the effects over time. Findings on screening with MRI among those with positive PSA tests were limited to one round of screening, did not include effects on benefit outcomes, and were not available by age. We did not include studies directly comparing differences in screening eligibility (e.g., by age or cancer risk) or intervals (e.g., based on PSA values or age), and findings from ongoing studies (e.g., PROBASE^86^, ProScreen^83^) on these topics will be informative.

When using absolute effects, findings differed based on age. No study started to screen everyone at age 55 though we added this scenario to provide estimates of these effects which will be impacted by lower event rates than for those starting at age 55-69 (average 62). Though few data were available about differential (relative) effects of screening by race, ethnicity or family history, variation in baseline risks is the most likely contributor to differing absolute effects^87^ and our calculated absolute effects may not generalize well to some groups. Race and ethnicity likely associate with higher prostate-cancer incidence and mortality in the US and UK, especially for Black men,^88, 89^ likely due to multiple factors including biological differences and social determinants of health negatively impacting screening, treatment, and access to care.^90–92^ In a Canadian cohort, Black men presented with cancer at an earlier age but not with more aggressive disease or poorer survival, whereas Indigenous men (in rural and urban settings) were twice as likely to be diagnosed with metastatic disease possibly due to less use and access to health care, which may include PSA testing/screening.^93, 94^ Men with a first-degree family history of prostate cancer may face about 2.5 fold increased risk for the disease, largely from biological constructs, and there is evidence that family history increases the risk of fatal disease especially if the relative(s) had high-grade or metastatic disease.^92^

### Conclusions

This review provides clinicians and other interest holders with anticipated absolute benefits and harms of PSA screening, by age and with assessments of certainty, across many important outcomes and with approximately two decades of follow-up. The benefit-to-harm profile is likely higher for those screening multiple times every 2-4 years, starting at around 60 years of age, compared with screening once or starting at a younger or older age. Nevertheless, findings for most critical benefits and harms exceeded thresholds, thereby demonstrating the complexity of guideline developers’ and patients’ decision-making regarding screening. Adding DRE or screening annually did not show additional benefit. Findings apply to a general population and may differ for specific groups. Findings about adding MRI for those with a positive PSA test were limited and would require additional consideration of costs, infrastructure, expertise, and equity.

## Data Availability

All data produced in the present work are contained in the manuscript and appendices

## Competing interests

No competing interests were declared.

## Contributors

Jennifer Pillay, Roland Grad, Guylène Thériault, Philipp Dahm, Keith J Todd, Gail Macartney, Brett Thombs conceived the design of the study. Jennifer Pillay, Lindsay Gaudet, Sholeh Rahman and Sabrina Saba acquired the data. Jennifer Pillay, Lindsay Gaudet, and Sholeh Rahman interpreted the data and drafted the manuscript. LH contributed to analysis and interpreting the data. All of the authors revised the manuscript and contributed important intellectual content and gave final approval of the version to be published. The authors acknowledge Maria Tan (MLIS, University of Alberta) for developing and running the searches; Rachel Couban (MISt, McMaster University) for peer reviewing the MEDLINE search; Benjamin Vandermeer (University of Alberta) for running the analyses for KQ2, and Donna Reynolds (University of Toronto) for reviewing the draft manuscript.

## Content licence

This is an Open Access article distributed in accordance with the terms of the Creative Commons Attribution (CC BYNC-ND 4.0) licence, which permits use, distribution and reproduction in any medium, provided that the original publication is properly cited, the use is noncommercial (i.e., research or educational use), and no modifications or adaptations are made. See: https://creativecommons.org/licenses/by-nc-nd/4.0/.

## Funding

Lisa Hartling is funded by a Tier 1 Canada Research Chair in Knowledge Synthesis and Translation. This review was conducted with funding provided by the Public Health Agency of Canada (PHAC). The views expressed in this article do not necessarily represent those of PHAC or the Government of Canada. Staff of the Global Health and Guidelines Division at PHAC (Casey Gray, Heather Limburg) provided input during the development of the protocol and reviewed a draft version of the manuscript, but did not take part in the selection of studies, data extraction, analysis, or interpretation of the findings. The reviews reported here were intended to inform recommendations by the Canadian Task Force on Preventive Health Care; however, this Task Force’s mandate has since concluded and the Public Health Agency of Canada has initiated a renewed Task Force that may or may not issue recommendations on this topic or use these findings for their recommendations.

## Data sharing

The relevant data in this study are available in the manuscript or appendices, or from Lisa Hartling.

## Notes

### Competing Interest Statement

The authors have declared no competing interest.

