## Appendices 1-4 for "Screening for prostate cancer using PSA with and without MRI: systematic reviews with meta-analysis"

DRAFT

Appendices

Contents (click on each to link):

Appendix 1. Additional methods

Appendix 2. Additional information for KQ1

Appendix 3. Additional information for KQ2

Appendix 4. Methods to estimate burden of management harms among overdiagnosed cases of low-risk prostate cancer

**Screening for prostate cancer using PSA with and without MRI: systematic reviews with meta-analysis**

Jennifer Pillay, Lindsay Gaudet, Sholeh Rahman, Roland Grad, Guylène Thériault, Philipp Dahm, Keith J Todd, Gail McCartney, Brett Thombs, Sabrina Saba, Lisa Hartling

### Appendix 1. Additional methods

### DRAFT

#### Contents:

Eligibility criteria for KQ1 and 2

Database search strategies

Additional methods for data extraction and analysis

Modification to NIH risk of bias tool for uncontrolled cohorts

Development of decision thresholds

Development of control event rates

**Screening for prostate cancer using PSA with and without MRI: systematic reviews with meta-analysis**

Jennifer Pillay, Lindsay Gaudet, Sholeh Rahman, Roland Grad, Guylène Thériault, Philipp Dahm, Keith J Todd, Gail McCartney, Brett Thombs, Sabrina Saba, Lisa Hartling

#### Eligibility criteria for KQ1

| | Inclusion criteria | Exclusion criteria |
|---|---|---|
| Population | Individuals from the general population not selected for being at increased risk*<br><br>Specific populations of interest for subgroup analyses:<br>• Age: <55 vs. 55 to <70 vs. ≥70 years<br>• Race or ethnicity<br>• Family history: none/some (e.g., one second or higher degree relative, unspecified) vs. first-degree/strong history**<br><br>* For outcomes of complications from biopsy, if feasible and very low/low certainty from screening studies, we will include studies (>100 participants) limited to those getting biopsy after a positive PSA screening result<br><br>**Strong history, such as (as defined by authors but more than one first degree of any age): 1) people with one brother or father or two or more male relatives with one of the following: a) diagnosed with prostate cancer at age < 60 years; b) any of whom died of prostate cancer; c) any of whom had metastatic prostate cancer. 2) family history of other cancers with two or more cancers in hereditary breast and ovarian cancer syndrome or Lynch syndrome spectrum.(Wie et al. 2023, Giri et al. 2017) | >20% <18 years.<br><br>>20% high-risk for prostate cancer (i.e., working with chemicals known to be carcinogenic, known germline mutations, or strong family history**).<br><br>Studies may include populations who have had a previous PSA screen (including a negative biopsy), and/ or individuals with a “normal” age-related change in urine function. Normal will be defined by clinician judgment. |
| Interventions | PSA +/- DRE<br><br>We will also include interventions using multi-step/sequential screening strategies including PSA (e.g., risk calculation/stratification before PSA, MRI after PSA) as long as there is a comparison of no screening.<br><br>DRE-alone screening<br><br>Other modality used as the primary screening method (e.g., MRI only, MRI + other tests) | |
| Comparators | No screening (may include some opportunistic screening, i.e., initiated during a routine healthcare visit)<br><br>For PSA-alone: PSA with DRE; DRE-alone (KQ1a) | |
| Outcomes | Potential benefits<br>1. Reduced prostate cancer mortality<br>2. Reduced all-cause mortality<br>3. Reduced incidence of metastatic cancer*<br>Potential harms<br>4. Positive screen benign for cancer (i.e., false positive); cumulative risk (i.e., number of people with ≥1 event over 2+ rounds])<br>5. Complications due to biopsy: a. mortality (shortly e.g. 30-day after biopsy and before treatment), b. severe AEs (e.g., hematuria, infections), c. serious AEs/hospitalizations (assuming one event per person if applicable), d. all complications (among all biopsies and among those benign for cancer) (revised to also include any bleeding or infection)<br>6. Incontinence from treatment: a. urinary, b. bowel (risks/binary effects from patient-report ascertained actively)<br>7. Erectile dysfunction from treatment (risks/binary effects from patient-report ascertained actively)<br>8. Overdiagnosis (all cancers and those not deemed clinically important (e.g. Gleason <7 or International Society of Urological Pathology (ISUP) grade <2, as defined by authors); excess number of cancers among those screened vs. not screened (will calculate if not reported but data on incidence/cumulative detection is reported)<br>Either benefit or harm<br>9. Quality of life/functioning: a. generic, b. specific to cancer/prostate cancer; composite scores from scales with acceptable measurement properties (e.g., validity, reliability)<br>10. Psychological effects*<br><br>*Reduced incidence of metastatic cancer and psychological effects were considered important, while all others were rated as critical for decision making by the task force | |

| | Inclusion criteria | Exclusion criteria |
|---|---|---|
| | working group. For metastatic cancer, we were interested in all cases during follow-up (i.e., including progression) but revised our criteria allowed for detection rates as indirect outcomes. | |
| Timing of outcome assessment | For benefits & outcome 8: minimum of 5 years after enrollment<br><br>For outcomes:<br>4: after a minimum of 2 rounds of screening<br>5b-d: any timing; subgroups ≤30 days post-biopsy vs. longer<br>6, 7, 9: any timing; subgroups for short-term (e.g., ≤6 months) vs. longer-term<br>10: after screening, after positive screening result, after knowledge of benign results, longer term (e.g. ≥6 months) (with similar timepoints in controls) | |
| Setting | Settings generalizable to primary care, including organized screening programs<br><br>Any country | |
| Study design | All outcomes: RCTs/quasi-randomized trials; nonrandomized trials (nonRCTs; exposure allocation by researchers/policy) and controlled prospective cohort studies (study designed and participant enrollment before data collection), both with concurrent controls<br><br>For nonRCTs/cohorts, studies must use design or analysis methods to account for multiple confounders (e.g., age, family history)<br><br>Outcomes 4 and 5: uncontrolled prospective cohorts (>100 participants) will be included for PSA screening and (if studies are located that report on 1+ benefit outcome) for other screening interventions. | |
| Publication date, type and language | No date limitations for RCTs/quasi-RCTs of PSA or DRE screening or studies on harms<br><br>2014-onwards for other study designs for benefits of PSA or DRE screening (to limit to studies most relevant to contemporary diagnostic procedures e.g. 10-12 vs. 6 core biopsies, 2014 Grading criteria, and higher use of active surveillance of low-risk cancers), and for any study design on other comparisons e.g. MRI vs. no screening<br><br>Reports of final study results will be included if not peer reviewed (e.g., clinical trial registry data)<br><br>Studies with full texts in English or French | Editorials, commentaries, letters, conference proceedings, government reports, narrative reviews, systematic reviews |

#### Eligibility criteria for KQ2

| | Inclusion criteria | Exclusion criteria |
|---|---|---|
| Population | Individuals from the general population not selected for being at increased risk<br><br>OR<br><br>Studies may be restricted to those ≥18 years old with an elevated* PSA test after screening (e.g. referred from primary care) sent for biopsy<br><br>**definition of elevated to be determined by the included study but must use PSA in ≥80%* | >20% enrolled who were positive on other screening test (e.g. DRE alone) or enrolled after clinical indication/not specified<br><br>Studies that restrict eligibility to those with specific PSA values (e.g., studies that attempt to limit their eligible population to only those with moderately increased PSA (i.e., not all eligible screeners), or having repeat biopsy |

**Screening for prostate cancer using PSA with and without MRI: systematic reviews with meta-analysis**

Jennifer Pillay, Lindsay Gaudet, Sholeh Rahman, Roland Grad, Guylène Thériault, Philipp Dahm, Keith J Todd, Gail McCartney, Brett Thombs, Sabrina Saba, Lisa Hartling

| | | |
|---|---|---|
| Interventions | Sequential screening i.e., PSA with addition of MRI to determine/triage the need for biopsy<br>In studies only enrolling those with an elevated PSA, MRI before biopsy will be the only intervention | Any post-biopsy variable/test (e.g., MRI that stratifies risk of an already diagnosed cancer)<br>Use of MRI only to assist biopsy among all with elevated PSA<br>When MRI is used based on a different PSA threshold or criteria than the comparator group |
| Comparators | Screening with PSA +/- DRE (systematically biopsied after positive screen) | Comparisons of no screening will be eligible for KQ1 |
| Outcomes | Potential benefits<br>1. Reduced prostate cancer mortality<br>2. Reduced all-cause mortality<br>3. Reduced incidence of metastatic cancer*<br>Potential harms<br>4. Positive screen benign for cancer (i.e., false positives = number with benign screen positive / number screened)<br>5. Complications due to biopsy a. mortality (e.g., 30-day), b. severe AEs (e.g., hematuria, infections), c. serious AEs/hospitalizations, d. all complications<br>6. Incontinence: a) urinary, b. bowel; binary/risks using patient-report via active ascertainment<br>7. Erectile dysfunction; binary/risks using patient-report via active ascertainment<br><br>Either benefit or harm<br>8. Quality of life or functioning i. generic and ii) disease-specific; composite scores from scales with acceptable measurement properties (e.g., validity, reliability)<br>9. Psychological effects*<br>Benefit or harm<br>10. Detection of ii) clinically insignificant cancer (e.g., Gleason 6), and ii) clinically significant cancer*<br>11. Biopsy referrals*<br><br>*Outcomes considered important but not critical for decision making. | |
| Timing of outcome assessment | Benefits: minimum of 5 years after enrollment<br>Outcomes 5-8: any timing and 9: after screening, after positive screening result, after knowledge of benign results, longer term (e.g. ≥6 months) (with similar timepoints in controls); subgroups for 5b-d: ≤30 days post-biopsy vs. longer and for 6-8 short-term (e.g., ≤6 months) vs. longer-term | |
| Setting | From studies limited to people receiving biopsies, ≥80% recruited from primary care or organized screening program<br><br>Any country | |
| Study design | All outcomes: RCTs/quasi-randomized; nonrandomized trials and controlled prospective cohort studies, both with concurrent controls<br>For nonRCTs/cohorts, studies must use design or analysis methods to account for multiple confounders; both methods may be undertaken in same participants (e.g., with blinding to results of comparator) | |
| Publication date & type & language | 2014-onwards (all studies in a recent systematic review were published after this date)<br>Reports of final study results will be included even if not peer reviewed (e.g., clinical trial registry)<br>Studies with full texts in English or French | Editorials, commentaries, letters, conference proceedings, government reports, narrative reviews, systematic reviews |

**Screening for prostate cancer using PSA with and without MRI: systematic reviews with meta-analysis**

Jennifer Pillay, Lindsay Gaudet, Sholeh Rahman, Roland Grad, Guylène Thériault, Philipp Dahm, Keith J Todd, Gail McCartney, Brett Thombs, Sabrina Saba, Lisa Hartling

#### Database search strategies

Database(s): Ovid MEDLINE(R) ALL 1946 to January 27, 2026

| # | Searches |
|---|---|
| 1 | exp Prostatic Neoplasms/ |
| 2 | ((prostat* adj3 (neoplas* or cancer* or carcinoma* or adenocarcinom* or tumour* or tumor* or malignan* or metasta* or angiosarcoma* or sarcoma* or teratoma* or lymphoma* or blastoma* or microcytic* or leiomyosarcoma*)) or cspca).tw,kf. |
| 3 | 1 or 2 [Prostate CA] |
| 4 | Mass Screening/ |
| 5 | screen*.tw,kf. |
| 6 | Early Detection of Cancer/ |
| 7 | "early detection".tw,kf. |
| 8 | or/4-7 [Screening] |
| 9 | 3 and 8 [PCa SCREENING] |
| 10 | (controlled clinical trial or randomized controlled trial or pragmatic clinical trial or equivalence trial).pt. |
| 11 | "Controlled Clinical Trials as Topic"/ |
| 12 | exp Randomized Controlled Trials as Topic/ |
| 13 | (allocat* or randomi#ed or randomi#ation? or randomly or RCT or placebo*).tw,kf. |
| 14 | ((singl* or doubl* or trebl* or tripl*) adj (mask* or blind* or dumm*)).tw,kf. |
| 15 | trial.ti. or trial.ab. /freq=2 |
| 16 | or/10-15 [RCT] |

17 9 and 16 [PCa SCREENING - RCT]

18 controlled clinical trial.pt.

19 Controlled Clinical Trial/ or Controlled Clinical Trials as Topic/

20 (control* adj2 trial).tw,kf.

21 Non-Randomized Controlled Trials as Topic/

22 (nonrandom* or non-random* or quasi-random* or quasi-experiment*).tw,kf.

23 (nRCT or non-RCT).tw,kf.

24 (control* adj2 study).tw,kf.

25 Control Groups/

26 (control* adj2 group?).tw,kf.

27 Cohort studies/

28 (cohort adj (study or studies)).tw,kf.

29 Cohort analys$2.tw,kf.

30 Follow-up studies/

31 Longitudinal studies/

32 Prospective studies/

33 ((Follow-up or followup) adj (study or studies)).tw,kf.

34 Longitudinal.tw,kf.

35 Prospective.tw,kf.

36 (match* or propensity or referent).tw,kf.

37 (discontinuity adj3 (regression or design or study)).tw,kf.

38 "instrumental variable*".tw,kf.

| | |
|---|---|
| 39 | "difference in difference".tw,kf. |
| 40 | (pilot adj3 study).tw,kf. |
| 41 | (or/18-40) not (case-control or retrospective).mp. |
| 42 | 9 and 41 [PCa SCREENING - nRCT, pilot studies] |
| 43 | 17 or 42 [PCa SCREENING - RCT, nRCT, pilot studies] |
| 44 | (Animals/ or Models, Animal/ or Disease Models, Animal/) not Humans/ |
| 45 | ((animal or animals or canine* or dog or dogs or feline or hamster* or lamb or lambs or mice or monkey or monkeys or mouse or murine or pig or pigs or piglet* or porcine or primate* or rabbit* or rats or rat or rodent* or sheep* or veterinar*) not (human* or patient*)).ti,kf,jw. |
| 46 | 44 or 45 [MUHC Animal filter] |
| 47 | 43 not 46 [ANIMALS REMOVED] |
| 48 | (editorial or news or newspaper article).pt. |
| 49 | (letter not (letter and (clinical trial or controlled clinical trial or multicenter study or observational study or randomized controlled trial))).pt. |
| 50 | (editorial or commentary).ti. |
| 51 | or/48-50 |
| 52 | 47 not 51 [OPINION PIECES REMOVED] |

**Screening for prostate cancer using PSA with and without MRI: systematic reviews with meta-analysis**

Jennifer Pillay, Lindsay Gaudet, Sholeh Rahman, Roland Grad, Guylène Thériault, Philipp Dahm, Keith J Todd, Gail McCartney, Brett Thombs, Sabrina Saba, Lisa Hartling

Database(s): Embase 1974 to 2026 January 26

| # | Searches |
|---|---|
| 1 | prostate tumor/ |
| 2 | exp prostate cancer/ |
| 3 | ((prostat* adj3 (neoplas* or cancer* or carcinoma* or adenocarcinom* or tumour* or tumor* or malignan* or metasta* or angiosarcoma* or sarcoma* or teratoma* or lymphoma* or blastoma* or microcytic* or leiomyosarcoma*)) or cspca).tw. |
| 4 | or/1-3 [Prostate CA] |
| 5 | mass screening/ |
| 6 | cancer screening/ |
| 7 | screen*.ti,kw. and screen*.ab. |
| 8 | screen.ab. /freq=2 |
| 9 | early cancer diagnosis/ |
| 10 | "early detection".tw. |
| 11 | or/5-10 [SCREENING] |
| 12 | 4 and 11 [PROSTATE CA - SCREENING] |
| 13 | exp randomized controlled trial/ or controlled clinical trial/ |
| 14 | clinical trial/ |
| 15 | exp "controlled clinical trial (topic)"/ |
| 16 | (allocat* or randomi#ed or randomi#ation? or randomly or RCT or placebo*).tw,kw. |

17 ((singl* or doubl* or trebl* or tripl*) adj (mask* or blind* or dumm*)).tw,kw.

18 trial.ti. or trial.ab. /freq=2

19 or/13-18 [RCT FILTER]

20 12 and 19 [PCa SCREENING - RCT]

21 controlled clinical trial/

22 "controlled clinical trial (topic)"/

23 (control* adj2 trial*).tw.

24 (nonrandom* or non-random* or quasi-random* or quasi-experiment*).tw.

25 (nRCT or non-RCT).tw.

26 controlled study/

27 (control* adj2 study).tw.

28 (control* adj2 group?).tw.

29 (cohort adj (study or studies)).tw.

30 cohort analysis/

31 Cohort analys$2.tw.

32 follow up/

33 longitudinal study/

34 prospective study/

35 ((Follow-up or followup) adj (study or studies)).tw.

36 Longitudinal.tw.

37 Prospective.tw.

38 (match* or propensity or referent).tw.

39 (discontinuity adj3 (regression or design or study)).tw.

40 "instrumental variable*".tw.

41 "difference in difference".tw.

42 (pilot adj3 study).tw.

43 (or/21-42) not (case-control or retrospective).mp. [non-RCT]

44 12 and 43 [PROSTATE CA - SCREENING - non-RCT]

45 20 or 44 [PCa SCREENING - RCT, nRCT, pilot studies]

46 exp animal/ or exp animal experimentation/ or exp animal model/ or exp animal experiment/ or nonhuman/ or exp vertebrate/

47 exp human/ or exp human experimentation/ or exp human experiment/

48 46 not 47

49 ((animal or animals or canine* or dog or dogs or feline or hamster* or lamb or lambs or mice or monkey or monkeys or mouse or murine or pig or pigs or piglet* or porcine or primate* or rabbit* or rats or rat or rodent* or sheep* or veterinar*) not human*).mp.

50 48 or 49

51 45 not 50 [ANIMALS REMOVED]

52 (conference abstract or editorial).pt.

53 letter.pt. not (letter.pt. and (clinical trial/ or controlled clinical trial/ or multicenter study/ or randomized controlled trial/))

54 (editorial or commentary).ti.

55 52 or 53 or 54

56 51 not 55 [CONF ABS, OPINION PIECES REMOVED]

Database: *Cochrane* Central Register of Controlled Trials, Issue 1 of 12, January 2026

| ID | Search |
|---|---|
| #1 | [mh "Prostatic Neoplasms"] |
| #2 | ((prostat* NEAR/2 (neoplas* or cancer* or carcinoma* or adenocarcinom* or tumour* or tumor* or malignan* or metasta* or angiosarcoma* or sarcoma* or teratoma* or lymphoma* or blastoma* or microcytic* or leiomyosarcoma*)) or cspca):ti,ab,kw |
| #3 | #1 OR #2 |
| #4 | [mh ^"Mass Screening"] |
| #5 | screen*:ti,ab,kw |
| #6 | [mh ^"Early Detection of Cancer"] |
| #7 | "early detection":ti,ab,kw |
| #8 | #4 OR #5 OR #6 OR #7 |
| #9 | #3 AND #8 |

#### Additional methods for data extraction and analysis

**Data extraction:** We intended to extract dichotomous data by arm (e.g., events among number enrolled) to conduct our own analysis using relative risks. However, in KQ1 many age- and site-specific results were reported only as rate ratios (RRs), so we prioritized this data. For continuous data on HRQoL and psychological effects, we planned to extract mean baseline and endpoint or change scores, standard deviations or other measures of variability, and group sample sizes; only mean scores and p values at follow-up were reported by the single eligible study. For FPs and complications from biopsies, we extracted the number of participants experiencing one or more events among those enrolled (or screened if necessary). For the intervention group in KQ2, we only used data relevant to sequential screening with MRI after a positive PSA test. For example, for within-subject designs where all participants received both a PSA and MRI test, we only used data on cancer detection among those for whom both tests were positive.

**Meta-analysis**: For controlled dichotomous data, we used the DerSimonian Laird model in Review Manager (version 5.4.1, The Cochrane Collaboration, 2020) with the generic inverse variance method. For uncontrolled data on harms, we used the *metaprop* function in the R-package *meta* (version 8.2-1) to pool proportions using inverse-variance weighting, with between-study variances estimated by the restricted maximum-likelihood method because there were few studies and/or a wide range of sample sizes.[1, 2] For KQ2, in STATA (version 17.0) we used the reciprocal of the opposite treatment arm size correction to include trials with no events. Further, when combining data from RCTs and within-subject designs in KQ2, standard errors for the dependent trials were computed manually assuming a proportional distribution of patients based on the fixed marginal distributions. Meta-analysis of continuous data was not undertaken because of a single study reporting on HRQoL. If an analysis would have had 10 or more studies, we would have tested visually (funnel plots) and statistically[3] for small-study bias. To avoid unit-of-analysis errors, for cluster RCTs we used author-reported adjusted effects or would have adjusted findings ourselves.

#### Adaptations to risk bias tool for uncontrolled cohorts

To assess risk of bias in non-comparative outcomes, we adapted the NIH Observational Cohort Tool, removing questions that assess whether the risk of bias in a comparison (see Table below).

To determine overall ratings (Low risk, Some Concerns, High Risk) we considered the number of "No" responses to each question. We rated down a full level (i.e., from Low risk to Some Concerns or Some Concerns to High Risk) for a "No" in each of questions: 3, 7, 11. For all other questions, we rated down a half-step; we did not round ratings (i.e., other than questions 3, 7 & 11, we required a "No" response to 2 questions to rate down).

| Signalling question | Responses |
|---|---|
| 1. Was the research question or objective in this paper clearly stated? | As we were assessing outcomes that were frequently not a primary objective, we considered this question not applicable. |
| 2. Was the screening population clearly specified and defined? | Put Yes if the study specifies eligibility criteria including at minimum:<br>1. Age of eligibility ("all ages" is acceptable, as long as it is clearly stated).<br>2. Proportion of participants with PCa screening history (or exclusion of men with screening history)<br>3. Exclusion of men with a history of PCa or PCa diagnosed before randomization. |
| **3. Was the participation rate of eligible persons at least 75%?** | Put Yes if biopsy attendance was ≥75%.<br><br>Base this on the number of tests, NOT men with 1+ biopsies.<br><br>For survey-based outcomes reporting on a selected sample, must be consecutive or random OR shown to be representative. |
| 4. Were all participants selected/recruited from the same population according to prespecified and uniformly applied inclusion/exclusion criteria? | Not applicable, since we are assessing outcomes in only one group. |
| 5. Are variance and effect estimates provided? | Not Applicable. This question assesses whether a study is adequately powered to detect an association, however we are not conducting comparative analyses and thus statistical power is not relevant. |

| Signalling question | Responses |
|---|---|
| 6. Did authors confirm participants were free from the outcome of interest before screening started?<br><br>*We have modified this question to be about the outcome, instead of the exposure. The tool indicates that whether participants are free from the outcome should be addressed in Question 2, but because most harms are secondary or tertiary outcomes, we think this is more appropriately addressed in a separate question.* | For False Positives put Yes<br>For Biopsy Mortality put Yes<br>For Hospitalization put Yes<br>Otherwise, put Yes if study describes whether or not participants were free from the outcome before biopsy, considering both direct reporting of baseline characteristics and eligibility/exclusion criteria. |
| **7. Was the timeframe of the outcome measurement adequate** | For Complications outcomes, including mortality, put Yes if complications were ascertained within a reasonable time frame (~1 month) OR if authors directly attributed complication events (including deaths) e.g., through chart review. |
| 8. For exposures that can vary in amount or level, did the study examine different levels of the exposure as related to the outcome (e.g., categories of exposure, or exposure measured as continuous variable)? | Not Applicable. We are analyzing different levels of exposure (i.e., different numbers of rounds) separately. |
| 9. Was the same type of screening test or biopsy implemented consistently across all study participants? | “Consistent use” includes type of screening test, biopsy referral thresholds, and/or type of biopsy.<br><br>For False Positives: put Yes if the same screening strategy was used across a large majority (~80%) of participants.<br><br>For Biopsy Complications outcomes, put yes if both the screening strategy and type of biopsy were clearly described and used consistently for a large majority (i.e., 80% of those screened and 80% of those referred to biopsy). |
| 10. Were participants screened more than once over time? | Not applicable. This question is to assess potential for misclassification of the exposure in case exposure status (i.e., screening status) changed over the course of the study. |
| **11. Were outcomes clearly defined, valid, and actively ascertained across all study participants?** | For False Positives: Put Yes if the number of men with 1+ False Positives was directly reported by the study. Put No if False Positives are estimated from total number of men with 1+ positive screen minus screen detected cancers or if there is any indication that biopsy results were not actively ascertained.<br><br>For Biopsy Complications: put Yes if the complications are clearly defined by the study authors, and attempts were made to actively ascertain outcomes from participants using a structured checklist. |
| 12. Were outcome assessors blinded to screening status of participants? | Not applicable, as we assessed non-comparative outcomes. |
| 13. Was loss to follow-up after baseline 20% or less? | Put “Yes” unless there is clear evidence for concern including explicit reports that >20% of participants withdrew or were censored before PCa diagnosis (e.g., through emigration or failed attempts to contact) and/or results are available for <80% of biopsies.<br><br>Death and PCa diagnosis after randomization are not considered censoring events. |
| 14. Were key potential confounding variables measured and adjusted statistically for their impact on the relationship between exposure(s) and outcome(s)? | Not applicable; all individuals had the exposure and thus it is not possible to adjust for confounding on this basis. |

### Development of decision thresholds for KQs 1 and 2

#### Overview & results for KQ1

**Process used, informed by GRADE framework and CORE GRADE guidance.**

1. Provide some background information e.g. control event rates, patient preferences
2. Develop survey to choose a minimally important threshold by outcome
3. Send out survey to members for independent completion (1 week prior to meeting)
4. Discuss results of survey with blinded results provided
5. Reach consensus on threshold for each outcome
6. Apply to review findings & GRADE certainty assessments

##### Background information provided to Working Group

Thresholds used for recent Canadian Task Force reviews on cancer screening

| Outcome | **Breast (per 1000 screened) 3-4 rounds with 10 year follow-up** | **Lung (per 1000 screened) 3-4 rounds with 10 year follow-up** | **Cervical (per 10,000 screened) (over 1 round of screening)** |
|---|---|---|---|
| All-cause mortality | 1 fewer | 0.5 and 1.0 fewer (GRADE for each) | 2 fewer |
| Cancer specific mortality | 0.5 and 1.0 fewer (GRADE for each) | 1 fewer | 2 fewer |
| Other benefit | Advanced stage disease: stage ≥III 2 fewer; stage ≥II 3 fewer; stage IV 1 fewer | | Incidence of CIN 3/3+: 10 fewer |
| Other benefit | Morbidity: requiring chemotherapy or mastectomy: 2 fewer | | Incidence of invasive cervical cancer: 3 fewer |
| FPs requiring imaging +/- biopsy to resolve | 150 | 75 | 300 |
| FPs requiring biopsy | 15 | | |
| Overdiagnosis | 5 | 2.5 | |
| Other harms | 6 interval cancers | Major adverse events (complications or morbidity) from invasive procedure: 2.5 | |
| Other harms | | Death from invasive procedure among those without cancer: 0.1 | |
| Other harms | | Incidental findings: 100 and 150 | |
| Other outcomes | | PROMs: as per tools used eg 5 & 10 on 0-100 scale | |

### Screening for prostate cancer using PSA with and without MRI: systematic reviews with meta-analysis

Jennifer Pillay, Lindsay Gaudet, Sholeh Rahman, Roland Grad, Guylène Thériault, Philipp Dahm, Keith J Todd, Gail McCartney, Brett Thombs, Sabrina Saba, Lisa Hartling

Previous findings and statements from TF cancer screening recommendations **not using thresholds**

| Topic | Recommendation by age | Benefits | Harms |
|---|---|---|---|
| Colorectal cancer (CRC) | 50-59 **weak** for FOBT q 2 yrs or flex sig 2 10 yrs | CRC mortality: FOBT: NNS 2655 over 18 yrs **(0.38 fewer per 1000)**; Flexible Sigmoidoscopy: NNS 1853 (**0.54 fewer per 1000**) over 11 yrs<br>All-cause: all ages: No change with gFOBT (RR 1.00, 95% CI 1.00–1.01) or flexible sigmoidoscopy (RR 1.00, 95% CI 0.96–1.00) | FOBT: FP rate 12 per 1000<br>Flex sig: rare (intestinal perforation occurred in 0.001% (0.01 per 1000), minor bleeding in 0.05% (0.5 per 1000), major bleeding in 0.009% (0.09 per 1000) and death in 0.015% (0.15 per 1000)). |
| | 60-74 **strong for FOBT** q 2 yrs or flex sig 2 10 yrs | CRC mortality: FOBT: **NNS 492 over 18 yrs (2.03 per 1000)**; Flex Sig: NNS 343 (2.92 per 1000) over 11 yrs<br>All-cause all ages: No change was reported with gFOBT (RR 1.00, 95% CI 1.00–1.01) or flexible sigmoidoscopy (RR 1.00, 95% CI 0.96–1.00) | |
| Prostate cancer | <55 strong against<br>55-69 weak against<br>70+ strong against | Screening with the PSA test may lead to **a small reduction** in prostate cancer mortality but not a reduction in all-cause mortality. Did not include results from 3 high ROB RCTs (not showing benefit).<br>Main RCT ERSPC: cancer mortality RR 0.79 (0.69–0.91; ARR 0.128%, or **1.3 fewer per 1000 men 55-69 yrs** invited for screening NNS 769), with a pretrial PSA testing rate of 20%; all-cause RR 1.00 (0.98–1.02)<br>PLCO RCT (RR 1.09, 95% CI 0.87–1.36; 0 deaths from prostate cancer prevented per 10 000 invited for screening**)**; all-cause 0.96 (0.93–1.00) (PLCO trial had a high rate of pretrial PSA testing (52%) & high rates of opportunistic screening). | Harms (e.g., bleeding, infection, urinary incontinence, a false-positive result and overdiagnosis) “are common”.<br>Overdx: 40%–56% of cases diagnosed in screening arm were overdiagnosed<br>FPs: 11.3 to 19.8% **(of those screened at least once**) depending on threshold<br>**Of men who had a biopsy**: death 0.17% & hospitalization 2.07% & hematuria 30.9% (ERSC note: positive PSA test results typically lead to biopsy; so dilute by 5-10 times for overall sample of screened) |

**Notes related to prostate cancer:**

- In the CAP RCT no screening group (50-69 years; ~15% opportunistic screening), at 15 years, the numbers of prostate cancer diagnoses were 7% (70 per 1000), prostate-cancer deaths 0.78% (7.8 per 1000) and all deaths 23% (230 per 1000). About 20% of cancers were metastatic cancer (e.g., 14 of 70) at diagnosis.
- For the *additional number* of erectile dysfunction/bowel or urinary incontinence in the screening versus control group, we will assume there is more detection/overdiagnosis in the screening arm but mostly at early/curative stage. The additional harms will mainly occur here.
- With the main intervention of PSA screening, all/most patients with positive screening test are referred to biopsy (unlike breast and lung cancer where ~10-20% require biopsy).
- Complications from biopsy can include bleeding (quite common), infections, pain, among others. Many will be short lived.
- In an existing patient preferences review: one study reported that men (over 18 years) were willing to accept a substantial overdiagnosis to reduce their risk of prostate cancer mortality https://www.bmj.com/content/350/bmj.h980 “To reduce the prostate cancer mortality by 10% ( 1 fewer) and 50% (5 fewer), the participating men were willing to accept 126 (95% CI 100 to 150) and 231 (95% CI 200 to 250) cases of overdetection in 1000 people

screened, respectively." "People aged 50 or over accepted less overdetection (~100 was acceptable), whereas respondents with a degree or above accepted more (although the latter only in the higher benefit scenario, suggesting that they made a more explicit trade off between benefit and risk)." The respondents were not tested for their understanding of overdiagnosis.

#### Survey

#### Instructions

**Purpose:** To consider the working group's perspective on what patients would consider to be a **minimally important effect** for each outcome considered in relation to prostate cancer screening.
**Survey:** We present a series of questions based on different risk scenarios for each outcome. We want to determine one threshold for each outcome that the working group agrees would be **important to a majority (>50%) of informed patients** during decision-making (i.e., a minimally important effect). At this point, the question is abstract because only one outcome is considered at a time (all the other benefits, harms, or burdens of interventions and their magnitude are not considered). These judgments are challenging. If possible, reflect on the question based on your knowledge of primary studies, previous focus groups, conversations with friends or family, or shared decision-making with patients. We will discuss the answers (blinded to name) across the group at the next meeting and come to consensus.

**A few items to note:**

- *All effects are measured per 1,000 patients screened. If the risk reduction is <1 person (e.g., 0.4 fewer deaths per 1,000) you can think about it as per 10,000 patients screened (e.g., 4 fewer deaths per 10,000).*
- *Some scenarios involve a decrease (e.g., 1 fewer death per 1,000) or increase (e.g. more harms from treatment) because they are compared with what happens to those not screened, while others (i.e. harms attributed to screening) involve only a proportion because we are assuming there are zero (or very few) in a non-screened population e.g., 100 false positives per 1,000.*
- *Please consider your answers in relation to a screening program where patients are followed for at least 15 years after baseline.*
- *There's no right or wrong answer. All thresholds can be trivial, all can be important, or it can vary.*

**Example of survey questions**

Patients are considering the possibility of having a **major/severe infection from a biopsy** during prostate cancer screening. Prostate cancer screening leads to risk in different scenarios in the table below over multiple rounds of screening. **Please choose an option that will reflect whether the majority of patients (>50%) would think this risk is an important or trivial effect.**

| | Risk reduction scenarios |
|---|---|
| a. | For every **1,000** patients that participate in screening, there will be **5** that experience at least 1 major/severe infection from a biopsy. |
| b. | For every **1,000** patients that participate in screening, there will be **10** that receive at least 1 major/severe infection from a biopsy. |
| c. | For every **1,000** patients that participate in screening, there will be **15** that receive at least 1 major/severe infection from a biopsy. |
| d. | For every **1,000** patients that participate in screening, there will be **20** that receive at least 1 major/severe infection from a biopsy. |
| e. | For every **1,000** patients that participate in screening, there will be **25** that receive at least 1 major/severe infection from a biopsy. |
| f. | For every **1,000** patients that participate in screening, there will be **30** that receive at least 1 major/severe infection from a biopsy. |

**Screening for prostate cancer using PSA with and without MRI: systematic reviews with meta-analysis**
Jennifer Pillay, Lindsay Gaudet, Sholeh Rahman, Roland Grad, Guylène Thériault, Philipp Dahm, Keith J Todd, Gail McCartney, Brett Thombs, Sabrina Saba, Lisa Hartling

#### Overview of range of thresholds considered and chosen thresholds

| **Outcome** | **Range of thresholds considered** (5-6 values across range presented) | **Chosen threshold** (per 1000 screened vs. no screened) | **Discussion points/Notes** |
|---|---|---|---|
| PCa mortality | 0.5 – 2 fewer/more per 1000 | 1 fewer/more | |
| All-cause mortality | 0.5 – 2 fewer/more per 1000 | 1 fewer/more | |
| Incidence of metastatic cancer | 2-12 fewer/more per 1000 | 5 fewer/more (3 fewer/more if using metastatic cancer at diagnosis) | Most common was 2 (range 2-10). Some discussion that metastatic prostate cancer is not of critical importance to patients because, even though it's considered terminal, it has a long survival time.<br>For Metastatic cancer at diagnosis, we used data from studies having both overall and at diagnosis (ERSPC Sweden, Netherlands & Switzerland where metastatic at dx was consistently 60-65%) to adjust the threshold (see CER document) |
| False positives | 100-225 per 1000 | 150 | |
| One or more complications from biopsy | 50-200 per 1000 | 100 (80 for bleeding & 40 for infection) | |
| One or more major/severe complications from biopsy | 20-100 per 1000 | 50 | |
| Major/severe infection or bleeding from biopsy | 5-30 per 1000 | 20 infection & 40 bleeding | |
| One or more biopsy complications requiring hospital admission (*majority likely from infection*) | 2-12 | 6 | |
| Death due to biopsy procedure (ideally measured among FPs) | 0.05-0.5 per 1000 | 0.1 | |
| Overdiagnosis | 5-30 per 1000 | 20 | Clinicians reported that in their experience patients (and many doctors) don't understand what "overdiagnosis" means and reported that the Quebec-based tool for shared-decision making uses 40 per 1000 (though this is from data and not a threshold for an important effect and reflects 13 y follow-up which is much shorter than ours). The working group initially suggested to use 30 per |

| | | | |
|---|---|---|---|
| | | | 1000, but then reconsidered to use the median from the poll of 20 when also considering that our baseline rates (of incidence of cancer) for the synthesis will be lower than originally presented because we are reducing the current incidence rates in Canada based on the assumption there is overdiagnosis and that some of the individuals will be at high risk which is not the target population of this guideline. |
| Erectile dysfunction | 2.5-15 per 1000 | 10 | Studies on utility values for these outcomes suggest quite similar disutility across these outcomes.<br>The findings may only be for ~10 year follow-up but this timeframe was considered adequate to know the effects over 15-20 years.<br>The review will also examine effects over shorter term e.g. 1-2 years but have not assigned thresholds (may not GRADE) |
| Urinary incontinence | 2.5-15 per 1000 | 10 | |
| Bowel incontinence | 2.5-15 per 1000 | 10 | |

#### Results for KQ2

##### Process was similar but relied on open discussion rather than blinded results

###### Background information & results

KQ 2 examines **one round** of PSA+MRI vs. PSA screening among generally unscreened samples, with data applicable to 50-69 years and very little age-specific data.

Across multiple relevant studies of PSA screening (e.g. control group in KQ2 studies among generally unscreened samples, PSA screening CAP), an estimate of cancer detection rate from one round of PSA screening for 50-69 years to use for control event rate = 3.5% 35 per 1000 (Clinically significant: 1.5% 15 per 1000 and insignificant: 2.0% 20 per 1000, since more insignificant vs significant cancers are detected in PSA screening)
Rate for FPs: approximately 5% in one round (50 per 1000)
Rate for serious AEs approx. 0.3% in one round (3 per 1000)
For harms we can use study data

| **Outcome** | **Range of thresholds considered** (5 values across range presented) | **Chosen threshold** (per 1000 screened with PSA & MRI vs. screened with PSA alone) |
|---|---|---|
| Clinically significant cancer | 1-5 fewer/more per 1000 | 1 fewer/more |
| Clinically insignificant cancer | 5-13 fewer/more per 1000 | 5 fewer/more |
| False positives | 15-35 fewer/more per 1000 | 20 fewer/more |
| Serious AEs from biopsies | 0.5-2 fewer/more per 1000 | 1 fewer/more |
| Infection | ~25-30% of KQ1 40 | 10 |
| Complications: serious AEs (i.e. hospitalization) | | 1 per 1000 |
| Complications: mortality following biopsy | | 0.1 per 1000 |

#### Development of control event rates

### KQ1

###### Background

CERs (rates among a "no screening" population) will be used with the relative effects from the analyses to calculate estimates of absolute effects. The decisions used input and consensus from the Working Group, before analysis and presentation of findings.

CERs will be estimated for starting screening at 50y, 55y, between 55-69y, and 70-74y, all over an approximate 20 years for the first three and 15 years for the 70-74y age group, knowing that study data reflects this shorter duration and this is realistic for most older patients.

To inform potential CERs, we collected relevant data from recent Canadian statistics/cohorts and from control arms of screening trials where it was judged there was an acceptable level *ad hoc* screening/contamination (e.g. <20-25%; CAP, ERSPC Netherlands and Sweden) (see following pages). For each outcome we used the best approach to choose among the data sources, as described below.

##### Summary by outcome

**For overdiagnosis (excess incidence):** we used Canadian 2019 incidence data (see below) as the most applicable, though reduced the rates with assumption that some excess incidence is created by *ad hoc* screening. For these calculations we used an estimate of overdiagnosis from longestterm follow-up among the included trials (i.e., across all sites of the ERSPC trial at 23 y with an average of 2 screens per man [30% higher incidence]). This reduction will also help account for the fact that some people among the overall Canadian statistics will be at high risk and not the target for this review.

**For prostate-cancer mortality** we mostly relied on the data by age from the lower risk of bias RCTs at longest follow-up. For the estimate for screening from 55 yr (over 20 years) we relied on information that about 80% of PCa deaths in Canada occur among those 70+years (i.e. at most 25% [8 of 33 per 1000 total over lifespan] among 55-75 yr olds).

**For incidence of (overall) metastatic cancer and metastatic cancer at diagnosis.** RCT data was mostly used for both outcomes, and this data agreed with Canadian information of approximately 60% mortality rates (somewhat higher in oldest) for those with metastatic disease (at any point in the follow-up). From CAP 55-69 year old data, metastatic cancer at diagnosis is about 15% of overall cancer detection over 15 years. Data from 3 ERSPC sites, when comparing metastatic cancer at diagnosis with that through follow-up, consistently showed that rates of metastatic cancer at diagnosis are about 60-65% of overall metastatic cancer.

**For all-cause mortality** we used Canadian data**.**

#### Data Sources: Canadian

Canadian Cancer Statistics 2025: Lifetime risk of prostate cancer: 12.3% (123 per 1000 [95 if removing the effects of overdiagnosis]); Lifetime risk of prostate-cancer death 3.3% (33 per 1000; >80% among 70+ y); 41% 5-year survival for stage 4/metastatic (i.e. 60% mortality)

The other data sources were 2019 incidence rates by age and 2023 all-cause mortality rates by age (see Table). Data on prostate-cancer mortality by age and metastatic cancer by age by was not appropriate due to older data (1999) and only capturing metastatic cancer at diagnosis (not through follow-up), respectively.

| Incidence | PCa mortality | PCa metastases | All-cause |
|---|---|---|---|
| Statistics Canada in 2019 (excluding Quebec)- annual incidence<br>50-54 y: 73.5 per 100,000=367.5 (for 5 yr)<br>55-59 y: 173.9 per 100,000=869.5<br>60-64 y: 319.9 per 100,000 =1599.5<br>65-69 y: 524.5 per 100,000=2,622.5<br>70-74 y: 598.3 per 100,000=2,991.5<br>75-79 y: 592.8 per 100,000=2,964<br>80-84 y: 551.0 per 100,000=2,755<br>85-89 y: 519.6 per 100,000=2,598<br>≥90 y: 449.9 per 100,000=2,249.5 | Only used for 50y | Not used due to data at diagnosis only over 9 years | Statistics Canada 2023<br>50-54 y: 4.0 per 1,000<br>55-59 y: 6.1 per 1,000<br>60-64 y: 9.0 per 1,000<br>65-69 y: 13.7 per 1,000<br>70-74 y: 21.0 per 1,000<br>75-79 y: 33.5 per 1,000<br>80-84 y: 58.4 per 1,000<br>85-89 y: 106.2 per 1,000<br>≥90 y: 220.8 per 1,000 |

**Screening for prostate cancer using PSA with and without MRI: systematic reviews with meta-analysis**

Jennifer Pillay, Lindsay Gaudet, Sholeh Rahman, Roland Grad, Guylène Thériault, Philipp Dahm, Keith J Todd, Gail McCartney, Brett Thombs, Sabrina Saba, Lisa Hartling

| | |
|---|---|
| Calculation of incidence rates e.g. for starting at 50y, use rates for 50-69y over 20 years: 5,459 per 100000 = 5.5%; 55 per 1000 over 20y; for starting at 55-69y we calculated estimates for 55, 60 and 65 (as above for 50y) and took an average.<br><br>55=8,083 per 100000=8.1%/81 per 1000<br>60=10,177.5 per 100000=10.2%/102 per 1000<br>65=11,333 per 1000 =11.3%/113 per 1000<br>70=11,309 per 100000=11.3%/113 per 1000 (8711 over 15 years)<br>**Average forages 55-69=100** | Calculated as per incidence:<br>50=164 per 1000<br>55=249 per 1000 (144 over 15)<br>60=386 per 1000 (219 over 15y)<br>65=633 per 1000 (341 over 15 y)<br>70=565 (over 15 yrs) |

#### Data from control groups in KQ1 RCTs

| Age | Incidence | PCa mortality | PCa metastases (overall & at diagnosis) | All-cause mortality |
|---|---|---|---|---|
| 50-54 | ERSPC Sweden (<55y)-18 y<br>314/3,998 7.85%<br>(Hugosson 2018)<br><br>CAP (50-54)-15.4y<br>2,198/63,398 3.47%<br>(author contact) | ERSPC Sweden (<55y)-18 y<br>18/3,998 0.45%<br>(Hugosson 2018)<br><br>CAP (50-54)-15.4y<br>154/63,398 0.24%<br>(author contact) | <u>Metastatic cancer at diagnosis</u><br>CAP (50-54)-15.4y<br>348/63,398 0.55%<br>(author contact) | ERSPC Sweden (<55y)-18y<br>703/3,998 17.58%<br>(Hugosson 2018)<br><br>CAP (50-54)-15.4 y<br>7,266/63,398 11.46%<br>(author contact) |
| 55-69 | CAP (55-69); 15.4 y<br>10,760/155,997 6.90%<br>(author contact)<br><br>ERSPC Netherlands – 21 y<br>1,706/17,389 9.81%<br>(de Vos 2023)<br><br>ERSPC Sweden (55-64)-18 y<br>648/5,951 10.89%<br>(Hugosson 2018)<br><br>ERSPC across sites-23y<br>9,870/89,348 11.05%<br>(Roobol 2025) | CAP (55-69) 15.4 y<br>1,297/155,997 0.83%<br>(author contact)<br><br>ERSPC Netherlands-21 y<br>268/17,389 1.54%<br>(de Vos 2023)<br><br>ERSPC – Sweden -18y (55-64)<br>104/5,951 1.74%<br>55-59: 49/3164=1.55%=15.5 per 1000<br>(Hugosson 2018)<br><br>ERSPC across sites-23 y<br>1,385/89,348 1.5%<br>(Roobol 2025)<br><br>ERSPC across sites 16y<br>793/89351=8.9% = 9 per 1000<br>55-59: 0.42 per 1000 py<br>60-64: 0.70 per 1000 py | <u>Overall metastatic CA</u><br><br>ERSPC Netherland-21y<br>439/17,389 2.52%<br>(de Vos 2023)<br><br>ERSPC-Sweden 14.9y<br>(55-64)<br>107/5,951 1.80%<br>(Schroder 2012)<br><br><u>Metastatic cancer at diagnosis</u><br>CAP (55-69)-15.4y<br>1,980/155,997 1.27%<br>(author contact)<br>ERSPC Netherlands -21y:<br>278/17389 (63% of incidence/overall)<br>RSPC Sweden -14.9y<br>At diagnosis 70/5,951 vs. overall 107/5,951 vs.= 65%<br>ERSPC Switzerland -9.1 y<br>At diagnosis 13/4,955 vs. overall 23/4,955 = 56% | CAP (55-69)-18y<br>43,070/155,997 27.6%<br>(author contact)<br><br>ERSPC Sweden (55-64)-18y<br>2,154/5,951 36.2%<br>(Hugosson 2018)<br><br>ERSPC-Finland (55-67) 16.8 y<br>16,506/48,299 34.2%<br>(Kilpelainen, 2016)<br><br>ERSPC Netherlands- 21 y<br>7,870/17,755 44.3%<br>(author contact)<br><br>ERSPC across sites -23y<br>43,829/89,348 49.1%<br>(Roobol 2025) |

**Screening for prostate cancer using PSA with and without MRI: systematic reviews with meta-analysis**
Jennifer Pillay, Lindsay Gaudet, Sholeh Rahman, Roland Grad, Guylène Thériault, Philipp Dahm, Keith J Todd, Gail McCartney, Brett Thombs, Sabrina Saba, Lisa Hartling

| | | | | |
|---|---|---|---|---|
| 70-74 | ERSPC Netherlands -<21 y<br>CG: 2,991/3,582 – 8.35%<br>(de Vos 2023) | ERSPC Netherlands-<21 y<br>CG: 73/3,527 2.07%<br>(de Vos 2023) | ERSPC Netherland-<21y<br>Overall metastatic CA<br>101/3,527 2.86%<br>(de Vos 2023) | ERSPC Netherlands -<21 y<br>2,991/3,582 83.5%<br>(author contact) |

#### Final CERs

| | Over 20 y follow-up (per 1000) (15 y only for 70-74 age group) | | | | |
|---|---|---|---|---|---|
| Starting age | Incidence (CAN data)- crude & adjusted for 30% overdiagnosis | PCa mortality | PCa metastases (overall) | PCa metastatic at dx (60-65% of overall) | All-cause |
| 50 y | 55 = 42 | 5 | 8 | 5 | 164 |
| 55 y | 81 = 62 | 8 | 13 | 8 | 249 |
| 55-69 y | 100 = 77 | 15 | 25 | 16 | 423 |
| 70 y | 87 = 67 | 21 | 29 | 18 | 565 |

## KQ2

Across multiple relevant studies of PSA screening (e.g. control group in KQ2 studies among generally unscreened samples, PSA screening CAP), an estimate of cancer detection rate from one round of PSA screening for 50-74 years to use for control event rate = 3.5% 35 per 1000 (Clinically significant: 1.5% 15 per 1000 and insignificant: 2.0% 20 per 1000, more insignificant vs significant cancers detected in PSA screening)

Rate for FPs: approximately 5% in one round (50 per 1000)

For complications we will use study data from the PSA group

**Screening for prostate cancer using PSA with and without MRI: systematic reviews with meta-analysis**

Jennifer Pillay, Lindsay Gaudet, Sholeh Rahman, Roland Grad, Guylène Thériault, Philipp Dahm, Keith J Todd, Gail McCartney, Brett Thombs, Sabrina Saba, Lisa Hartling

### Appendix 2. Additional information for KQ1

### DRAFT

**Contents:**

Summary of risk of bias assessments

- Comparative effects (ROB2 tool)
- FPs and biopsy complications (Adapted NIH tool)

Additional figures on comparative data

Subgroup analysis findings from comparative data

Within-study analysis of effects by screening intensity or per protocol estimates

Within-study analysis for specific populations

Findings from the single study reporting on DRE screening

Study characteristics of studies only reporting on FP and/or biopsy complications

Summary of findings for FPs and biopsy complications

Within-study subgroup data on FPs and biopsy complications

#### Summary of risk of bias assessments

##### Comparative effects (ROB2)

##### A: Prostate-cancer mortality

**≤54-year-old age group**

| Study ID | D1 | D2 | D3 | D4 | D5 | Overall |
|---|---|---|---|---|---|---|
| ERSPC-Sweden | + | ! | + | + | + | ! |
| ERSPC-Spain | - | ! | + | + | + | - |
| CAP | ! | - | + | + | + | - |

**55-69-year-old age group**

| Study ID | D1 | D2 | D3 | D4 | D5 | Overall |
|---|---|---|---|---|---|---|
| ERSPC-Finland | + | - | + | + | + | - |
| ERSPC-Netherlands | ! | ! | + | + | + | ! |
| ERSPC-Sweden | + | ! | + | + | + | ! |
| ERSPC-Italy | + | - | + | + | + | - |
| ERSPC-Belgium | ! | - | + | + | + | - |
| ERSPC-Switzerland | ! | - | + | + | + | - |
| ERSPC-Spain | - | - | + | + | + | - |
| ERSPC-France | + | - | + | + | + | - |
| PLCO | + | - | + | + | + | - |
| CAP | ! | - | + | + | + | - |
| Stockholm | + | + | + | ! | + | ! |

**70-74-year-old age group**

+ Low risk
! Some concerns
- High risk

D1 Randomisation process
D2 Deviations from the intended interventions
D3 Missing outcome data
D4 Measurement of the outcome
D5 Selection of the reported result

| | | | | | | |
|---|---|---|---|---|---|---|
| ERSPC-Netherlands | ! | ! | + | + | + | ! |
| PLCO | + | - | + | + | + | - |

#### B: All-cause mortality

| Study ID | D1 | D2 | D3 | D4 | D5 | Overall |
|---|---|---|---|---|---|---|
| **≤54-year-old age group** | | | | | | |
| ERSPC-Sweden | + | ! | + | + | + | ! |
| ERSPC-Spain | - | ! | + | + | + | - |
| CAP | ! | ! | + | + | + | ! |
| **55-69-year-old age group** | | | | | | |
| ERSPC-All sites (excluding France) | ! | ! | + | + | + | ! |
| PLCO | + | ! | + | + | + | ! |
| CAP | ! | ! | + | + | + | ! |
| Stockholm | + | + | + | + | + | + |
| **70-74-year-old age group** | | | | | | |
| PLCO | + | ! | + | + | + | ! |

#### C: Overdiagnosis (i.e., cancer incidence over long-term follow-up

| Study ID | D1 | D2 | D3 | D4 | D5 | Overall |
|---|---|---|---|---|---|---|
| **≤54-year-old age group** | | | | | | |
| ERSPC-Sw eden | + | ! | + | + | + | ! |
| ERSPC-Spain | - | ! | + | + | + | - |
| CAP | ! | - | + | + | + | - |
| **55-69-year-old age group** | | | | | | |
| ERSPC-Finland | + | - | + | + | + | - |
| ERSPC-Netherlands | ! | ! | + | ! | + | ! |
| ERSPC-Sw eden | + | ! | + | + | + | ! |
| ERSPC-Italy | + | - | + | + | + | - |
| ERSPC-Belgium | ! | - | + | + | + | - |
| ERSPC-Sw itzerland | ! | ! | + | + | + | ! |
| ERSPC-Spain | - | ! | + | + | + | - |
| ERSPC-France | + | - | + | + | + | - |
| PLCO | + | - | ! | - | + | - |
| CAP | ! | - | + | + | + | - |
| Stockholm | + | + | + | ! | + | ! |
| **70-74-year-old age group** | | | | | | |
| ERSPC-Netherlands | ! | ! | + | ! | + | ! |
| PLCO | + | - | ! | - | + | - |

#### D: Incidence of metastatic prostate cancer

**Metastatic (at diagnosis), ≤54-year-old age group**

| Study ID | D1 | D2 | D3 | D4 | D5 | Overall |
|---|---|---|---|---|---|---|
| ERSPC-Spain | - | ! | + | + | + | - |
| CAP | ! | - | + | + | + | - |

**Metastatic (at diagnosis), 55-69-year-old age group**

| Study ID | D1 | D2 | D3 | D4 | D5 | Overall |
|---|---|---|---|---|---|---|
| ERSPC-Finland | + | ! | + | + | + | ! |
| ERSPC-Netherlands | ! | ! | + | ! | + | ! |
| ERSPC-Sweden | + | ! | + | + | + | ! |
| ERSPC-Italy | + | ! | + | + | + | ! |
| ERSPC-Belgium | ! | ! | + | + | + | ! |
| ERSPC-Switzerland | ! | ! | + | ! | + | ! |
| ERSPC-Spain | - | ! | + | + | + | - |
| PLCO | + | - | ! | - | + | - |
| CAP | ! | - | + | + | + | - |

**Metastatic (at diagnosis), 70-74-year-old age group**

| Study ID | D1 | D2 | D3 | D4 | D5 | Overall |
|---|---|---|---|---|---|---|
| ERSPC-Netherlands | ! | ! | + | ! | + | ! |
| PLCO | + | ! | ! | - | + | - |

**Metastatic (through follow-up), 55-69-year-old age group**

| Study ID | D1 | D2 | D3 | D4 | D5 | Overall |
|---|---|---|---|---|---|---|
| ERSPC-Finland | + | ! | + | + | + | ! |
| ERSPC-Netherlands | ! | ! | + | ! | + | ! |
| ERSPC-Sweden | + | ! | + | + | + | ! |
| ERSPC-Switzerland | ! | ! | + | ! | + | ! |

| **Metastatic (through follow-up), 70-74-year-old age group** | | | | | | |
|---|---|---|---|---|---|---|
| ERSPC-Netherlands | ! | ! | + | ! | + | ! |

##### FPs and biopsy complications (Adapted NIH tool)

##### Intention-to screen data

| Study | Q2 | Q3 | Q6 | Q7 | Q9 | Q11 | Q13 | Overall |
|---|---|---|---|---|---|---|---|---|
| **False Positives** | | | | | | | | |
| ERSPC Belgium | ✗ | ✗ | ✓ | ✓ | ✗ | ✗ | ✓ | High risk |
| ERSPC Finland | ✓ | ✓ | ✓ | ✓ | ✓ | ✗ | ✓ | Some concerns |
| ERSPC France | ✓ | ✗ | ✓ | ✓ | ✓ | ✗ | ✓ | High risk |
| ERSPC Italy | ✗ | ✗ | ✓ | ✓ | ✓ | ✗ | ✓ | High risk |
| ERSPC Netherlands | ✓ | ✓ | ✓ | ✓ | ✓ | ✗ | ✓ | Some concerns |
| ERSPC Spain | ✓ | ✓ | ✓ | ✓ | ✓ | ✗ | ✓ | Some concerns |
| ERSPC Sweden | ✓ | ✓ | ✓ | ✓ | ✗ | ✗ | ✓ | Some concerns |
| ERSPC Switzerland | ✗ | ✓ | ✓ | ✓ | ✓ | ✗ | ✓ | Some concerns |
| PLCO | ✓ | ✗ | ✓ | ✓ | ✓ | ✓ | ✓ | Some concerns |
| **Biopsy mortality, 1st round** | | | | | | | | |
| CAP | ✗ | ✓ | ✓ | ? | ✓ | ✓ | ✓ | Some concerns |
| ERSPC Belgium | ✗ | ✗ | ✓ | ✓ | ✓ | ✓ | ✓ | Some concerns |
| ERSPC Finland | ✓ | ✓ | ✓ | ✓ | ✓ | ✓ | ✓ | Low risk |
| ERSPC Italy | ? | ✗ | ✓ | ✓ | ✓ | ✓ | ✓ | Some concerns |
| ERSPC Netherlands | ✓ | ✓ | ✓ | ✓ | ✓ | ✓ | ✓ | Low risk |
| ERSPC Spain | ✓ | ✓ | ✓ | ✓ | ✓ | ✓ | ✓ | Low risk |
| ERSPC Sweden | ✓ | ✓ | ✓ | ✓ | ✗ | ✓ | ✓ | Low risk |
| ERSPC Switzerland | ✗ | ✓ | ✓ | ✓ | ✓ | ✓ | ✓ | Low risk |
| Göteborg-2 | ✓ | ✓ | ✓ | ✓ | ✓ | ✓ | ✓ | Low risk |
| PLCO | ✓ | ✗ | ✓ | ✓ | ✓ | ✓ | ✓ | Some concerns |

| | | | | | | | | |
|---|---|---|---|---|---|---|---|---|
| **Biopsy mortality, ≥2 rounds** | | | | | | | | |
| ERSPC Belgium | ✗ | ✗ | ✓ | ✓ | ✓ | ✓ | ✓ | Some concerns |
| ERSPC Finland | ✓ | ✓ | ✓ | ✓ | ✓ | ✓ | ✓ | Low risk |
| ERSPC Italy | ? | ✗ | ✓ | ✓ | ✓ | ✓ | ✓ | Some concerns |
| ERSPC Netherlands | ✓ | ✓ | ✓ | ✓ | ✓ | ✓ | ✓ | Low risk |
| ERSPC Spain | ✓ | ✗ | ✓ | ✓ | ✓ | ✓ | ✓ | Some concerns |
| ERSPC Sweden | ✓ | ✓ | ✓ | ✓ | ✗ | ✓ | ✓ | Low risk |
| ERSPC Switzerland | ✗ | ✓ | ✓ | ✓ | ✓ | ✓ | ✓ | Low risk |
| PLCO | ✓ | ✗ | ✓ | ✓ | ✓ | ✓ | ✓ | Some concerns |
| **Major biopsy complications** | | | | | | | | |
| PLCO | ✓ | ✗ | ✓ | ✓ | ✓ | ✗ | ✓ | High risk |
| **Any infection after biopsy, 1st round** | | | | | | | | |
| ERSPC Netherlands | ✓ | ✓ | ? | ✓ | ✓ | ✗ | ✓ | Some concerns |
| Göteborg-2 | ✓ | ✓ | ✓ | ✓ | ✓ | ✓ | ✓ | Low risk |
| **Any infection after biopsy, ≥2 rounds** | | | | | | | | |
| ERSPC Netherlands | ✓ | ✓ | ✓ | ✓ | ✓ | ✗ | ✓ | Some concerns |
| **Major bleeding (≥1 rounds)** | | | | | | | | |
| PLCO | ✓ | ✗ | ✓ | ✓ | ✓ | ✗ | ✓ | High risk |
| **Hospitalization after biopsy (≥1 rounds)** | | | | | | | | |
| ERSPC Netherlands | ✓ | ✓ | ✓ | ✓ | ✓ | ✓ | ✓ | Low risk |
| Göteborg-2 | ✓ | ✓ | ✓ | ✓ | ✓ | ✓ | ✓ | Low risk |

**Note: See Appendix 1 for questions and explanation of assessment tool. ✓= Yes; ✗= No; ?**

**= insufficient information to assess.**

#### Per protocol data

| Study | Q2 | Q3 | Q6 | Q7 | Q9 | Q11 | Q13 | Overall |
|---|---|---|---|---|---|---|---|---|
| **False positives** | | | | | | | | |
| ERSPC Belgium | ✗ | ✗ | ✓ | ✓ | ✗ | ✗ | ✓ | High risk |
| ERSPC Finland | ✓ | ✓ | ✓ | ✓ | ✓ | ✗ | ✓ | Some concerns |
| ERSPC France | ✓ | ✗ | ✓ | ✓ | ✓ | ✗ | ✓ | High risk |
| ERSPC Italy | ✗ | ✗ | ✓ | ✓ | ✓ | ✗ | ✓ | Some concerns |
| ERSPC Netherlands | ✓ | ✓ | ✓ | ✓ | ✓ | ✗ | ✓ | Some concerns |

| | | | | | | | | |
|---|---|---|---|---|---|---|---|---|
| ERSPC Spain | ✓ | ✓ | ✓ | ✓ | ✓ | ✗ | ✓ | Some concerns |
| ERSPC Sweden | ✓ | ✓ | ✓ | ✓ | ✓ | ✗ | ✓ | Some concerns |
| ERSPC Switzerland | ✗ | ✓ | ✓ | ✓ | ✓ | ✗ | ✓ | Some concerns |
| PLCO | ✓ | ✗ | ✓ | ✓ | ✓ | ✓ | ✓ | Some concerns |
| SABOR | ✗ | ✗ | ✓ | ✓ | ✗ | ✓ | ✓ | High risk |
| **Any complications** | | | | | | | | |
| ERSPC Netherlands | ✓ | ✓ | ? | ✓ | ✓ | ✓ | ✓ | Low risk |
| PROBE | ✓ | ✗ | ? | ✓ | ✓ | ✓ | ✓ | Some concerns |
| **Major complications** | | | | | | | | |
| PLCO | ✓ | ✗ | ✓ | ✓ | ✓ | ✗ | ✓ | High risk |
| PROBE | ✓ | ✗ | ? | ✓ | ✓ | ✓ | ✓ | Some concerns |
| Sung 2021 | ✓ | ✓ | ? | ✓ | ? | ? | ? | Some concerns |
| **Biopsy mortality, 1 round** | | | | | | | | |
| ERSPC Belgium | ✗ | ✗ | ✓ | ✓ | ✗ | ✓ | ✓ | Some concerns |
| ERSPC Finland | ✓ | ✓ | ✓ | ✓ | ✓ | ✓ | ✓ | Low risk |
| ERSPC Italy | ✗ | ✗ | ✓ | ✓ | ✓ | ✓ | ✓ | Some concerns |
| ERSPC Netherlands | ✓ | ✓ | ✓ | ✓ | ✓ | ✓ | ✓ | Low risk |
| ERSPC Spain | ✓ | ✓ | ✓ | ✓ | ✓ | ✓ | ✓ | Low risk |
| ERSPC Sweden | ✓ | ✓ | ✓ | ✓ | ✗ | ✓ | ✓ | Low risk |
| ERSPC Switzerland | ✗ | ✓ | ✓ | ✓ | ✓ | ✓ | ✓ | Low risk |
| Göteborg-2 | ✓ | ✓ | ✓ | ✓ | ✓ | ✓ | ✓ | Low risk |
| PLCO | ✓ | ✗ | ✓ | ✓ | ✓ | ✓ | ✓ | Some concerns |
| PROBE | ✓ | ✗ | ✓ | ✓ | ✓ | ✓ | ✓ | Some concerns |
| **Biopsy mortality, ≥2 rounds** | | | | | | | | |
| ERSPC Belgium | ✗ | ✗ | ✓ | ✓ | ✗ | ✓ | ✓ | Some concerns |
| ERSPC Finland | ✓ | ✓ | ✓ | ✓ | ✓ | ✓ | ✓ | Low risk |
| ERSPC Italy | ✗ | ✗ | ✓ | ✓ | ✓ | ✓ | ✓ | Some concerns |
| ERSPC Netherlands | ✓ | ✓ | ✓ | ✓ | ✓ | ✓ | ✓ | Low risk |
| ERSPC Spain | ✓ | ✗ | ✓ | ✓ | ✓ | ✓ | ✓ | Some concerns |
| ERSPC Sweden | ✓ | ✓ | ✓ | ✓ | ✗ | ✓ | ✓ | Low risk |
| ERSPC Switzerland | ✗ | ✓ | ✓ | ✓ | ✓ | ✓ | ✓ | Low risk |
| PLCO | ✓ | ✗ | ✓ | ✓ | ✓ | ✓ | ✓ | Some concerns |

| | | | | | | | | |
|---|---|---|---|---|---|---|---|---|
| **Biopsy hospitalization** | | | | | | | | |
| ERSPC Netherlands | ✓ | ✓ | ✓ | ✓ | ✓ | ✓ | ✓ | Low risk |
| Göteborg-2 | ✓ | ✓ | ✓ | ✓ | ✓ | ✓ | ✓ | Low risk |
| PROBE | ✓ | ✗ | ✓ | ✓ | ✓ | ✓ | ✓ | Some concerns |
| **Any infection, 1 round** | | | | | | | | |
| ERSPC Netherlands | ✓ | ✓ | ? | ✓ | ✓ | ✗ | ✓ | Some concerns |
| Göteborg-2 | ✓ | ✓ | ✓ | ✓ | ✓ | ✓ | ✓ | Low risk |
| PROBE | ✓ | ✗ | ✓ | ✓ | ✓ | ✗ | ✓ | High risk |
| **Any infection ≥2 rounds** | | | | | | | | |
| ERSPC Netherlands | ✓ | ✓ | ✓ | ✓ | ✓ | ✗ | ✓ | Some concerns |
| **Major infection, 1 round** | | | | | | | | |
| PROBE | ✓ | ✗ | ✓ | ✓ | ✓ | ✓ | ✓ | Some concerns |
| Sung 2021 | ✓ | ✓ | ✓ | ✓ | ? | ? | ? | Some concerns |
| **Major infection, ≥2 rounds** | | | | | | | | |
| PLCO | ✓ | ✗ | ✓ | ✗ | ✓ | ✗ | ✓ | High risk |
| **Major bleeding** | | | | | | | | |
| PLCO | ✓ | ✗ | ✓ | ✗ | ✓ | ✗ | ✓ | High risk |
| PROBE | ✓ | ✗ | ✓ | ✓ | ✓ | ✓ | ✓ | Some concerns |
| **Any bleeding** | | | | | | | | |
| ERSPC Netherlands | ✓ | ✓ | ✓ | ✓ | ✓ | ✓ | ✓ | Low risk |
| PROBE | ✓ | ✗ | ? | ✓ | ✓ | ✓ | ✓ | Some concerns |

**Note: See Appendix 1 for questions and explanation of assessment tool. ✓= Yes; ✗= No; ?**

**= insufficient information to assess.**

**Screening for prostate cancer using PSA with and without MRI: systematic reviews with meta-analysis**
Jennifer Pillay, Lindsay Gaudet, Sholeh Rahman, Roland Grad, Guylène Thériault, Philipp Dahm, Keith J Todd, Gail McCartney, Brett Thombs, Sabrina Saba, Lisa Hartling

#### Additional KQ1 Figures. Note: also see manuscript Figure 2

##### 1. Prostate-cancer mortality

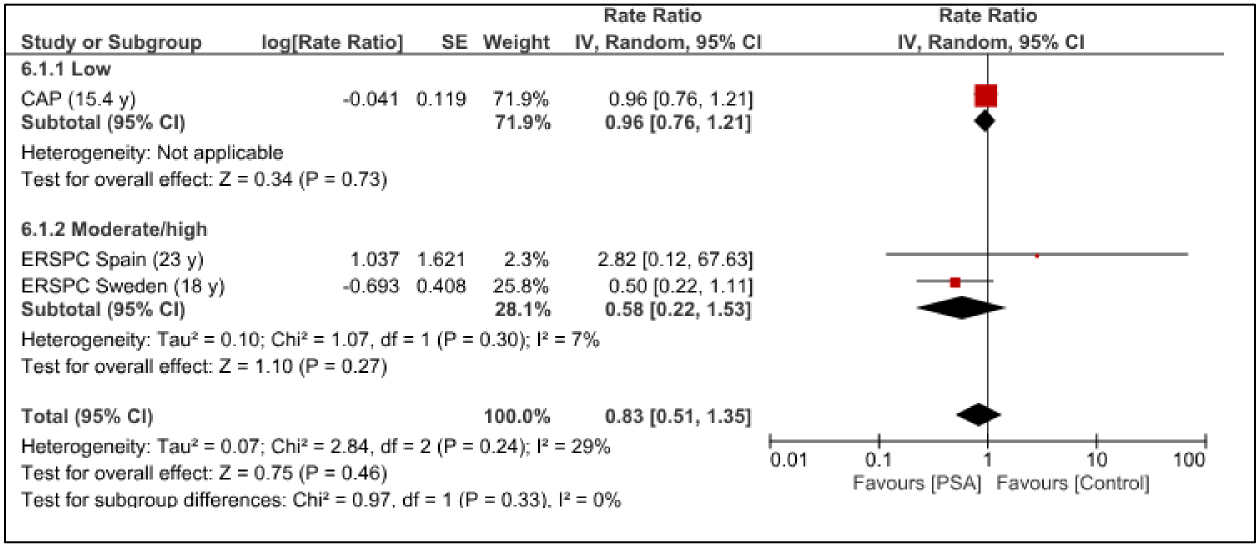


Prostate-cancer mortality by screening intensity for screening starting at age 50-54 years

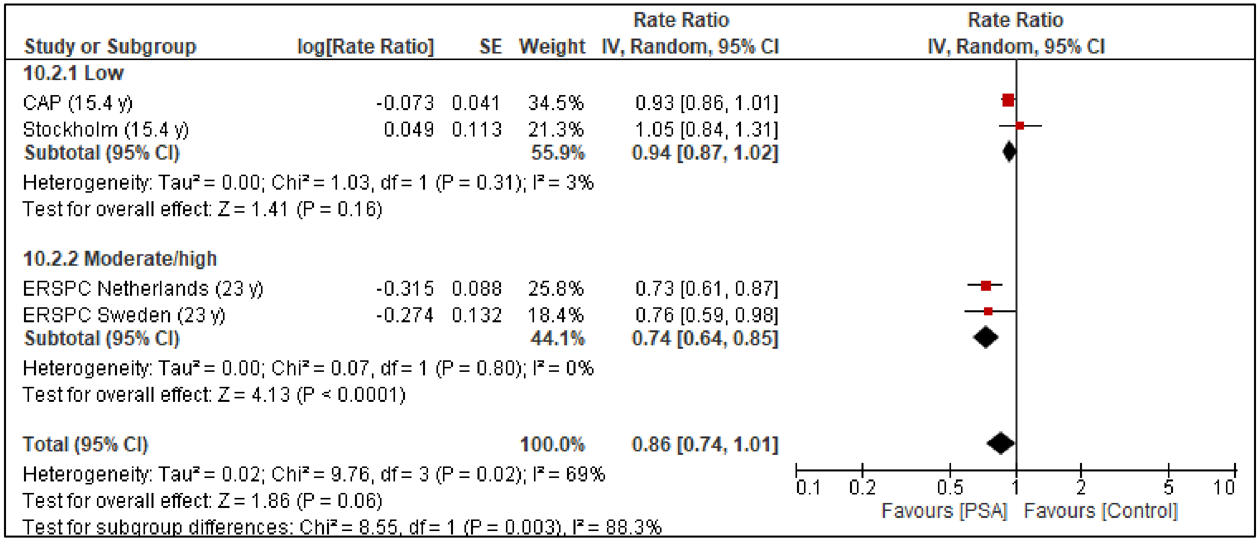


Prostate-cancer mortality by screening intensity for screening starting at age 55-69 years

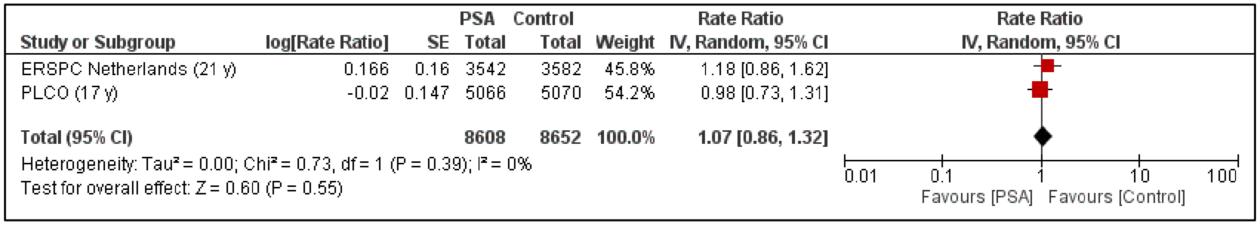


Prostate-cancer mortality for screening starting at age 70-74 years (low-intensity screening)

#### 2. All-cause mortality

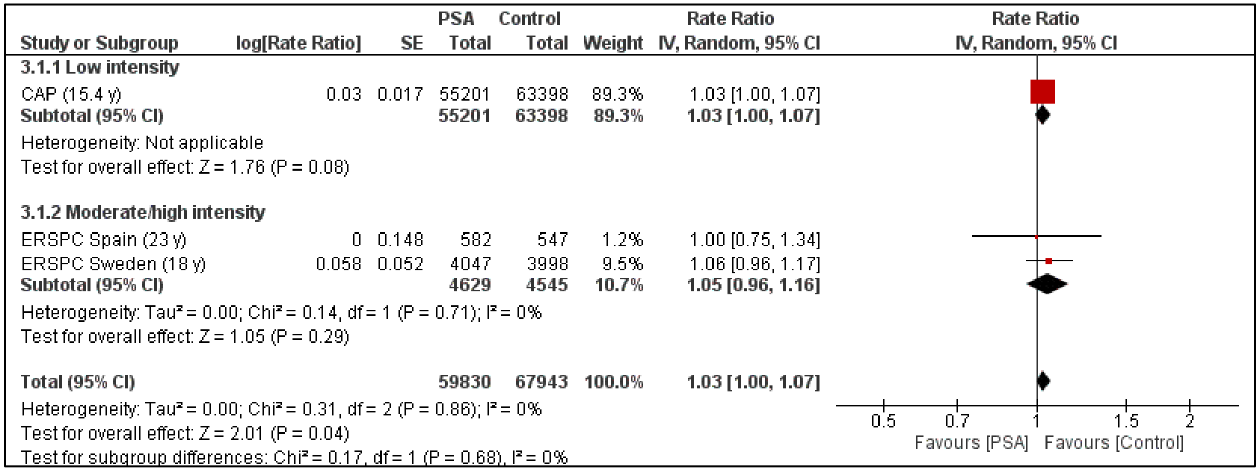


All-cause mortality by screening intensity for screening starting at age 50-54 years

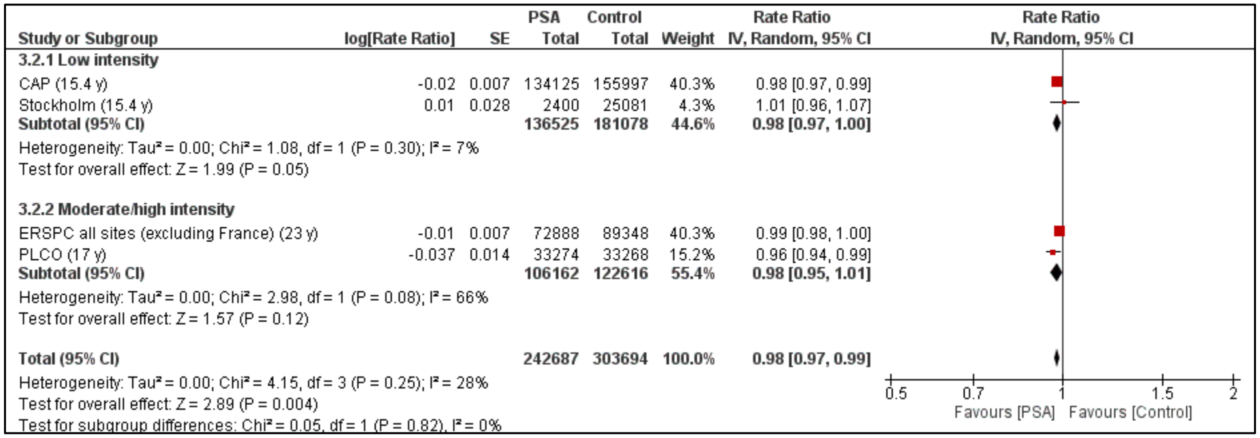


All-cause mortality by screening intensity for screening starting at age 55-69 years

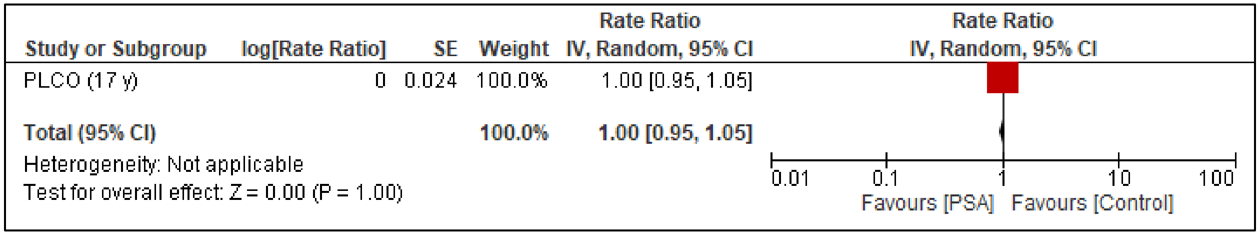


All-cause mortality for screening starting at age 70-74 years (low-intensity screening)

#### 3. Incidence of metastatic prostate cancer

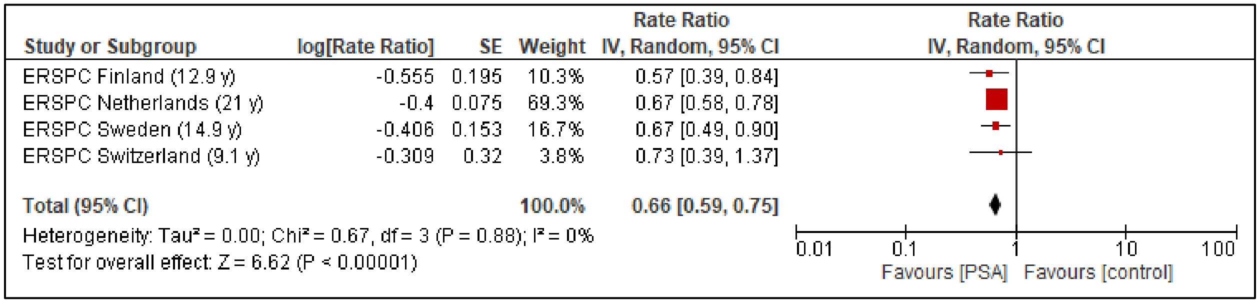


Incidence of metastatic prostate cancer (through follow-up) for screening starting at age 55-69 years (moderate/high-intensity screening)

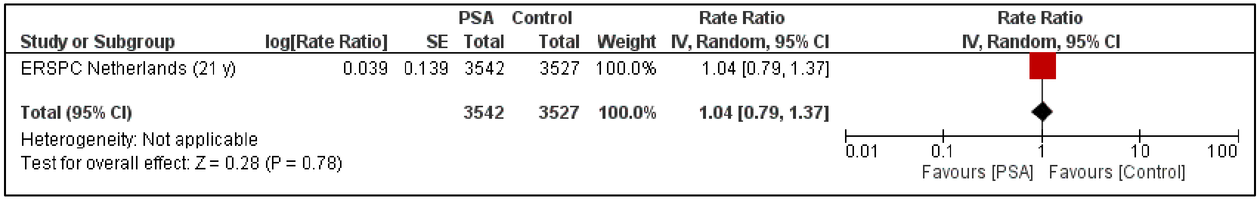


Incidence of metastatic prostate cancer (through follow-up) for screening starting at age 70-74 years (low-intensity screening)

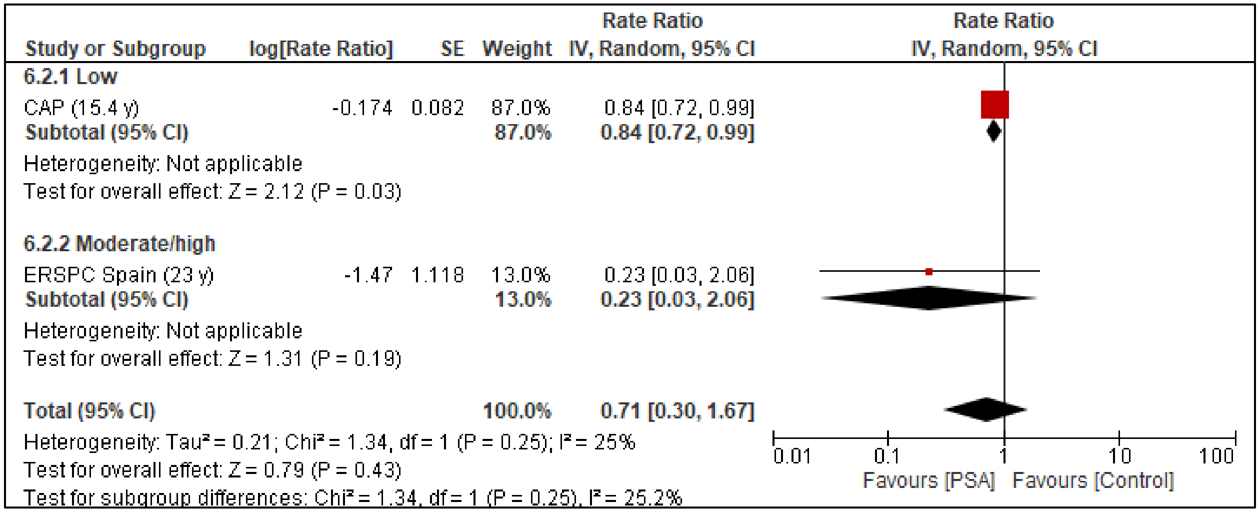

Incidence of metastatic prostate cancer, i.e., advanced cancer (at diagnosis) by screening intensity for screening starting at age 50-54 years

| Study or Subgroup | log[Rate Ratio] | SE | Weight | Rate Ratio IV, Random, 95% CI |
|---|---|---|---|---|
| **7.3.1 Low** | | | | |
| CAP (15.4 y) | -0.174 | 0.039 | 20.4% | 0.84 [0.78, 0.91] |
| **Subtotal (95% CI)** | | | **20.4%** | **0.84 [0.78, 0.91]** |
| Heterogeneity: Not applicable | | | | |
| Test for overall effect: Z = 4.46 (P < 0.00001) | | | | |
| **7.3.2 Moderate/high** | | | | |
| ERSPC Netherlands (21 y) | -0.799 | 0.11 | 17.3% | 0.45 [0.36, 0.56] |
| ERSPC Switzerland (23 y) | -0.616 | 0.231 | 10.9% | 0.54 [0.34, 0.85] |
| ERSPC Italy (21 y) | -0.315 | 0.366 | 6.3% | 0.73 [0.36, 1.50] |
| ERSPC Sweden (23 y) | -0.301 | 0.145 | 15.3% | 0.74 [0.56, 0.98] |
| ERSPC Finland (23 y) | -0.274 | 0.077 | 19.0% | 0.76 [0.65, 0.88] |
| ERSPC Belgium (23 y) | 0.285 | 0.231 | 10.9% | 1.33 [0.85, 2.09] |
| **Subtotal (95% CI)** | | | **79.6%** | **0.70 [0.53, 0.93]** |
| Heterogeneity: Tau² = 0.09; Chi² = 26.01, df = 5 (P < 0.0001); I² = 81% | | | | |
| Test for overall effect: Z = 2.47 (P = 0.01) | | | | |
| **Total (95% CI)** | | | **100.0%** | **0.72 [0.58, 0.89]** |
| Heterogeneity: Tau² = 0.06; Chi² = 36.83, df = 6 (P < 0.00001); I² = 84% | | | | |
| Test for overall effect: Z = 2.97 (P = 0.003) | | | | |
| Test for subgroup differences: Chi² = 1.52, df = 1 (P = 0.22), I² = 34.1% | | | | |

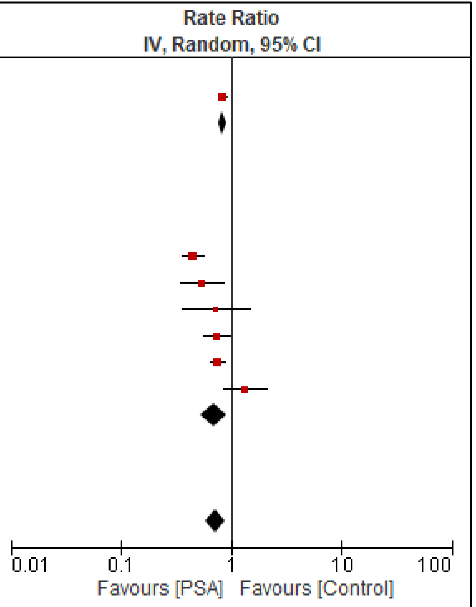


Incidence of metastatic prostate cancer, i.e., advanced cancer (at diagnosis) by screening intensity for screening starting at age 55-69 years

| Study or Subgroup | log[Rate Ratio] | SE | PSA Total | Control Total | Weight | Rate Ratio IV, Random, 95% CI |
|---|---|---|---|---|---|---|
| ERSPC Netherlands (21 y) | -0.4 | 0.197 | 3542 | 3527 | 61.7% | 0.67 [0.46, 0.99] |
| PLCO (15 y) | -0.548 | 0.25 | 5066 | 5070 | 38.3% | 0.58 [0.35, 0.94] |
| **Total (95% CI)** | | | **8608** | **8597** | **100.0%** | **0.63 [0.47, 0.86]** |
| Heterogeneity: Tau² = 0.00; Chi² = 0.22, df = 1 (P = 0.64); I² = 0% | | | | | | |
| Test for overall effect: Z = 2.95 (P = 0.003) | | | | | | |

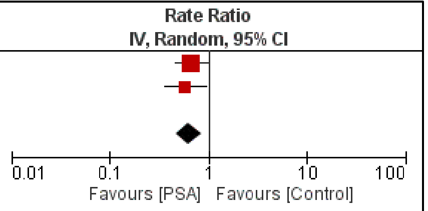


Incidence of metastatic prostate cancer (at diagnosis) for screening starting at age 70-74 years (low-intensity screening)

#### 4. Overdiagnosis (i.e., cancer incidence over long-term follow-up)

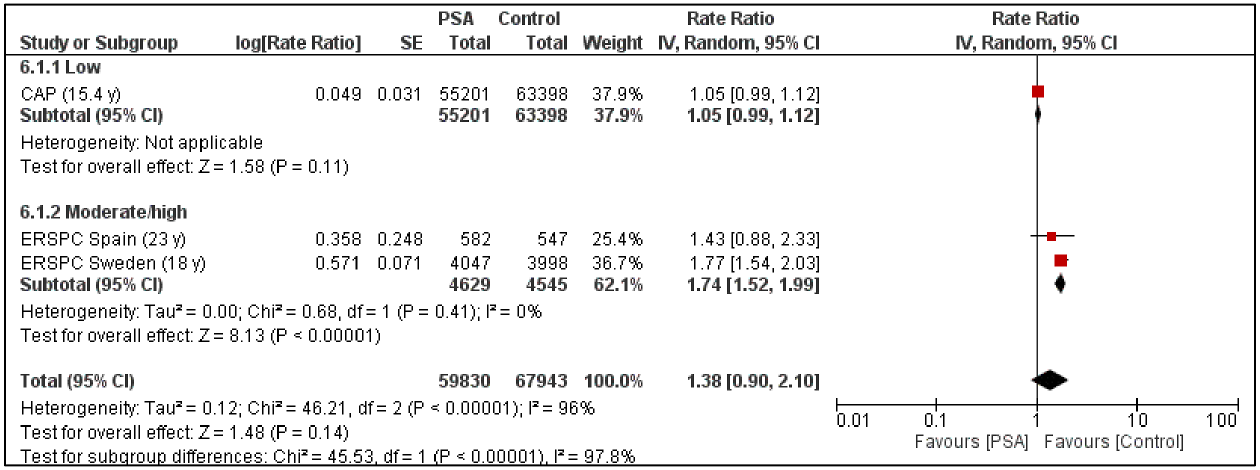


Overdiagnosis by screening intensity for screening starting at age 50-54 years

| Study or Subgroup | log[Rate Ratio] | SE | Weight | Rate Ratio IV, Random, 95% CI |
|---|---|---|---|---|
| **10.2.1 Low** | | | | |
| CAP (15.4 y) | 0.068 | 0.014 | 20.7% | 1.07 [1.04, 1.10] |
| Stockholm (15.4 y) | 0.113 | 0.062 | 19.4% | 1.12 [0.99, 1.26] |
| **Subtotal (95% CI)** | | | **40.1%** | **1.07 [1.04, 1.10]** |
| Heterogeneity: Tau² = 0.00; Chi² = 0.50, df = 1 (P = 0.48); I² = 0% | | | | |
| Test for overall effect: Z = 5.14 (P < 0.00001) | | | | |
| **10.2.2 Moderate/high** | | | | |
| ERSPC Sweden (23 y) | 0.278 | 0.048 | 19.9% | 1.32 [1.20, 1.45] |
| ERSPC Switzerland (22 y) | 0.419 | 0.058 | 19.5% | 1.52 [1.36, 1.70] |
| ERSPC Netherlands (23 y) | 0.476 | 0.03 | 20.4% | 1.61 [1.52, 1.71] |
| **Subtotal (95% CI)** | | | **59.9%** | **1.48 [1.31, 1.68]** |
| Heterogeneity: Tau² = 0.01; Chi² = 12.24, df = 2 (P = 0.002); I² = 84% | | | | |
| Test for overall effect: Z = 6.20 (P < 0.00001) | | | | |
| **Total (95% CI)** | | | **100.0%** | **1.31 [1.07, 1.61]** |
| Heterogeneity: Tau² = 0.05; Chi² = 180.69, df = 4 (P < 0.00001); I² = 98% | | | | |
| Test for overall effect: Z = 2.59 (P = 0.010) | | | | |
| Test for subgroup differences: Chi² = 24.79, df = 1 (P < 0.00001), I² = 96.0% | | | | |

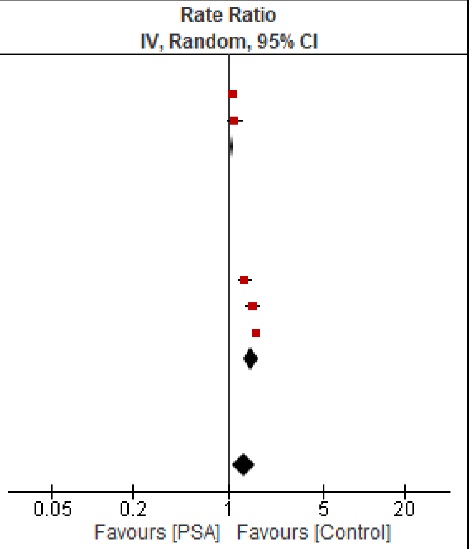


Overdiagnosis by screening intensity for screening starting at age 55-69 years

| Study or Subgroup | log[Rate Ratio] | SE | PSA Total | Control Total | Weight | Rate Ratio IV, Random, 95% CI |
|---|---|---|---|---|---|---|
| ERSPC Netherlands (21 y) | 0.399 | 0.073 | 3542 | 3527 | 48.7% | 1.49 [1.29, 1.72] |
| PLCO (15 y) | 0.082 | 0.053 | 5066 | 5070 | 51.3% | 1.09 [0.98, 1.20] |
| **Total (95% CI)** | | | **8608** | **8597** | **100.0%** | **1.27 [0.93, 1.73]** |
| Heterogeneity: Tau² = 0.05; Chi² = 12.35, df = 1 (P = 0.0004); I² = 92% | | | | | | |
| Test for overall effect: Z = 1.49 (P = 0.14) | | | | | | |

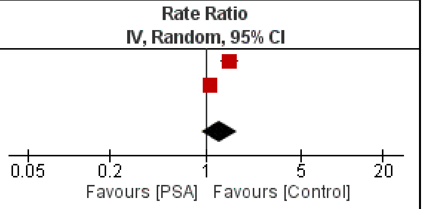


Overdiagnosis for screening starting at age 70-74 years (low-intensity screening)

#### Subgroup analysis findings from comparative data

##### 1. Prostate-cancer mortality

| | N trials | n | RaR (95% CI) | Mean control event rate(s) at 20 y unless stated | $I^2$ | Absolute effect with screening [95% CI] per 1,000 | Absolute difference (95% CI) per 1,000 |
|---|---|---|---|---|---|---|---|
| **Prostate cancer morality** | | | | | | | |
| All studies | | | | | | | |
| 50-54y | 3 | 127,773 | 0.83 [0.51, 1.35] | 5 per 1000 | 29% | 4.15 [2.55, 6.75] | -0.85 [-2.45, 1.75] |
| 55-69y | 11 | 623,123 | 0.91 [0.85, 0.98] | 55: 8 per 1000<br><br>55-69: 15 per 1000 | 19% | 55: 7.28 [6.80, 7.84]<br><br>55-69: 13.65 [12.75, 14.70] | 55: -0.72 [-1.20, -0.16]<br><br>55-69: -1.35 [-2.25, -0.30] |
| 70-74y | 2 | 17,260 | 1.07 [0.86, 1.32] | 21 per 1000 (15y) | 0% | 22.47 [18.06, 27.72] | 1.47 [-2.94, 6.72] |
| 50-54y: By intensity (low intensity [one screen] vs. moderate intensity [2-4 rounds [offered to majority] &/or ≥3 y screening interval] vs. high intensity [5+ rounds &/or annual/biennial screening]); p=0.24 (p=0.33 for low vs. moderate/high) | | | | | | | |
| Low intensity | 1 | 118,599 | 0.96 [0.76, 1.21] | 5 per 1000 | NA | 4.80 [3.80, 6.05] | -0.20 [-1.20, 1.05] |
| Moderate intensity | 1 | 1,129 | 2.82 [0.12, 67.63] | 5 per 1000 | NA | 14.10 [0.60, 338.15] | 9.10 [-4.40, 333.15] |
| High intensity | 1 | 8,045 | 0.50 [0.22, 1.11] | 5 per 1000 | NA | 2.50 [1.10, 5.55] | -2.50 [-3.90, 0.55] |
| Moderate/high intensity | 2 | 9,174 | 0.58 [0.22, 1.53] | 5 per 1000 | 7% | 2.90 [1.10, 7.65] | -2.10 [-3.90, 2.65] |
| 55-69 y: By intensity (e.g., low intensity [one screen] vs. moderate intensity [2-4 rounds [offered to majority] &/or 3+ year screening interval] vs. high intensity [5+ rounds &/or annual/biennial screening]); p=0.61 (p=0.34 for low vs. moderate/high) | | | | | | | |
| Low intensity | 2 | 317,603 | 0.94 [0.87, 1.02] | 55: 8 per 1000<br><br>55-69: 15 per 1000 | 3% | 55: 7.52 [6.96, 8.16]<br><br>55-69: 14.10 [13.05, 15.30] | 55: -0.48 [-1.04, 0.16]<br><br>55-69: -0.90 [-1.95, 0.30] |
| Moderate intensity | 7 | 227,124 | 0.91 [0.80, 1.03] | 55: 8 per 1000<br><br>55-69: 15 per 1000 | 29% | 55: 7.28 [6.40, 8.24]<br><br>55-69: 13.65 [12.00, 15.45] | 55: -0.72 [-1.60, 0.24]<br><br>55-69: -1.35 [-3.00, 0.45] |
| High intensity | 2 | 78,396 | 0.86 [0.72, 1.03] | 55: 8 per 1000<br><br>55-69: 15 per 1000 | 31% | 55: 6.88 [5.76, 8.24]<br><br>55-69: 12.90 [10.80, 15.45] | 55: -1.12 [-2.24, 0.24]<br><br>55-69: -2.10 [-4.20, 0.45] |
| Moderate/high intensity | 9 | 305,520 | 0.89 [0.81, 0.98] | 55: 8 per 1000<br><br>55-69: 15 per 1000 | 20% | 55: 7.12 [6.48, 7.84]<br><br>55-69: 13.35 [12.15, 14.70] | 55: -0.88 [-1.52, -0.16]<br><br>55-69: -1.65 [-2.85, -0.30] |
| 50-54y: high vs lower ROB; p=0.30 **(among moderate/high intensity interventions)** | | | | | | | |
| High ROB | 1 | 1,129 | 2.82 [0.12, 67.63] | 5 per 1000 | NA | 14.10 [0.60, 338.15] | 9.10 [-4.40, 333.15] |

| | N trials | n | RaR (95% CI) | Mean control event rate(s) at 20 y unless stated | $I^2$ | Absolute effect with screening [95% CI] per 1,000 | Absolute difference (95% CI) per 1,000 |
|---|---|---|---|---|---|---|---|
| Lower ROB | 1 | 8,045 | 0.50 [0.22, 1.11] | 5 per 1000 | NA | 2.50 [1.10, 5.55] | -2.50 [-3.90, 0.55] |
| 55-69y: high vs lower ROB; p=0.31 **(among low intensity interventions)** | | | | | | | |
| High ROB | 1 | 290,122 | 0.93 [0.86, 1.01] | 55: 8 per 1000<br><br>55-69: 15 per 1000 | NA | 55: 7.44 [6.88, 8.08]<br><br>55-69: 13.95 [12.90, 15.15] | 55: -0.56 [-1.12, 0.08]<br><br>55-69: -1.05 [-2.10, 0.15] |
| Lower ROB | 1 | 27,481 | 1.05 [0.84, 1.31] | 55: 8 per 1000<br><br>55-69: 15 per 1000 | NA | 55: 8.40 [6.72, 10.48]<br><br>55-69: 15.75 [12.60, 19.65] | 55: 0.40 [-1.28, 2.48]<br><br>55-69: 0.75 [-2.40, 4.65] |
| 55-69y: high vs lower ROB; **p=0.003 (among moderate/high intensity interventions)** | | | | | | | |
| High ROB | 7 | 258,521 | 0.95 [0.87, 1.04] | 55: 8 per 1000<br><br>55-69: 15 per 1000 | 0% | 55: 7.60 [6.96, 8.32]<br><br>55-69: 14.25 [13.05, 15.60] | 55: -0.40 [-1.04, 0.32]<br><br>55-69: -0.75 [-1.95, 0.60] |
| Lower ROB | 2 | 46,999 | 0.74 [0.64, 0.85] | 55: 8 per 1000<br><br>55-69: 15 per 1000 | 0% | 55: 5.92 [5.12, 6.80]<br><br>55-69: 11.10 [9.60, 12.75] | 55: -2.08 [-2.88, -1.20]<br><br>55-69: -3.90 [-5.40, -2.25] |
| 70-74y: high vs lower ROB; p=0.39 | | | | | | | |
| High ROB | 1 | 10,136 | 0.98 [0.73, 1.31] | 21 per 1000 (15y) | NA | 20.58 [15.33, 27.51] | -0.42 [-5.67, 6.51] |
| Lower ROB | 1 | 7,124 | 1.18 [0.86, 1.62] | 21 per 1000 (15y) | NA | 24.78 [18.06, 34.02] | 3.78 [-2.94, 13.02] |
| 50-54y: 50-54y vs. broader age; p=0.30 **(among moderate/high intensity interventions)** | | | | | | | |
| 50-54y | 1 | 8,045 | 0.50 [0.22, 1.11] | 5 per 1000 | NA | 2.50 [1.10, 5.55] | -2.50 [-3.90, 0.55] |
| 45-54y | 1 | 1,129 | 2.83 [0.12, 67.63] | 5 per 1000 | NA | 14.10 [0.60, 338.15] | 9.10 [-4.40, 333.15] |
| 55-69y: PSA +/- DRE vs. PSA OR DRE; p=0.31 **(among low intensity interventions)** | | | | | | | |
| PSA +/- DRE | 1 | 290,122 | 0.93 [0.86, 1.01] | 55: 8 per 1000<br><br>55-69: 15 per 1000 | NA | 55: 7.44 [6.88, 8.08]<br><br>55-69: 13.95 [12.90, 15.15] | 55: -0.56 [-1.12, 0.08]<br><br>55-69: -1.05 [-2.10, 0.15] |
| PSA OR DRE | 1 | 27,481 | 1.05 [0.84, 1.31] | 55: 8 per 1000<br><br>55-69: 15 per 1000 | NA | 55: 8.40 [6.72, 10.48]<br><br>55-69: 15.75 [12.60, 19.65] | 55: 0.40 [-1.28, 2.48]<br><br>55-69: 0.75 [-2.40, 4.65] |
| 55-69y: PSA +/- DRE vs. PSA OR DRE; p=0.53 **(among moderate/high intensity interventions)** | | | | | | | |
| PSA +/- DRE | 7 | 230,416 | 0.88 [0.78, 1.00] | 55: 8 per 1000<br><br>55-69: 15 per 1000 | 35% | 55: 7.52 [6.4, 8.8]<br><br>55-69: 13.20 [11.70, 15.15] | 55: -0.48 [-1.6, 0.8]<br><br>55-69: -1.80 [-3.30, 0.15] |
| PSA OR DRE | 2 | 75,104 | 0.94 [0.80, 1.10] | 55: 8 per 1000<br><br>55-69: 15 per 1000 | 0% | 55: 7.04 [6.24, 8.08]<br><br>55-69: 14.10 [12.00, 16.50] | 55: -0.96 [ -1.76, 0.08]<br><br>55-69: -0.90 [-3.00, 1.50] |
| 70-74y: PSA +/- DRE vs. PSA OR DRE; p=0.39 | | | | | | | |

| | N trials | n | RaR (95% CI) | Mean control event rate(s) at 20 y unless stated | $I^2$ | Absolute effect with screening [95% CI] per 1,000 | Absolute difference (95% CI) per 1,000 |
|---|---|---|---|---|---|---|---|
| PSA +/- DRE | 1 | 7,124 | 1.18 [0.86, 1.62] | 21 per 1000 (15y) | NA | 24.78 [18.06, 34.02] | 3.78 [-2.94, 13.02] |
| PSA OR DRE | 1 | 10,136 | 0.98 [0.73, 1.31] | 21 per 1000 (15y) | NA | 20.58 [15.33, 27.51] | -0.42 [-5.67, 6.51] |

#### 2. All-cause mortality

| | N trials | n | RaR (95% CI) | Mean control event rate(s) at 20 y unless stated | $I^2$ | Absolute effect with screening [95% CI] per 1,000 | Absolute difference (95% CI) per 1,000 |
|---|---|---|---|---|---|---|---|
| **All-cause morality** | | | | | | | |
| All studies | | | | | | | |
| 50-54y | 3 | 127,773 | 1.03 [1.00, 1.07] | 164 per 1000 | 0% | 168.92 [164.00, 175.48] | 4.92 [0.00, 11.48] |
| 55-69y | 10 | 546,381 | 0.98 [0.97, 0.99] | 55: 249 per 1000<br><br>55-69: 423 per 1000 | 28% | 55: 244.02 [241.53, 246.51]<br><br>55-69: 414.54 [410.3, 418.77] | 55: -4.98 [-7.47, -2.49]<br><br>55-69: -8.46 [-12.69, -4.23] |
| 70-74y | 1 | 10,136 | 1.00 [0.95, 1.05] | 565 per 1000 [15y] | NA | 565.00 [536.75, 593.25] [15y] | 0.00 [-28.25, 28.25] [15y] |
| 50-54y: By intensity (low intensity [one screen] vs. moderate intensity [2-4 rounds [offered to majority] &/or ≥3 y screening interval] vs. high intensity [5+ rounds &/or annual/biennial screening]); p=0.86 (p=0.68 low vs. moderate/high intensity) | | | | | | | |
| Low intensity | 1 | 118,599 | 1.03 [1.00, 1.07] | 164 per 1000 | NA | 168.92 [164.00, 175.48] | 4.92 [0.00, 11.48] |
| Moderate intensity | 1 | 1,129 | 1.00 [0.75, 1.34] | 164 per 1000 | NA | 164.00 [123.00, 219.76] | 0.00 [-41.00, 55.76] |
| High intensity | 1 | 8,045 | 1.06 [0.96, 1.17] | 164 per 1000 | NA | 173.84 [157.44, 191.88] | 9.84 [-6.56, 27.88] |
| Moderate/high intensity | 2 | 9,174 | 1.05 [0.96, 1.16] | 164 per 1000 | 0% | 172.20 [157.44, 190.24] | 8.20 [-6.56, 26.24] |
| 55-69 y: By intensity (e.g., low intensity [one screen] vs. moderate intensity [2-4 rounds [offered to majority] &/or 3+ year screening interval] vs. high intensity [5+ rounds &/or annual/biennial screening]); p=0.22 (p=0.82 for low vs. moderate/high intensity) | | | | | | | |
| Low intensity | 2 | 317,603 | 0.98 [0.97, 1.00] | 55: 249 per 1000<br><br>55-69: 423 per 1000 | 7% | 55: 244.02 [241.53, 249.00]<br><br>55-69: 414.54 [410.31, 423.00] | 55: -4.98 [-7.47, 0.00]<br><br>55-69: -8.46 [-12.69, 0.00] |
| Moderate intensity | 7 | 162,236 | 0.99 [0.98, 1.00] | 55: 249 per 1000<br><br>55-69: 423 per 1000 | NA (data across sites) | 55: 246.51 [244.02, 249.00]<br><br>55-69: 418.77 [414.54, 423.00] | 55: -2.49 [-4.98, 0.00]<br><br>55-69: -4.23 [-8.46, 0.00] |

| | N trials | n | RaR (95% CI) | Mean control event rate(s) at 20 y unless stated | $I^2$ | Absolute effect with screening [95% CI] per 1,000 | Absolute difference (95% CI) per 1,000 |
|---|---|---|---|---|---|---|---|
| High intensity | 1 | 66,542 | 0.96 [0.94, 0.99] | 55: 249 per 1000<br><br>55-69: 423 per 1000 | NA | 55: 239.04 [234.06, 246.51]<br><br>55-69: 406.08 [397.62, 418.77] | 55: -9.96 [-14.94, -2.49]<br><br>55-69: -16.92 [-25.38, -4.23] |
| Moderate/high intensity | 8 | 228,778 | 0.98 [0.95, 1.01] | 55: 249 per 1000<br><br>55-69: 423 per 1000 | 66% | 55: 244.02 [236.55, 251.49]<br><br>55-69: 414.54 [401.85, 427.23] | 55: -4.98 [-12.45, 2.49]<br><br>55-69: -8.46 [-21.15, 4.23] |
| 50-54y: high vs lower ROB; p=0.71 (among moderate/high intensity interventions) | | | | | | | |
| High ROB | 1 | 1,129 | 1.00 [0.75, 1.34] | 164 per 1000 | NA | 164.00 [123.00, 219.76] | 0.00 [-41.00, 55.76] |
| Lower ROB | 1 | 8,045 | 1.06 [0.96, 1.17] | 164 per 1000 | NA | 173.84 [157.44, 191.88] | 9.84 [-6.56, 27.88] |
| 55-69y: high vs lower ROB; p=NA (among low intensity interventions) | | | | | | | |
| High ROB | 0 | NA | NA | NA | NA | NA | NA |
| Lower ROB | All studies | | | | | | |
| 55-69y: high vs lower ROB; p=NA (among moderate/high intensity interventions) | | | | | | | |
| High ROB | 0 | NA | NA | NA | NA | NA | NA |
| Lower ROB | All studies | | | | | | |
| 70-74y: high vs lower ROB; p=NA | | | | | | | |
| High ROB | 0 | NA | NA | NA | NA | NA | NA |
| Lower ROB | All studies | | | | | | |
| 50-54y: 50-54y vs. broader age; p=0.71 (among moderate/high intensity interventions) | | | | | | | |
| 50-54y | 1 | 8,045 | 1.06 [0.96, 1.17] | 164 per 1000 | NA | 173.84 [157.44, 191.88] | 9.84 [-6.56, 27.88] |
| 45-54y | 1 | 1,129 | 1.00 [0.75, 1.34] | 164 per 1000 | NA | 164.00 [123.00, 219.76] | 0.00 [-41.00, 55.76] |
| 55-69y: 55-69 vs broader age p=NA (among low intensity interventions) | | | | | | | |
| 55-69y | All studies | | | | | | |
| Broader age range | 0 | NA | NA | NA | NA | NA | NA |
| 55-69y: 55-69 vs broader age p=NA (among moderate/high intensity interventions) | | | | | | | |
| 55-69y | All studies | | | | | | |
| Broader age range | 0 | NA | NA | NA | NA | NA | NA |
| 50-54y: PSA +/- DRE vs. PSA OR DRE (among moderate/high intensity interventions) | | | | | | | |
| PSA +/- DRE | All studies | | | | | | |
| PSA OR DRE | 0 | NA | NA | NA | NA | NA | NA |
| 55-69y: PSA +/- DRE vs. PSA OR DRE; p=0.30 **(among low intensity interventions)** | | | | | | | |
| PSA +/- DRE | 1 | 290,122 | 0.98 [0.97, 0.99] | 55: 249 per 1000<br><br>55-69: 423 per 1000 | NA | 55: 244.02 [241.53, 246.51]<br><br>55-69: 414.54 [410.31, 418.77] | 55: -4.98 [-7.47, -2.49]<br><br>55-69: -8.46 [-12.69, -4.23] |

| | N trials | n | RaR (95% CI) | Mean control event rate(s) at 20 y unless stated | $I^2$ | Absolute effect with screening [95% CI] per 1,000 | Absolute difference (95% CI) per 1,000 |
|---|---|---|---|---|---|---|---|
| PSA OR DRE | 1 | 27,481 | 1.01 [0.96, 1.07] | 55: 249 per 1000<br><br>55-69: 423 per 1000 | NA | 55: 251.49 [239.04, 266.43]<br><br>55-69: 427.23 [406.08, 452.61] | 55: 2.49 [-9.96, 17.43]<br><br>55-69: 4.23 [-16.92, 29.61] |
| 55-69y: PSA +/- DRE vs. PSA OR DRE; **p=0.08** (among moderate/high intensity interventions) | | | | | | | |
| PSA +/- DRE | 7 | 162,236 | 0.99 [0.98, 1.00] | 55: 249 per 1000<br><br>55-69: 423 per 1000 | NA (data across sites) | 55: 246.51 [244.02, 249.00]<br><br>55-69: 418.77 [414.54, 423.00] | 55: -2.49 [-4.98, 0.00]<br><br>55-69: -4.23 [-8.46, 0.00] |
| PSA OR DRE | 1 | 66,542 | 0.96 [0.94, 0.99] | 55: 249 per 1000<br><br>55-69: 423 per 1000 | NA | 55: 239.04 [234.06, 246.51]<br><br>55-69: 406.08 [397.62, 418.77] | 55: -9.96 [-14.94, -2.49]<br><br>55-69: -16.92 [-25.38, -4.23] |
| 70-74y: PSA +/- DRE vs. PSA OR DRE; p=NA | | | | | | | |
| PSA +/- DRE | 0 | NA | NA | NA | NA | NA | NA |
| PSA OR DRE | All studies | | | | | | |

#### 3. Incidence of metastatic cancer (through follow-up)

| | N trials | n | RaR (95% CI) | Mean control event rate(s) at 20 y unless stated | $I^2$ | Absolute effect with screening [95% CI] per 1,000 | Absolute difference (95% CI) per 1,000 |
|---|---|---|---|---|---|---|---|
| **Metastatic cancer - through follow-up** | | | | | | | |
| All studies | | | | | | | |
| 50-54y | 0 | | | | | | |
| 55-69y | 5 | 153,494 | 0.73 [0.59, 0.90] | 55: 13 per 1000<br><br>55-69: 25 per 1000 | 68% | 55: 9.49 [7.67, 11.70]<br><br>55-69: 18.25 [14.75, 22.50] | 55: -3.51 [-5.33, -1.30]<br><br>55-69: -6.75 [-10.25, -2.50] |
| 70-74y | 1 | 7,069 | 1.04 [0.79, 1.37] | 29 per 1000 [15y] | NA | 30.16 [22.91, 39.73] | 1.16 [-6.09, 10.73] |
| 55-69 y: By intensity (e.g., low intensity [one screen] vs. moderate intensity [2-4 rounds [offered to majority] &/or 3+ year screening interval] vs. high intensity [5+ rounds &/or annual/biennial screening]); p=0.28 | | | | | | | |
| Low intensity | 0 | NA | NA | NA | NA | NA | NA |
| Moderate intensity | 3 | 64,959 | 0.66 [0.58, 0.76] | 55: 13 per 1000<br><br>55-69: 25 per 1000 | 0% | 55: 8.58 [7.54, 9.88]<br><br>55-69: 16.50 [14.50, 19.00] | 55: -4.42 [-5.46, -3.12]<br><br>55-69: -8.50 [-10.50, -6.00] |
| High intensity | 2 | 88,535 | 0.82 [0.57, 1.20] | 55: 13 per 1000<br><br>55-69: 25 per 1000 | 78% | 55: 10.66 [7.41, 15.60]<br><br>55-69: 20.50 [14.25, 30.00] | 55: -2.34 [-5.59, 2.60]<br><br>55-69: -4.50 [-10.75, 5.00] |

| | N trials | n | RaR (95% CI) | Mean control event rate(s) at 20 y unless stated | $I^2$ | Absolute effect with screening [95% CI] per 1,000 | Absolute difference (95% CI) per 1,000 |
|---|---|---|---|---|---|---|---|
| 55-69y: high vs lower ROB; **p=0.0005 (among moderate/high intensity interventions)** | | | | | | | |
| High ROB | 1 | 76,683 | 0.98 [0.81, 1.18] | 55: 13 per 1000<br><br>55-69: 25 per 1000 | NA | 55: 12.74 [10.53, 15.34]<br><br>55-69: 24.50 [20.25, 29.50] | 55: -0.26 [-2.47, 2.34]<br><br>55-69: -0.50 [-4.75, 4.50] |
| Lower ROB | 4 | 76,811 | 0.66 [0.59, 0.75] | 55: 13 per 1000<br><br>55-69: 25 per 1000 | 0% | 55: 8.58 [7.67, 9.75]<br><br>55-69: 16.50 [14.75, 18.75] | 55: -4.42 [-5.33, -3.25]<br><br>55-69: -8.50 [-10.25, -6.25] |
| 55-69y: 55-69 vs broader age; **p=0.0005 (among moderate/high intensity interventions)** | | | | | | | |
| 55-69y | 4 | 76,811 | 0.66 [0.59, 0.75] | 55: 13 per 1000<br><br>55-69: 25 per 1000 | 0% | 55: 8.58 [7.67, 9.75]<br><br>55-69: 16.50 [14.75, 18.75] | 55: -4.42 [-5.33, -3.25]<br><br>55-69: -8.50 [-10.25, -6.25] |
| Broader age range (PLCO 55-74) | 1 | 76,683 | 0.98 [0.81, 1.18] | 55: 13 per 1000<br><br>55-69: 25 per 1000] | NA | 55: 12.74 [10.53, 15.34]<br><br>55-69: 24.50 [20.25, 29.50] | 55: -0.26 [-2.47, 2.34]<br><br>55-69: -0.50 [-4.75, 4.50] |
| 55-69y: PSA +/- DRE vs. PSA OR DRE; **p=0.005 (among moderate/high intensity interventions)** | | | | | | | |
| PSA +/- DRE | 4 | 76,811 | 0.66 [0.59, 0.75] | 55: 13 per 1000<br><br>55-69: 25 per 1000 | 0% | 55: 8.58 [7.67, 9.75]<br><br>55-69: 16.50 [14.75, 18.75] | 55: -4.42 [-5.33, -3.25]<br><br>55-69: -8.50 [-10.25, -6.25] |
| PSA OR DRE | 1 | 76,683 | 0.98 [0.81, 1.18] | 55: 13 per 1000<br><br>55-69: 25 per 1000 | NA | 55: 12.74 [10.53, 15.34]<br><br>55-69: 24.50 [20.25, 29.50] | 55: -0.26 [-2.47, 2.34]<br><br>55-69: -0.50 [-4.75, 4.50] |

#### 4. Incidence of metastatic cancer (at diagnosis)

| | N trials | n | RaR (95% CI) | Mean control event rate(s) at 20 y unless stated | $I^2$ | Absolute effect with screening [95% CI] per 1,000 | Absolute difference (95% CI) per 1,000 |
|---|---|---|---|---|---|---|---|
| **Metastatic cancer at diagnosis** | | | | | | | |
| All studies | | | | | | | |
| 50-54y | 2 | 119,728 | 0.71 [0.30, 1.67] | 5 per 1000 | 25% | 3.55 [1.50, 8.35] | -1.45 [-3.50, 3.35] |
| 55-69y | 9 | 518,693 | 0.76 [0.63, 0.92] | 55: 8 per 1000<br><br>55-69: 16 per 1000 | 80% | 55: 6.08 [5.04, 7.36]<br><br>55-69: 12.16 [10.08, 14.72] | 55: -1.92 [-2.96, -0.64]<br><br>55-69: -3.84 [-5.92, -1.28] |
| 70-74y | 2 | 17,205 | 0.63 [0.47, 0.86] | 18 per 1000 | 0% | 11.34 [8.46, 15.48] | -6.66 [-9.54, -2.52] |
| 50-54y: By intensity (low intensity [one screen] vs. moderate intensity [2-4 rounds [offered to majority] &/or ≥3 y screening interval] vs. high intensity [5+ rounds &/or annual/biennial screening]); p=0.25 | | | | | | | |

| | N trials | n | RaR (95% CI) | Mean control event rate(s) at 20 y unless stated | $I^2$ | Absolute effect with screening [95% CI] per 1,000 | Absolute difference (95% CI) per 1,000 |
|---|---|---|---|---|---|---|---|
| Low intensity | 1 | 118,599 | 0.84 [0.72, 0.99] | 5 per 1000 | NA | 4.20 [3.60, 4.95] | -0.80 [-1.40, -0.05] |
| Moderate intensity | 1 | 1,129 | 0.23 [0.03, 2.06] | 5 per 1000 | NA | 1.15 [0.15, 10.30] | -3.85 [-4.85, 5.30] |
| High intensity | 0 | NA | NA | NA | NA | NA | NA |
| 55-69 y: By intensity (e.g., low intensity [one screen] vs. moderate intensity [2-4 rounds [offered to majority] &/or 3+ year screening interval] vs. high intensity [5+ rounds &/or annual/biennial screening]); **p=81 (p=0.45 for low vs. moderate or high intensity)** | | | | | | | |
| Low intensity | 1 | 290,122 | 0.84 [0.78, 0.91] | 55: 8 per 1000<br><br>55-69: 16 per 1000 | NA | 55: 6.72 [6.24, 7.28]<br><br>55-69: 13.44 [12.48, 14.56] | 55: -1.28 [-1.76, -0.72]<br><br>55-69: -2.56 [-3.52, -1.44] |
| Moderate intensity | 6 | 150,175 | 0.75 [0.52, 1.07] | 55: 8 per 1000<br><br>55-69: 16 per 1000 | 83% | 55: 6.00 [4.16, 8.56]<br><br>55-69: 12.00 [8.32, 17.12] | 55: -2.00 [-3.84, 0.56]<br><br>55-69: -4.00 [-7.68, 1.12] |
| High intensity | 2 | 78,396 | 0.83 [0.67, 1.02] | 55: 8 per 1000<br><br>55-69: 16 per 1000 | 13% | 55: 6.64 [5.36, 8.16]<br><br>55-69: 13.28 [10.72, 16.32] | 55: -1.36 [-2.64, 0.16]<br><br>55-69: -2.72 [-5.28, 0.32] |
| Moderate/high intensity | 8 | 228,571 | 0.76 [0.59, 0.98] | 55: 8 per 1000<br><br>55-69: 16 per 1000 | 79% | 55: 6.08 [4.72, 7.84]<br><br>55-69: 12.16 [9.44, 15.68] | 55: -1.92 [3.28, -0.16]<br><br>55-69: -3.84 -6.56, -0.32] |
| 55-69y: high vs lower ROB; p=0.25 **(among moderate/high intensity interventions)** | | | | | | | |
| High ROB | 2 | 68,739 | 1.21 [0.50, 2.94] | 55: 8 per 1000<br><br>55-69: 16 per 1000 | 52% | 55: 9.68 [4.00, 23.52]<br><br>55-69: 19.36 [8.00, 47.04] | 55: 1.68 [-4.00, 15.52]<br><br>55-69: 3.36 [-8.00, 31.04] |
| Lower ROB | 6 | 159,832 | 0.70 [0.53, 0.93] | 55: 8 per 1000<br><br>55-69: 16 per 1000 | 81% | 55: 5.60 [4.24, 7.44]<br><br>55-69: 11.20 [8.48, 14.88] | 55: -2.40 [-3.76, -0.56]<br><br>55-69: -4.80 [-7.52, -1.12] |
| 70-74y: high vs lower ROB; p=0.64 | | | | | | | |
| High ROB | 1 | 10,136 | 0.58 [0.35, 0.94] | 18 per 1000 [15y] | NA | 10.44 [6.30, 16.92] | -7.56 [-11.70, -1.08] |
| Lower ROB | 1 | 7,069 | 0.67 [0.46, 0.99] | 18 per 1000 [15y] | NA | 12.06 [8.28, 17.82] | -5.94 [-9.72, -0.18] |
| 55-69y: PSA +/- DRE vs. PSA OR DRE; **p=0.04 (among moderate/high intensity interventions)** | | | | | | | |
| PSA +/- DRE | 6 | 153,467 | 0.66 [0.50, 0.87] | 55: 8 per 1000<br><br>55-69: 16 per 1000 | 76% | 55: 5.28 [4.00, 6.96]<br><br>55-69: 10.56 [8.00, 13.92] | 55: -2.72 [-4.00, -1.04]<br><br>55-69: -5.44 [-8.00, -2.08] |
| PSA OR DRE | 2 | 75,104 | 1.05 [0.74, 1.50] | 55: 8 per 1000<br><br>55-69: 16 per 1000 | 48% | 55: 8.40 [5.92, 12.00]<br><br>55-69: 16.80 [11.84, 24.00] | 55: 0.40 [-2.08, 4.00]<br><br>55-69: 0.80 [-4.16, 8.00] |
| 70-74y: PSA +/- DRE vs. PSA OR DRE; p=0.64 | | | | | | | |

| | N trials | n | RaR (95% CI) | Mean control event rate(s) at 20 y unless stated | $I^2$ | Absolute effect with screening [95% CI] per 1,000 | Absolute difference (95% CI) per 1,000 |
|---|---|---|---|---|---|---|---|
| PSA +/- DRE | 1 | 7,069 | 0.67 [0.46, 0.99] | 18 per 1000 [15y] | NA | 12.06 [8.28, 17.82] | -5.94 [-9.72, -0.18] |
| PSA OR DRE | 1 | 10,136 | 0.58 [0.35, 0.94] | 18 per 1000 [15y] | NA | 10.44 [6.30, 16.92] | -7.56 [-11.70, -1.08] |

#### 5. Overdiagnosis

| Outcome and categories | N trials | n | RaR [95% CI] | Mean control event rate[s] at 20 y unless stated | $I^2$ | Absolute effect with screening [95% CI] per 1,000 | Absolute difference [95% CI] per 1,000 |
|---|---|---|---|---|---|---|---|
| **Overdiagnosis** | | | | | | | |
| All studies | | | | | | | |
| 50-54y | 3 | 127,773 | 1.38 [0.90, 2.10] | 42 per 1000 | 96% | 57.96 [37.80, 88.20] | 15.96 [-4.20, 46.20] |
| 55-69y | 11 | 622,791 | 1.23 [1.12, 1.34] | 55: 62 per 1000<br>55-69: 77 per 1000 | 95% | 55: 76.26 [69.44, 83.08]<br>55-69: 94.71 [86.24, 103.18] | 55: 14.26 [7.44, 21.08]<br>55-69: 17.71 [9.24, 26.18] |
| 70-74y | 2 | 13,678 | 1.27 [0.93, 1.73] | 67 per 1000 [15y] | 92% | 85.09 [62.31, 115.91] | 18.09 [-4.69, 48.91] |
| 50-54y: By intensity [low intensity [one screen] vs. moderate intensity [2-4 rounds [offered to majority] &/or ≥3 y screening interval] vs. high intensity [5+ rounds &/or annual/biennial screening]]; **p<0.00001 (p<0.0001 for low vs. moderate/high)** | | | | | | | |
| Low intensity | 1 | 118,599 | 1.05 [0.99, 1.12] | 42 per 1000 | NA | 44.10 [41.58, 47.04] | 2.10 [-0.42, 5.04] |
| Moderate intensity | 1 | 1,129 | 1.43 [0.88, 2.33] | 42 per 1000 | NA | 60.06 [36.96, 97.86] | 18.06 [-5.04, 55.86] |
| High intensity | 1 | 8,045 | 1.77 [1.54, 2.03] | 42 per 1000 | NA | 74.34 [64.68, 85.26] | 32.34 [22.68, 43.26] |
| Moderate/high intensity | 2 | 9,174 | 1.74 [1.52, 1.99] | 42 per 1000 | 0% | 73.08 [63.84, 83.58] | 31.08 [21.84, 41.58] |
| 55-69 y: By intensity [e.g., low intensity [one screen] vs. moderate intensity [2-4 rounds [offered to majority] &/or 3+ year screening interval] vs. high intensity [5+ rounds &/or annual/biennial screening]]; **p=0.07** (**p=0.02** for low vs. moderate/high) | | | | | | | |
| Low intensity | 2 | 317,603 | 1.07 [1.04, 1.10] | 55: 62 per 1000<br>55-69: 77 per 1000 | 0% | 55: 66.34 [64.48, 68.20]<br>55-69: 82.39 [80.08, 84.70] | 55: 4.34 [2.48, 6.20]<br>55-69: 5.39 [3.08, 7.70] |
| Moderate intensity | 7 | 226,792 | 1.47 [1.11, 1.94] | 55: 62 per 1000<br>55-69: 77 per 1000 | 99% | 55: 91.14 [68.82, 120.28]<br>55-69: 113.19 [85.4,149.38] | 55: 29.14 [6.82, 58.28]<br>55-69: 36.19 [8.47, 72.38] |
| High intensity | 2 | 78,396 | 1.17 [0.93, 1.47] | 55: 62 per 1000 | 95% | 55: 72.54 [57.66, 91.14] | 55: 10.54 [-4.34, 29.14] |

| Outcome and categories | N trials | n | RaR [95% CI] | Mean control event rate[s] at 20 y unless stated | $I^2$ | Absolute effect with screening [95% CI] per 1,000 | Absolute difference [95% CI] per 1,000 |
|---|---|---|---|---|---|---|---|
| | | | | 55-69: 77 per 1000 | | 55-69: 90.09 [71.61, 113.19] | 55-69: 13.09 [-5.39, 36.19] |
| Moderate/high | 9 | 305,188 | 1.26 [1.13, 1.41] | 55: 62 per 1000<br>55-69: 77 per 1000 | 96% | 55: 78.12 [70.06, 87.42]<br>55-69: 97.02 [87.01, 108.57] | 55: 16.12 [8.06, 25.42]<br>55-69: 20.02 [10.01, 31.57] |
| 50-54y: high vs lower ROB; p=0.41 **(among moderate/high intensity interventions)** | | | | | | | |
| High ROB | 1 | 1,129 | 1.43 [0.88, 2.33] | 42 per 1000 | NA | 60.06 [36.96, 97.86] | 18.06 [-5.04, 55.86] |
| Lower ROB | 1 | 8,045 | 1.77 [1.54, 2.03] | 42 per 1000 | NA | 74.34 [64.68, 85.26] | 32.34 [22.68, 43.26] |
| 55-69y: high vs lower ROB; p=0.48 **(among low intensity interventions)** | | | | | | | |
| High ROB | 1 | 290,122 | 1.07 [1.04, 1.10] | 55: 62 per 1000<br>55-69: 77 per 1000 | NA | 55: 66.34 [64.48, 68.20]<br>55-69: 82.39 [80.08, 84.70] | 55: 4.34 [2.48, 6.20]<br>55-69: 5.39 [3.08, 7.70] |
| Lower ROB | 1 | 27,481 | 1.12 [0.99, 1.26] | 55: 62 per 1000<br>55-69: 77 per 1000 | NA | 55: 69.44 [61.38, 78.12]<br>55-69: 86.24 [76.23, 97.02] | 55: 7.44 [-0.62, 16.12]<br>55-69: 9.24 [-0.77, 20.02] |
| 55-69y: high vs lower ROB; **p<0.0001; (among moderate/high intensity interventions)** | | | | | | | |
| High ROB | 6 | 248,600 | 1.13 [1.07, 1.19] | 55: 62 per 1000<br>55-69: 77 per 1000 | 69% | 55: 70.06 [66.34, 73.78]<br>55-69: 87.01 [82.39, 91.63] | 55: 8.06 [4.34, 11.78]<br>55-69: 10.01 [5.39, 14.63] |
| Lower ROB | 3 | 56,588 | 1.48 [1.31, 1.68] | 55: 62 per 1000<br>55-69: 77 per 1000 | 84% | 55: 91.76 [81.22, 104.16]<br>55-69: 113.96 [100.87, 129.36] | 55: 29.76 [19.22, 42.16]<br>55-69: 36.96 [23.87, 52.36] |
| 70-74y: high vs lower ROB; **p=0.0004** | | | | | | | |
| High ROB | 1 | 10,136 | 1.09 [0.98, 1.20] | 67 per 1000 [15y] | NA | 73.03 [65.66, 80.40] | 6.03 [-1.34, 13.40] |
| Lower ROB | 1 | 7,069 | 1.49 [1.29, 1.72] | 67 per 1000 [15y] | NA | 99.83 [86.43, 115.24] | 32.83 [19.43, 48.24] |
| 50-54y: 50-54 vs broader age [10-25% outside range]; p=0.41 **(among moderate/high intensity interventions)** | | | | | | | |
| 50-54y | 1 | 8,045 | 1.77 [1.54, 2.03] | 42 per 1000 | NA | 74.34 [64.68, 85.26] | 32.34 [22.68, 43.26] |
| 45-54y | 1 | 1,129 | 1.43 [0.88, 2.33] | 42 per 1000 | NA | 60.06 [36.96, 97.86] | 18.06 [-5.04, 55.86] |
| 50-54y: PSA +/- DRE vs. PSA OR DRE; p=NA | | | | | | | |
| PSA +/- DRE | All studies | | | | | | |
| PSA OR DRE | 0 | NA | NA | NA | NA | NA | NA |
| 55-69y: PSA +/- DRE vs. PSA OR DRE; p=0.48 **(among low intensity interventions)** | | | | | | | |
| PSA +/- DRE | 1 | 290,122 | 1.07 [1.04, 1.10] | 55: 62 per 1000 | NA | 55: 66.34 [64.48, 68.20] | 55: 4.34 [2.48, 6.20] |

| Outcome and categories | N trials | n | RaR [95% CI] | Mean control event rate[s] at 20 y unless stated | I² | Absolute effect with screening [95% CI] per 1,000 | Absolute difference [95% CI] per 1,000 |
|---|---|---|---|---|---|---|---|
| | | | | 55-69: 77 per 1000 | | 55-69: 82.39 [80.08, 84.70] | 55-69: 5.39 [3.08, 7.70] |
| PSA OR DRE | 1 | 27,481 | 1.12 [0.99, 1.26] | 55: 62 per 1000<br><br>55-69: 77 per 1000 | NA | 55: 69.44 [61.38, 78.12]<br><br>55-69: 86.24 [76.23, 97.02] | 55: 7.44 [-0.62, 16.12]<br><br>55-69: 9.24 [-0.77, 20.02] |
| 55-69y: PSA +/- DRE vs. PSA OR DRE; **p=0.06 (among moderate/high intensity interventions)** | | | | | | | |
| PSA +/- DRE | 7 | 230,094 | 1.31 [1.15, 1.50] | 55: 62 per 1000<br><br>55-69: 77 per 1000 | 95% | 55: 81.22 [71.30, 93.00]<br><br>55-69: 100.87 [88.55, 115.50] | 55: 19.22 [9.30, 31.00]<br><br>55-69: 23.87 [11.55, 38.50] |
| PSA OR DRE | 2 | 75,104 | 1.10 [0.97, 1.25] | 55: 62 per 1000<br><br>55-69: 77 per 1000 | 76% | 55: 68.20 [60.14, 77.50]<br><br>55-69: 84.70 [74.69, 96.25] | 55: 6.20 [-1.86, 15.50]<br><br>55-69: 7.70 [-2.31, 19.25] |
| 70-74y: PSA +/- DRE vs. PSA OR DRE; **p=0.0004** | | | | | | | |
| PSA +/- DRE | 1 | 7,069 | 1.49 [1.29, 1.72] | 67 per 1000 [15y] | NA | 99.83 [86.43, 115.24] | 32.83 [19.43, 48.24] |
| PSA OR DRE | 1 | 10,136 | 1.09 [0.98, 1.20] | 67 per 1000 [15y] | NA | 73.03 [65.66, 80.40] | 6.03 [-1.34, 13.40] |

#### Within-study analysis of effects by screening intensity or per protocol estimates

| Study | Outcome & follow-up timing | Results | Authors' definition of per protocol analysis |
|---|---|---|---|
| **ERSPC (Roobol 2025)**<br><br>**All sites except France** | PCa mortality, median 23 y | After correction for nonattendance, the risk ratio was 0.84 (95% CI, 0.76 to 0.92) for men attending at least one screening round. | Attenders of 1+ screening round adjustment for nonparticipation made with the use of Cuzick's method. The proportion of complete nonattendees (ie, never participating) in the screening group and the PCa mortality among them were calculated. The control group was modeled to include a nonattender subgroup of equal size and mortality rate, enabling calculation of the adjusted mortality rate among screening participants. |
| **ERSPC (Hugosson, 2019)**<br><br>**All sites except France** | PCa mortality in men attending screening at least once (N=60,261) vs. men in the control group (N=89,351)<br><br>Median of 15.5 y (≤16 y after randomization, through Dec 2014) | PCa mortality reduction in men attending at least one screening round: 25% (RaR: 0.75, 95% CI: 0.66, 0.75)<br><br>PCa mortality reduction in men attending at least two screening rounds: 48% (rate ratio: 0.52, 95% CI: 0.42, 0.63) if no mortality reduction was postulated from one test only, 43% (RR 0.57, 95% CI 0.47–0.70) if a mortality reduction of 10% was postulated, and 25% (RR 0.75, 95% CI 0.60– | To assess the impact of attending at least one screening round, adjusted rate ratios were calculated accounting for nonparticipation. The proportion of complete nonattendees (ie, never participating) in the screening group and the PCa mortality among them were calculated. The control group was modeled to include a nonattender subgroup of equal size and mortality rate, enabling calculation of the |

| Study | Outcome & follow-up timing | Results | Authors' definition of per protocol analysis |
|---|---|---|---|
| | | 0.92) if first screening was as effective as the following rounds. | adjusted mortality rate among screening participants. |
| **ERSPC (Schröder, 2014)**<br><br>**All sites except France** | PCa mortality in men screened vs. men in the control group<br><br>Median of 13.0 y (median range: 10.2-13.0 y across sites excluding France; through Dec 2010) | PCa mortality rate ratio adjusted for non-participation, core age group: 0.73 (95% CI: 0.61, 0.88), p<0.001<br><br>PCa mortality rate ratio adjusted for non-participation, all ages: 0.78 (95% CI: 0.66, 0.92) | Mortality in men screened, corrected for selection bias due to non-participation. |
| **ERSPC Sweden (Franlund, 2022)** | PCa mortality among all ages 50-64 attending screening at least once (N=7,635) vs. men in the control group (N=9,949)<br><br>At 22.0 y (through Dec 2016) | PCa mortality: 65/7,635 vs. 158/9,949 in CG<br><br>Correction for nonattendance (Cuzick method):<br>RR: 0.59 (95% CI: 0.43, 0.80) (vs. nonadjusted) at 22 years 0.71 (95% CI: 0.55, 0.91)) | Men in the screening group were classified as attenders (attended at least once) or nonattenders (never attended).<br>Analysis corrects for nonattendance (Cuzick method) |
| **ERSPC Sweden (Hugosson, 2018)** | PCa incidence and mortality and all-cause mortality among attendees in the screening group vs. an adjusted control group.<br><br>Metastatic PCa among attendees in the screening arm vs. control arm.<br><br>18 yr after randomization (through Dec 2012) | PCa incidence rate ratio:<br>50-54 y: 1.87 (95% CI: 1.62, 2.17)<br>55-59 y: 1.43 (95% CI: 1.23, 1.66)<br>60-64 y: 1.47 (95% CI: 1.25, 1.73)<br><br>PCa mortality rate ratios:<br>50-54 y: 0.31 (95% CI: 0.10, 0.94)<br>55-59 y: 0.37 (95% CI: 0.20, 0.67)<br>60-64 y: 0.80 (95% CI: 0.50, 1.27)<br><br>All-cause mortality rate ratios:<br>50-54 y: 1.15 (95% CI: 0.97, 1.28)<br>55-59 y: 0.98 (95% CI: 0.87, 1.09)<br>60-64 y: 0.94 (95% CI: 0.86, 1.04)<br><br>Metastatic PCa, 50-64 y: 116/7,647 in IG vs. 179/9,949 in CG | Men who attended screening at least once. The control group was adjusted by subtracting the incidence and mortality rate of prostate cancer among non-attendees in the screening group from the control group.<br><br>No adjustment for metastatic cancer. |
| **ERSPC Finland (Pakarainen, 2021)** | PCa incidence hazard ratio in men attending 1, 2, or 3 rounds of screening in IG compared to the men in CG<br><br>Max of 17 y (through Dec 2015) | PCa incidence HR (age-adjusted) by no. of screening rounds attended in IG vs. CG:<br>1 round: 1.52 (95% CI: 1.42, 1.63)<br>2 rounds: 1.18 (95% CI; 1.10, 1.27)<br>3 rounds: 1.33 (95% CI: 1.22, 1.45) | The incidence of PC among men in the screening arm who had been screened 0 to 3 times was compared with incidence in the control arm. Adjusted for age. |
| **ERSPC Finland (Pakarainen, 2019)** | PCa mortality and all-cause mortality in men attending 1, 2, or 3 rounds of screening in IG compared to the men in CG<br><br>Max of 15 y (through Dec 2013) | PCa mortality ORs (age-adjusted) by no. of screening rounds attended in IG vs. CG:<br>1 round: 1.68 (95% CI: 1.31, 2.16)<br>2 rounds: 0.50 (95% CI: 0.37, 0.69)<br>3 rounds: 0.16 (95% CI: 0.08, 0.32) | Number of screening rounds attended by men in the screening group. |

| Study | Outcome & follow-up timing | Results | Authors' definition of per protocol analysis |
|---|---|---|---|
| | | All-cause mortality ORs (age-adjusted) by no. of screening rounds attended in IG vs. CG:<br>1 round: 0.96 (95% CI: 0.92, 1.01)<br>2 rounds: 0.73 (95% CI: 0.70, 0.77)<br>3 rounds: 0.57 (95% CI: 0.53, 0.62) | |
| **ERSPC France (Villers, 2020)** | PCa incidence among screening attenders, ie., screened at least once, vs. men in CG<br><br>Median of 9.5 (IQR: 8.9-9.8) y | IG: 938/12,030<br>CG: 2,240/38,161 | PCa incidence among screening attenders |
| **Quebec RCT (Labrie, 2004)** | PCa mortality among screened men in IG vs. unscreened men in CG<br><br>11 y after randomization (median of 7.9 y in IG), through December 31, 1999 | IG: 10/7,348<br>CG: 74/14,231<br>Relative risk: 0.38 (95% CI: 0.20, 0.73), p<0.002<br>When adjusting for differences in follow-up between groups (max 7.93 y): RR 0.49 (95% CI: 0.25, 0.99) | Cancer-specific death during the first 11 years in the groups of screened versus non screened men originally invited and not invited for screening, respectively. |

#### Within-study analysis for specific populations

| Study | Subgroup categories | n/N [%]<br>*IG vs CG* | Findings<br>*IG vs CG* | Ratio [95% CI] | Absolute effect with screening [95% CI] per 1,000[†] | Absolute difference [95% CI] per 1,000 |
|---|---|---|---|---|---|---|
| **Outcome: Prostate cancer morality** | | | | | | |
| Race/ethnicity; p=0.75 | | | | | | |
| PLCO | Black | 1,713/34,756 [4.9%] vs. 1,657/33,792 [4.9%] | 24/1,713 [1.4%] vs. 21/1,657 [1.3%] | RR: 1.11 [0.62, 1.98] | 16.65 [9.30, 29.70] | 1.65 [-5.70, 14.70] |
| Miller 2018 (at 19 y for mortality) | White | 33,043/34,756 [95.1%] vs. 32,135/33,792 [95.1%] | 211/33,043 [0.64%] vs. 205/32,135 [0.64%] | RR: 1.00 [0.83, 1.21] | 15.00 [12.45, 18.15] | 0.00 [-2.55, 3.15] |
| **Outcome: Overdiagnosis** | | | | | | |
| Subgroup: Race/ethnicity; p=0.28 | | | | | | |
| PLCO | Black | 1,713/34,756 [4.9%] vs. 1,657/33,792 [4.9%] | 252/1,713 [14.7%] vs. 242/1,657 [14.6%] | RR: 1.01 [0.86, 1.19] | 77.77 [66.22, 91.63] | 0.77 [-10.78, 14.63] |
| Miller 2018 (at 13 y of FU for incidence) | White | 33,043/34,756 [95.1%] vs. 32,135/33,792 [95.1%] | 3,891/33,043 [11.8%] vs. 3,423/32,135 [10.7%] | RR: 1.11 [1.06, 1.15] | 85.47 [81.62, 88.55] | 8.47 [4.62, 11.55] |
| **Outcome: Metastatic cancer at diagnosis** | | | | | | |

| Study | Subgroup categories | n/N [%] *IG vs CG* | Findings *IG vs CG* | Ratio [95% CI] | Absolute effect with screening [95% CI] per 1,000† | Absolute difference [95% CI] per 1,000 |
|---|---|---|---|---|---|---|
| Subgroup: Race/ethnicity; p=0.43 | | | | | | |
| PLCO | Black | 1,713/34,756 [4.9%] vs. 1,657/33,792 [4.9%] | 16/1,713 [0.93%] vs. 21/1,657 [1.3%] | RR:0.74 [0.39, 1.41] | 11.84 [6.24, 22.56] | -4.16 [-9.76, 6.56] |
| Miller 2018 (at 13 y of FU for incidence) | White | 33,043/34,756 [95.1%] vs. 32,135/33,792 [95.1%] | 124/33,043 [0.38%] vs. 124/32,135 [0.39%] | RR: 0.97 [0.76, 1.25] | 15.52 [12.16, 20.00] | -0.48 [-3.84, 4.00] |

CG: control group; aHR: adjusted hazard ratio; IG: intervention group; RR: risk ratio
*Among men with complete information about family history in a first-degree relative. Estimates adjusted for age, race, BMI, and smoking.
† Calculated using control event rates (CER) in the 55-69-year-old age group category (CER: 15/1000 for PCa mortality; 77/1000 for overdiagnosis; 16/1000 for metastatic PCa at diagnosis).

**Figures**

**Prostate-cancer mortality by race/ethnicity**

| Study or Subgroup | PSA Events | PSA Total | Control Events | Control Total | Weight | Risk Ratio IV, Fixed, 95% CI |
|---|---|---|---|---|---|---|
| **1.18.1 Black** | | | | | | |
| PLCO | 24 | 1713 | 21 | 1657 | 9.8% | 1.11 [0.62, 1.98] |
| **Subtotal (95% CI)** | | **1713** | | **1657** | **9.8%** | **1.11 [0.62, 1.98]** |
| Total events | 24 | | 21 | | | |
| Heterogeneity: Not applicable | | | | | | |
| Test for overall effect: Z = 0.34 (P = 0.74) | | | | | | |
| **1.18.2 White** | | | | | | |
| PLCO | 211 | 33043 | 205 | 32135 | 90.2% | 1.00 [0.83, 1.21] |
| **Subtotal (95% CI)** | | **33043** | | **32135** | **90.2%** | **1.00 [0.83, 1.21]** |
| Total events | 211 | | 205 | | | |
| Heterogeneity: Not applicable | | | | | | |
| Test for overall effect: Z = 0.01 (P = 0.99) | | | | | | |
| **Total (95% CI)** | | **34756** | | **33792** | **100.0%** | **1.01 [0.84, 1.21]** |
| Total events | 235 | | 226 | | | |
| Heterogeneity: Chi² = 0.10, df = 1 (P = 0.75); I² = 0% | | | | | | |
| Test for overall effect: Z = 0.12 (P = 0.91) | | | | | | |
| Test for subgroup differences: Chi² = 0.10, df = 1 (P = 0.75), I² = 0% | | | | | | |

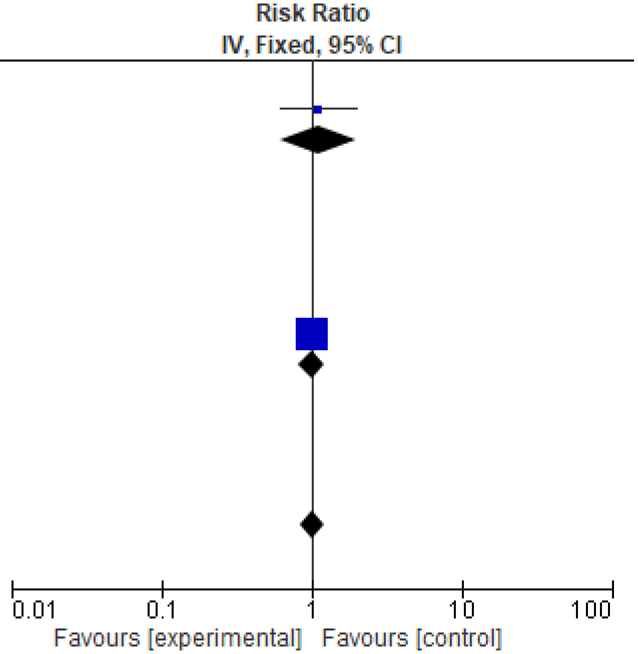

**Overdiagnosis by race/ethnicity**

| | PSA | | Control | | | Risk Ratio |
|---|---|---|---|---|---|---|
| Study or Subgroup | Events | Total | Events | Total | Weight | IV, Fixed, 95% CI |
| **1.19.1 Black** | | | | | | |
| PLCO | 252 | 1713 | 242 | 1657 | 6.6% | 1.01 [0.86, 1.19] |
| **Subtotal (95% CI)** | | **1713** | | **1657** | **6.6%** | **1.01 [0.86, 1.19]** |
| Total events | 252 | | 242 | | | |
| Heterogeneity: Not applicable | | | | | | |
| Test for overall effect: Z = 0.09 (P = 0.93) | | | | | | |
| **1.19.2 White** | | | | | | |
| PLCO | 3891 | 33043 | 3423 | 32135 | 93.4% | 1.11 [1.06, 1.15] |
| **Subtotal (95% CI)** | | **33043** | | **32135** | **93.4%** | **1.11 [1.06, 1.15]** |
| Total events | 3891 | | 3423 | | | |
| Heterogeneity: Not applicable | | | | | | |
| Test for overall effect: Z = 4.54 (P < 0.00001) | | | | | | |
| **Total (95% CI)** | | **34756** | | **33792** | **100.0%** | **1.10 [1.05, 1.15]** |
| Total events | 4143 | | 3665 | | | |
| Heterogeneity: $Chi^2 = 1.17$, df = 1 (P = 0.28); $I^2 = 14\%$ | | | | | | |
| Test for overall effect: Z = 4.41 (P < 0.0001) | | | | | | |
| Test for subgroup differences: $Chi^2 = 1.17$, df = 1 (P = 0.28), $I^2 = 14.5\%$ | | | | | | |

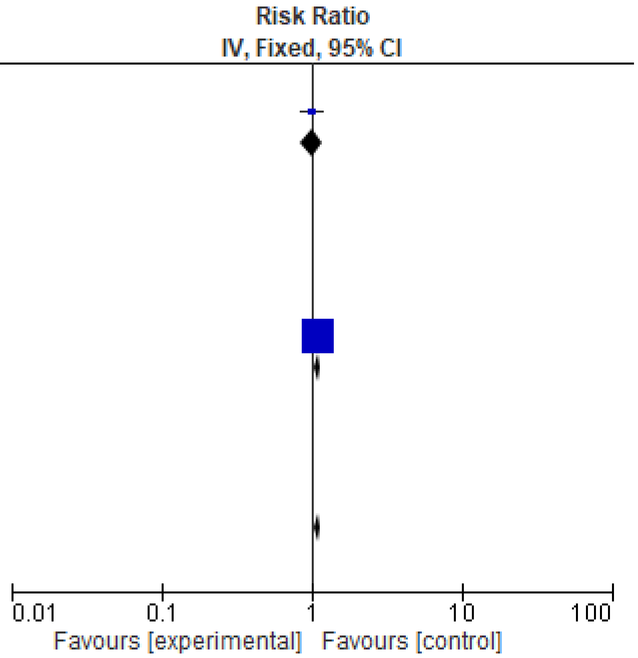

**Metastatic cancer at diagnosis by race/ethnicity**

| Study or Subgroup | PSA Events | PSA Total | Control Events | Control Total | Weight | Risk Ratio IV, Fixed, 95% CI |
|---|---|---|---|---|---|---|
| **1.20.1 Black** | | | | | | |
| PLCO | 16 | 1713 | 21 | 1657 | 12.9% | 0.74 [0.39, 1.41] |
| **Subtotal (95% CI)** | | **1713** | | **1657** | **12.9%** | **0.74 [0.39, 1.41]** |
| Total events | 16 | | 21 | | | |
| Heterogeneity: Not applicable | | | | | | |
| Test for overall effect: Z = 0.92 (P = 0.36) | | | | | | |
| **1.20.2 White** | | | | | | |
| PLCO | 124 | 33043 | 124 | 32135 | 87.1% | 0.97 [0.76, 1.25] |
| **Subtotal (95% CI)** | | **33043** | | **32135** | **87.1%** | **0.97 [0.76, 1.25]** |
| Total events | 124 | | 124 | | | |
| Heterogeneity: Not applicable | | | | | | |
| Test for overall effect: Z = 0.22 (P = 0.83) | | | | | | |
| **Total (95% CI)** | | **34756** | | **33792** | **100.0%** | **0.94 [0.74, 1.18]** |
| Total events | 140 | | 145 | | | |
| Heterogeneity: Chi² = 0.62, df = 1 (P = 0.43); I² = 0% | | | | | | |
| Test for overall effect: Z = 0.54 (P = 0.59) | | | | | | |
| Test for subgroup differences: Chi² = 0.62, df = 1 (P = 0.43), I² = 0% | | | | | | |

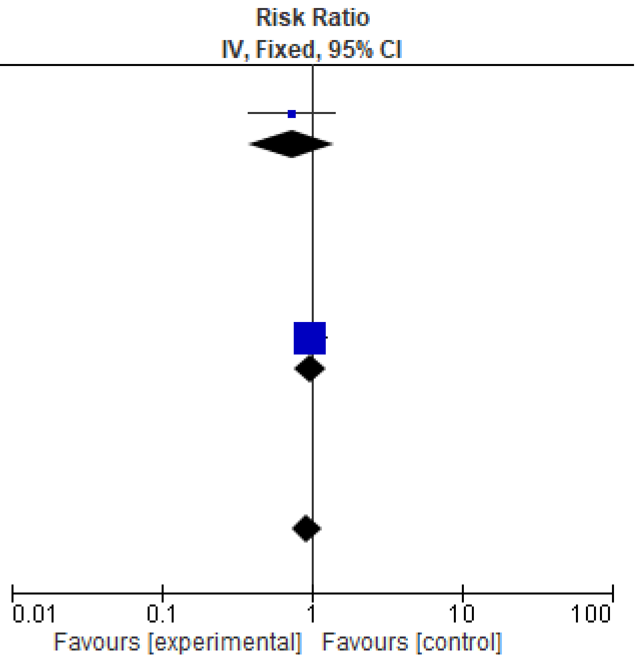

#### Findings from single study reporting on DRE screening

Conducted in Norrkoping Sweden, a quasi-randomized study (N=9026) screened men aged 50-69 every 3 years for 4 rounds using DRE alone for 2 rounds then adding PSA for 2 subsequent rounds, though a limited number were eligible (<70 years) for the last round. The only data eligible for this review were on overdiagnosis, for which there was an increase from screening (RR 1.50, 95% CI 1.17 to 1.92; 38.5 more cases per 1000, 95% CI 13 to 70 more). Only a subset of men with cancers detected early in the trial were followed for mortality.

Ref: Sandblom G, Varenhorst E, Rosell J, Löfman O, Carlsson P. Randomised prostate cancer screening trial: 20 year follow-up. BMJ. 2011;342.

#### Study characteristics of studies only reporting on FP and biopsy complications

| Characteristic | Rosario 2012 (PROBE; UK) | Sung 2021 (Hong Kong) | Ankerst 2009 (SABOR; US) | Hugosson 2022 (Goteburg-2, Sweden) |
|---|---|---|---|---|
| **Study design** | Uncontrolled prospective cohort (nested within RCT) | Uncontrolled prospective cohort | Uncontrolled prospective cohort | RCT |
| **Recruitment setting** | Primary care practices | Community (newspaper, radio & TV advertisements) | Not reported | Community (population-based) |
| **Enrolment period** | Feb 2006 to May 2008 | Aug 2018 to Apr 2020 | Starting in 2000 | 2015-2022 |
| **Eligible age range, yrs** | 50 to 69 years | Over the age of 50 | Not reported<br>Median (IQR):<br>No biopsy: 56.0 (28.3 to 88.6)<br>1 or more neg biopsies: 62.0 (33.8 to 82.2)<br>Prostate Ca: 62.8 (45.9 to 80.6) | 50 to 60 years |
| **Other eligibility criteria** | Asymptomatic men registered with a participating family physician | Asymptomatic men eligible for colorectal cancer screening | Men without diagnosis of prostate cancer | Men with prevalent prostate cancer or who died or emigrated between identification in the population register and randomization were excluded. |
| **Total screened, N** | 1,147 | 1,351 | 3,095 | 17,980 (reference group only) |

**Screening for prostate cancer using PSA with and without MRI: systematic reviews with meta-analysis**

Jennifer Pillay, Lindsay Gaudet, Sholeh Rahman, Roland Grad, Guylène Thériault, Philipp Dahm, Keith J Todd, Gail McCartney, Brett Thombs, Sabrina Saba, Lisa Hartling

| Characteristic | Rosario 2012 (PROBE; UK) | Sung 2021 (Hong Kong) | Ankerst 2009 (SABOR; US) | Hugosson 2022 (Goteburg-2, Sweden) |
|---|---|---|---|---|
| **Mean or median age at baseline, yrs** | 62.1 | 62 | No biopsy: 56.0<br>1 or more neg biopsies: 62.0<br>Prostate Ca: 62.8 | 56 |
| **Race/ethnicity, %** | White: 97.9%<br>Other: 0.5%<br>Missing: 1.5% | Not reported (Presumed to be primarily Asian) | White: 54.6%<br>Black: 12.4%<br>Hispanic: 32.4%<br>Other: 0.1% | Not reported |
| **Screening test(s)** | PSA | PSA +/- PHI | PSA or DRE | PSA |
| **Indication for biopsy** | PSA≥3.0 ng/ml | PSA >10 OR PSA ≥4 + PHI≥35 | PSA ≥2.5 ng/ml or abnormal DRE | PSA≥3.0 ng/ml |
| **Number of rounds** | 1 round | 1 round | Up to 6 rounds | 1 round |
| **Screening interval(s), yrs** | N/A | N/A | 1 year | N/A |
| **Screening rounds per man, mean** | N/A | N/A | 4 rounds | N/A |
| **Biopsy type** | TRUS 10+ core biopsy | Not reported | Not reported | TRUS 10-12 core |
| **Reported outcomes** | Biopsy complications | Biopsy complications | False positives | Biopsy complications |
| **Reported subgroups** | None | None | Race/ethnicity<br>Family history of PCa<br>PSA vs DRE | None |

#### Summary of findings for FPs and biopsy complications

| Outcome (Decision threshold) | No. participants (studies) | Cumulative incidence per 1000 (95 % CI) | Certainty of evidence | What happens? |
|---|---|---|---|---|
| Intention-to-Screen | | | | |
| 1+ positive screen not leading to cancer diagnosis (150 per 1000) | N=155931 (9 RCTs)<br>Reliance on lower risk of bias studies (N=104251; 6 RCTs) | 198 (143, 258) | ⊕⊕⊕⊝<br>**MODERATE** [1, 2]<br>due to inconsistency | Invitation to multiple rounds of PSA-based screening probably leads to 1 or more positive screening tests that do not lead to a prostate cancer diagnosis for at least 150 in 1000 men. |
| Any biopsy complication (100 per 1000) | No data | - | ⊕⊝⊝⊝<br>**VERY LOW** [3]<br>due to no evidence | We are uncertain about the cumulative incidence of biopsy complications after invitation to PSA-based screening. |
| Major/severe biopsy complications (50 per 1000) | N=38308 (1 RCT) | 1.64 (1.26, 2.08) | ⊕⊕⊝⊝<br>**LOW** [4, 5]<br>due to risk of bias and reporting bias | Invitation to multiple rounds of PSA-based screening probably leads to symptoms of infection after prostate biopsy for fewer than 50 in 1000 men. |
| Hospitalization after biopsy (6 per 1000) | 1 round<br>N=12629 (1 RCT) | 0.32 (0.09, 0.81) | ⊕⊕⊕⊝<br>**MODERATE** [5, 6]<br>due to reporting bias | Invitation to 1 round of PSA-based screening probably leads to hospitalization due to biopsy complications for fewer than 1 in 1000 men. |
| | Multiple rounds<br>N=21210 (1 RCT) | 2.07 (1.51, 2.78) | ⊕⊕⊕⊝<br>**MODERATE** [5]<br>due to reporting bias | Invitation to multiple rounds of PSA-based screening probably leads to hospitalization for biopsy complications in fewer than 2 per 1000 men. |
| Any infective symptoms (40 per 1000) | 1 round<br>N=33839 (2 RCTs) | 1.74 (0.03, 5.85) | ⊕⊕⊕⊕<br>**HIGH** | Invitation to 1 round of PSA-based screening results in symptoms of infection after biopsy for fewer than 6 per 1000 men. |
| | Multiple rounds<br>N=21210 (1 RCT) | 13.1 (11.6, 14.7) | ⊕⊕⊕⊝<br>**MODERATE** [5]<br>due to reporting bias | Invitation to multiple rounds of PSA-based screening probably results in symptoms of infection after biopsy for fewer than 15 per 1000 men. |
| Major infection (20 per 1000) | 1 round<br>No data | - | ⊕⊝⊝⊝<br>**VERY LOW** [3]<br>due to no evidence | We are uncertain about the cumulative incidence of any major infection from biopsy after 1 invitation to PSA-based screening. |
| | Multiple rounds<br>N=38308 (1 RCT)<br>*Infection requiring medical intervention* | 0.76 (0.50, 1.06) | ⊕⊕⊝⊝<br>**LOW** [4, 5]<br>due to risk of bias and reporting bias | Invitation to multiple rounds of PSA-based screening may result in fewer than 20 per 1000 men developing major infection after biopsy. |

| Outcome (Decision threshold) | No. participants (studies) | Cumulative incidence per 1000 (95 % CI) | Certainty of evidence | What happens? |
|---|---|---|---|---|
| Any bleeding complications (80 per 1000) | No data | - | ⊕⊖⊖⊖<br>**VERY LOW** [3]<br>due to no evidence | We are uncertain about the cumulative incidence of any severity of bleeding complications after 1 or more invitations to PSA-based screening. |
| Major bleeding complications (40 per 1000) | 1 round<br>No data | - | ⊕⊖⊖⊖<br>**VERY LOW** [3]<br>due to no evidence | We are uncertain about the cumulative incidence of any severity of bleeding complications after 1 or more invitations to PSA-based screening. |
| | Multiple rounds<br>N=38308 (1 RCT) | 0.37 (0.20, 0.59) | ⊕⊕⊖⊖<br>**LOW** [4, 5]<br>due to risk of bias and reporting bias | Invitation to multiple rounds of PSA-based screening may lead to major bleeding complications after biopsy for fewer than 40 per 1000 men. |
| Biopsy mortality (0.1 per 1000) | 1 round<br>N=320782 (10 RCTs) | 0.00 (0.00, 0.00) per 10,000 | ⊕⊕⊕⊕<br>**HIGH** | Invitation to 1 round of PSA-based screening leads death after biopsy in fewer than 1 per 10,000 men. |
| | Multiple rounds<br>N=111228 (8 RCTs) | 0 (0.00, 0.00) per 10,000 | ⊕⊕⊕⊕<br>**HIGH** | Invitation to multiple rounds of PSA-based screening leads to death after biopsy in fewer than 1 per 10,000 men. |
| Per Protocol | | | | |
| 1+ positive screen not leading to cancer diagnosis (150 per 1000) | N=115276 (9 RCTs, 1 OBS)<br>*Reliance on 9 RCTs (N=112181) due to indirectness of population in observational study* | 200 (164, 238) | ⊕⊕⊕⊕<br>**HIGH** [2] | Attending at least 1 round of PSA-based screening causes 1 or more positive screening test not leading to a cancer diagnosis in least 150 per 1000 men. |
| All biopsy complications (100 per 1000) | 1 round<br>N=18922 (1 OBS) | 49 (46, 53) | ⊕⊕⊖⊖<br>**LOW** [5, 7]<br>due to indirectness and reporting bias | Attending 1 round of PSA-based screening may lead to biopsy complications in fewer than 100 per 1000 men. |
| | Multiple rounds<br>N=19765 (1 RCT) | 236 (231, 242) | ⊕⊕⊕⊖<br>**MODERATE** [5, 8]<br>due to reporting bias | Attending at least 1 round of PSA-based screening probably leads to biopsy complication in at least 230 per 1000 men. |
| Major/severe biopsy complications (50 per 1000) | 1 round<br>N=20273 (2 OBS) | 2.33 (0.00, 14.04) | ⊕⊕⊕⊖<br>**MODERATE** [7]<br>due to indirectness | Attending 1 round of PSA-based screening probably leads to major/severe complications after biopsy for fewer than 15 per 1000 men. |

| Outcome (Decision threshold) | No. participants (studies) | Cumulative incidence per 1000 (95 % CI) | Certainty of evidence | What happens? |
|---|---|---|---|---|
| | Multiple rounds<br>N=35870 (1 RCT) | 1.76 (1.35, 2.25) | ⊕⊕⊝⊝<br>**LOW** [4, 5]<br>due to risk of bias and reporting bias | Attending at least 1 round of PSA-based screening may lead to fewer than 50 per 1000 men experiencing major/severe (“very bothersome”) complications after biopsy. |
| Hospitalization after biopsy<br>(6 per 1000) | 1 round<br>N=24916 (1 RCT, 1 OBS) | 0.75 (0.44, 1.14) | ⊕⊕⊕⊕<br>**HIGH** | Attending 1 round of PSA-based screening causes fewer than 2 per 1000 biopsy-related hospitalizations. |
| | Multiple rounds<br>N=19765 (1 RCT) | 2.23 (1.62, 2.99) | ⊕⊕⊕⊝<br>**MODERATE** [5]<br>due to reporting bias | Attending at least 1 round of PSA-based screening probably causes fewer than 3 per 1000 biopsy-related hospitalizations. |
| Any infective symptoms<br>(40 per 1000) | 1 round<br>N=44681 (2 RCTs, 1 OBS) | 4.12 (0.89, 9.65) | ⊕⊕⊕⊕<br>**HIGH** | Attending 1 round of PSA-based screening causes infective symptoms after biopsy in fewer than 11 per 1000 men |
| | Multiple rounds<br>N=19765 (1 RCT) | 14.0 (12.4, 15.7) | ⊕⊕⊕⊝<br>**MODERATE** [5]<br>due to reporting bias | Attending at least 1 round of PSA-based screening probably causes infective symptoms after biopsy in fewer than 16 per 1000 men |
| Major/severe infection<br>(20 per 1000) | 1 round<br>N=20273 (2 OBS) | 2.22 (1.60, 2.94) | ⊕⊕⊕⊝<br>**MODERATE** [7]<br>due to indirectness | Attending at least 1 round of PSA-based screening may cause major infection after biopsy in fewer than 3 per 1000 men |
| | Multiple rounds<br>N=35970 (1 RCT) | 0.81 (0.54, 1.13) | ⊕⊕⊝⊝<br>**LOW** [4, 5]<br>due to risk of bias and reporting bias | Attending at least 1 round of PSA-based screening may cause major infection after biopsy in fewer than 20 per 1000 men |
| Any bleeding complications<br>(80 per 1000) | 1 round<br>N=18922 (1 OBS) | 36.6 (34.0, 39.4) | ⊕⊕⊕⊝<br>**MODERATE** [5]<br>due to reporting bias | Attending 1 round of PSA-based screening probably causes bleeding complications after biopsy in fewer than 40 per 1000 men. |
| | Multiple rounds<br>N=19765 (1 RCT) | 180 (174, 185) | ⊕⊕⊕⊝<br>**MODERATE** [5]<br>due to reporting bias | Attending 1 or more rounds of PSA-based screening probably causes bleeding complications after biopsy in more than 174 per 1000 men. |
| Major/severe bleeding complications<br>(20 per 1000) | 1 round<br>N=18922 (1 OBS) | 3.22 (2.47, 4.14) | ⊕⊕⊕⊝<br>**MODERATE** [5]<br>due to reporting bias | Attending 1 round of PSA-based screening probably causes major bleeding complications after biopsy in fewer than 5 per 1000 men. |

| Outcome (Decision threshold) | No. participants (studies) | Cumulative incidence per 1000 (95 % CI) | Certainty of evidence | What happens? |
|---|---|---|---|---|
| | Multiple rounds<br>N=35870 (1 RCT) | 0.39 (0.21, 0.65) | ⊕⊕⊖⊖<br>**LOW** [4, 5]<br>due to risk of bias and reporting bias | Attending 1 round of PSA-based screening may cause major bleeding complications after biopsy in fewer than 40 per 1000 men. |
| Biopsy mortality (0.1 per 1000) | 1 round<br>N=120989 (9 RCTs, 1 OBS) | 0.00 (0.00, 0.00) per 10,000 | ⊕⊕⊕⊕<br>**HIGH** | Attending 1 round of PSA-based screening leads to fewer than 1 death after biopsy per 10,000 men invited. |
| | Multiple rounds<br>N=96073 (8 RCTs) | 0.00 (0.00, 0.00) per 10,000 | ⊕⊕⊕⊕<br>**HIGH** | Attending at least 1 round of PSA-based screening leads to fewer than 1 death after biopsy per 10,000 men invited. |

**Footnotes:**
1 Rated down for inconsistency around the threshold;
2 Not rated down for risk of bias because direction of bias in studies at high risk of bias is likely to underestimate events and thus is not a serious concern.
3 No eligible evidence for this outcome was identified.
4 Rated down for risk of bias because majority of the evidence came from studies assessed at high risk of bias.
5 We assume it was reasonable to expect studies to capture and report on all of this review's harms outcomes, therefore we had serious concerns about reporting bias and rated down outcomes not informed by at least two studies.
6 Majority of evidence came from study with indirectness from using primarily sextant biopsy, but the estimate is not close to the threshold so we do not have serious concerns and therefore did not rate down for indirectness.
7 Rated down for indirectness due to eligibility for biopsy within study limited to PSA <20 ng/ml.
8 Cumulative incidence of biopsy complications presented here is higher than cumulative incidence of false positives, so we had some concerns about inconsistency between outcomes. However, we did not rate down because we do not think the estimate would cross our threshold.

#### Within-study subgroup data on FPs and biopsy complications

| **Study**<br>Design<br>*sources* | **Age;**<br>Rounds;<br>Subgroups | **Outcome**<br>Analysis (PP, ITS)<br>*Study definition* | **Findings** |
|---|---|---|---|
| Race/Ethnicity | | | |
| **PLCO**<br>RCT<br>*Pinsky 2014* | 55-74y;<br>6 rounds;<br>Race/ethnicity | Any major/severe biopsy complication, multiple rounds<br>PP<br>*Any complication requiring medical intervention* | Black: 49.3/1,000 men biopsied (n=1,455)<br>Non-black: 19.1/1,000 men biopsied (n=34,415)<br><br>Black vs. non-black aOR (95% CI): 2.6 (1.2 to 5.9) |

| | | | |
|---|---|---|---|
| *Pinsky 2014* | 55-74y;<br>6 rounds;<br>Race/ethnicity | Major/severe infection, multiple rounds<br>PP<br>*Infectious complications requiring medical intervention* | Black: 42.3 per 1,000 men biopsied (n=1,455)<br>Non-black: 6.5 per 1,000 men biopsied (n=34,415)<br><br>Black vs. non-black aOR (95% CI): 7.1 (2.7 to 18) |
| *Miller 2018* | 54-74y;<br>6 rounds;<br>Race/ethnicity | False positive<br>PP<br>*No definition provided* | PSA only:<br>Black: 228/1,713<br>White: 3,915/33,043<br><br>PSA or DRE:<br>Black: 377/1,713<br>White: 7703/33,043 |
| **SABOR**<br>Observational<br>*Ankerst 2009* | Range: 28-80y;<br>2-6+ rounds;<br>Race/ethnicity | False positives<br>PP<br>*Men with 1+ benign biopsy* | White: 144/1,407<br>Black: 22/320<br>Hispanic: 54/835<br>Other: 1/16 |
| Family History | | | |
| **ERSPC Finland**<br>RCT<br>*Saarimäki 2015* | 55-69y;<br>3 rounds;<br>Family history (father or at least one brother diagnosed with PCa) | False positives<br>PP<br>*At least one positive PSA minus screen-detected PCa* | With FH: 262/1,723<br>Without: 2,578/21,033 |
| **SABOR**<br>Observational<br>*Ankerst 2009* | Range: 28-80y;<br>2-6+ rounds;<br>Family history (first degree family member with PCa) | False positives<br>PP<br>*Men with 1 or more negative biopsy* | With FH: 71/142<br>Without FH: 150/2,070 |

**Screening for prostate cancer using PSA with and without MRI: systematic reviews with meta-analysis**

Jennifer Pillay, Lindsay Gaudet, Sholeh Rahman, Roland Grad, Guylène Thériault, Philipp Dahm, Keith J Todd, Gail McCartney, Brett Thombs, Sabrina Saba, Lisa Hartling

#### Appendix 3. Additional information for KQ2

#### DRAFT

#### Contents

Study characteristics of studies in KQ2

Summary of risk of bias by outcome

Forest plots including subgroup analyses

**Screening for prostate cancer using PSA with and without MRI: systematic reviews with meta-analysis**

Jennifer Pillay, Lindsay Gaudet, Sholeh Rahman, Roland Grad, Guylène Thériault, Philipp Dahm, Keith J Todd, Gail McCartney, Brett Thombs, Sabrina Saba, Lisa Hartling

#### Study characteristics table for KQ2

| Study/trial | STHLM3-MRI | GÖTEBORG-2 trial | IP1-PROSTAGRAM | ERSPC Rotterdam, 5th screening round | GÖTEBORG trial, 10th screening round | ProstaPilot | PROBASE |
|---|---|---|---|---|---|---|---|
| **Study characteristics** | | | | | | | |
| Author, year | Eklund, 2021 | Hugosson, 2022 | Eldred-Evans, 2021 & 2023 | Alberts, 2018 | Grenabo Bergdahl, 2017 | Stanik, 2025 | Al-Monajjed, 2025 |
| Country | Sweden | Sweden | United Kingdom | Netherlands | Sweden | Czech Republic | Germany |
| Study design | RCT | RCT | Prospective cohort (within subject comparator) | Prospective cohort (within subject comparator) | Prospective cohort (within subject comparator) | Prospective cohort (within subject comparator) | Prospective cohort (within subject comparator) |
| Participants' allocation/randomization method | Computer-generated blocks of five, stratified into six Stockholm3 cancer-risk strata | Sequentially assignment by an external password-protected computer system, in a 1:1:1 ratio, stratified by y of birth | Volunteer participation | Group allocation determined by participants' consent | All men attending the 10th screening round | Volunteer participation | Random sample from population registries |
| Study setting | Community laboratories | Primary care facilities | Seven primary care practice and two imaging centers | ERSPC community settings in Rotterdam | Community-based | Local cancer institute and general practitioners' offices | Four local sites in Germany |
| Population eligibility | Men aged 50–74 from Stockholm County. This analysis included randomized participants with PSA ≥3 ng/ml, regardless of Stockholm3 results; men with a prior prostate biopsy >60 days before invitation or no prior biopsy were eligible | Men in the Swedish Population Register who were 50 to 60 y of age and were living in Gothenburg, Sweden, or its 10 surrounding municipalities in the 2015–2020 period | Men 50 to 69 y of age with a life expectancy of at least 10 y who had not undergone PSA testing or prostate MRI in the previous 2 years; had not had a urinary infection or prostatitis in the previous 6 months; and did not have a history of prostate biopsy, PCa, or any contraindication to MRI | Men aged 54–74 yr at enrollment with PSA≥3.0 ng/ml at the fifth screening (71–75 yr) round in the ERSPC Rotterdam trial. The MRI side study was initiated in January 2013 within the fifth (last) screening round of the trial | Men aged 50–64 y living in the city of Göteborg | Men aged 50-69 from the general population with life expectancy of at least 10 yr | 45-year-old men from the local population registries; needed to be eligible and consent (criteria NR)<br>(data also used for 50y olds screening after 5 y) |
| N invited/eligible | 49,118 | 25,850 | 2,034 | 825 | 596 | 432 | 46,642 |
| N screened (at baseline/the study's screening round) | 12,750 | 11,986 | 411 | 713 | 384 | 423 | 46,452 |
| **Sample characteristics** | | | | | | | |
| Age, median (IQR) | 61.0 (55.0-67.0) y | IG: 56.2 (52.9–59.8) y | 57.0 (53.0–61.0) y | 73.1 (72.3-74.0) y | 69.3 (69.0–69.6) y | 56.2 (NR) | Mean: 45.5 |

**Screening for prostate cancer using PSA with and without MRI: systematic reviews with meta-analysis**
Jennifer Pillay, Lindsay Gaudet, Sholeh Rahman, Roland Grad, Guylène Thériault, Philipp Dahm, Keith J Todd, Gail McCartney, Brett Thombs, Sabrina Saba, Lisa Hartling

| Study/trial | STHLM3-MRI | GÖTEBORG-2 trial | IP1-PROSTAGRAM | ERSPC Rotterdam, 5th screening round | GÖTEBORG trial, 10th screening round | ProstaPilot | PROBASE |
|---|---|---|---|---|---|---|---|
| | | CG: 56.0 (52.9–59.8) y | | | | *Mean: 57.2* | |
| Race/ethnicity | NR | Born in -<br>Sweden: 85%<br>Other European countries: 5%<br>Asia: 5%<br><br>*Majority were White, whereas Black, Hispanic and Asian men were underrepresented* | White: 38.0%<br>Black: 32.4%<br>Asian: 23.0%<br>Other: 4.4%<br>Mixed race: 2.2% | NR | NR | NR | Caucasian: 97.1%<br>Asian: 1.0%<br>African: 0.97%<br>Other: 0.95% |
| Personal &/or family history of PCa | NR | Family history of PCa: 10.5% | First-degree relative with PCa: 10.5%<br>Unknown: 1.2% | NR | NR | NR | Positive family history: 17% |
| Pre-trial screening | NR | 32% | 27% | NR | NR | PSA test within 2 yr before enrollment: 40%; 78% any previous PSA | PSA screened before age 45: 15.8%<br><br>DRE screened before age 45: 34.8% (37% more in delayed group by 5 years; with 2 cancers detected) |
| **MRI-based strategy (intervention)** | | | | | | | |
| Intervention | PSA + MRI, with targeted and systematic biopsy | PSA+ MRI with targeted biopsy | PSA+MRI followed by targeted biopsy | PSA+MRI ± fusion targeted biopsy | PSA+ MRI with targeted biopsy | PSA+ MRI with targeted biopsy | PSA+ MRI, with targeted and systematic biopsy |
| MRI type | Biparametric (i.e., combined T2- and diffusion-weighted imaging without contrast enhancement), without endorectal coil; image-fusion | Multiparametric, cognitive fusion, used until April 1, 2019; biparametric MRI used for all subsequent rounds of screening | Short MRI (T2-weighted and diffusion); image-fusion | Multiparametric; image-fusion | Multiparametric (mpMRI) | Biparametric (bpMRI), using 3-T MRI scanner | Multiparametric (mpMRI) |
| Indication for MRI (e.g. PSA positivity cutoff) | PSA ≥3 ng/ml | PSA ≥3 ng/ml | PSA ≥3 ng/ml | PSA ≥3 ng/ml | PSA ≥3 ng/ml | PSA ≥3 ng/ml | PSA ≥3 ng/ml |

| Study/trial | STHLM3-MRI | GÖTEBORG-2 trial | IP1-PROSTAGRAM | ERSPC Rotterdam, 5th screening round | GÖTEBORG trial, 10th screening round | ProstaPilot | PROBASE |
|---|---|---|---|---|---|---|---|
| MRI readings | By three uroradiologists; consensus by at least two radiologists required for each case | By two of four experienced prostate MRI radiologists in consensus, who were blinded to trial-group assignments, PSA levels, and clinical data. External validation of MRI readings was performed in a sample of 100 examinations | By two radiologists, one at each site, experienced in reporting prostate MRI scans. Scans were assessed using a scale of 1 to 5, with higher numbers indicating greater likelihood of clinically significant PCa | By three urogenital radiologists, each experienced in reading prostate MRI. Consensus on the PI-RADS score required for each identified lesion | All images were read in consensus by three radiologists of whom two had several years' experience of MRI-reading | Independently evaluated by two experienced uroradiologists blinded to PSA results. Discrepancies were resolved by a third radiologist. At study initiation, the radiologists each had 4 years' experience and had interpreted 400–1000 prostate MRI scans | Read by board-certified radiologists at the four sites with access to clinical data; no centralized pre-biopsy review was performed |
| Indication for biopsy | PI-RADS (v2.0–2.1) scores of ≥3. Men with PI-RADS ≤2 did not undergo biopsy unless at very high risk (Stockholm3 ≥25%) | PI-RADS (v2) scores of ≥3. If no suspicious lesions on MRI but PSA level of ≥10 ng/mL and a PSA density of ≥0.1 ng/mL, systematic biopsy was performed regardless of the MRI findings (only 0.5% of participants in the IG had PSA≥10 ng/ml) | PI-RADS (v2) scores of ≥3 | Presence of ≥1 PI-RADS ≥3 lesions | PI-RADS score in any sequence of ≥3 (equivocal) | PI-RADS (v2.1) of ≥4 | PI-RADS (v2.1) ≥3 |
| Biopsy type | Transrectal sampling of 3 to 4 biopsy cores in up to 3 clinically significant lesions , and a standard 10-to-12–core biopsy specimen immediately after the targeted biopsy | 4 cores obtained from MRI-positive lesions in biopsies directed by a cognitive MRI–transrectal-ultrasound fusion technique | Image-fusion targeted biopsies taken if the MRI or ultrasound was positive. Patients in whom a prostate lesion was found on MRI underwent targeted biopsy using cognitive co-registration, in addition to random 12-core biopsy. A maximum of 4 cores | 12-core TRUS biopsy blinded for MRI findings. One additional core was taken from each hypoechoic lesion. Targeted biopsy with 2 cores per lesion | Three additional cores (in addition to 10-core TRUS) sampled per suspicious region by means of "cognitive" targeting | 12-core TRUS-guided biopsy from the peripheral zone of the prostate and additional three core (NR if all lesions) MRI/TRUS fusion biopsy in MRI positive cases | mpMRI/ultrasound fusion biopsy of target lesions (maximum of 3 with 2 cores per target) followed by 12 to 18 systematic biopsies depending on the volume of the prostate. Biopsies were conducted transrectally or transperineally |

| Study/trial | STHLM3-MRI | GÖTEBORG-2 trial | IP1-PROSTAGRAM | ERSPC Rotterdam, 5th screening round | GÖTEBORG trial, 10th screening round | ProstaPilot | PROBASE |
|---|---|---|---|---|---|---|---|
| | | | was allowed for the primary target | | | | |
| Biopsy readings | By one of four experienced uropathologists, reporting GS and cancer length per core per ISUP 2014 guidelines. Overall GS were reported per method; combined biopsy scores reflected the highest score | Initially assessed by an experienced pathologist. Those showing cancer were independently reviewed by two external uropathologists, blinded to group assignment and clinical data | All specimens centrally reviewed by expert uropathologists according to the ISUP guidelines | Grading by one expert uropathologist according to the ISUP 2014 GS | NR | All specimens were reviewed by an expert uropathologist | Histopathological analysis of the cores was conducted locally and was reviewed centrally by an experienced genitourinary pathologist with 20 y of experience |
| **PSA-based strategy (comparator)** | | | | | | | |
| Indication for biopsy | PSA ≥3 ng/ml | PSA ≥3 ng/ml | PSA ≥3 ng/ml | PSA ≥3 ng/ml | PSA ≥3 ng/ml | PSA ≥3 ng/ml | PSA ≥3 ng/ml (confirmed with repeat PSA tests) |
| Biopsy type | TRUS-guided, 10 to 12 cores obtained from the peripheral zone of the prostate (apical, mid-gland, and base) | TRUS-guided; 10 to 12 cores obtained from the peripheral zone | Transperineal 12 core systematic biopsy. All biopsies were performed using a biplanar TRUS machine | TRUS-guided, 12-core biopsy blinded for MRI findings. One additional core was taken from each hypoechoic lesion | TRUS-guided, 10 core was sampled first, blinded to MRI results, using a scheme with 12 anterior and 12 posterior sectors of which 10 posterior were sampled routinely. The MRI results were revealed after the systematic biopsy | TRUS-guided systematic 12-core biopsy from the peripheral zone of the prostate | Transrectal or transperineal systematic 12 to 18 core biopsy. A scheme according to the size of the prostate was used, which comprised 12 independent systematic cores in most cases |
| Biopsy reading | Same as in the intervention group | Same as in the intervention group | Same as in the intervention group. Reporting radiologists were not present during the biopsy to ensure the reference test remained independent of the index test | Same as in the intervention group | NR | All specimens were reviewed by an expert uropathologist | Histopathological analysis of the cores was conducted locally and was reviewed centrally by an experienced genitourinary pathologist with 20 y of experience |

CG: control group; E: European Randomized study of Screening for Prostate Cancer; GS: Gleason score; IG: intervention group; ISUP: International Society of Urological Pathology; NR: not reported; PCa: prostate cancer; PSA: prostate specific antigen; RCT: randomized controlled trial; TRUS: transrectal ultrasonography

**Screening for prostate cancer using PSA with and without MRI: systematic reviews with meta-analysis**
Jennifer Pillay, Lindsay Gaudet, Sholeh Rahman, Roland Grad, Guylène Thériault, Philipp Dahm, Keith J Todd, Gail McCartney, Brett Thombs, Sabrina Saba, Lisa Hartling

#### Summary of risk of bias by outcome (QUADAS-C)

| Study<br>Outcome | Biopsy type in each arm (i.e., sequential screening and PSA-based screening) | Risk of bias (QUADAS-2) P | I | R | FT | O | Applicability concerns (QUADAS-2) P | I | R | Risk of bias (QUADAS-C) P | I | R | FT | O | Overall RoB |
|---|---|---|---|---|---|---|---|---|---|---|---|---|---|---|---|
| **Clinically significant PCa** | | | | | | | | | | | | | | | |
| Alberts, 2018 | MRI TBx | ✓ | ✓ | ? | ? | NA | ? | ✓ | ✓ | ✓ | ✓ | ? | ✓ | NA | Some concern |
| Clinically significant PCa, PI-RADS≥3 | 12-core TRUS-Bx | ✓ | ✓ | ? | ? | NA | | | | | | | | | |
| Eklund, 2021 | MRI TBx | ✓ | ✓ | ? | ? | NA | ✓ | ✓ | ✓ | ✓ | ✓ | ? | ✗ | NA | High risk |
| Clinically significant PCa, PI-RADS≥3 | 10-12 core TRUS bx | ✓ | ✓ | ? | ✗ | NA | | | | | | | | | |
| Grenabo Bergdahl, 2017 | MRI TBx | ✓ | ✓ | ? | ? | NA | ? | ✓ | ✓ | ✓ | ✓ | ✓ | ? | NA | Some concern |
| Clinically significant PCa, PI-RADS≥3 | 10-core TRUS systematic bx | ✓ | ✓ | ? | ? | NA | | | | | | | | | |
| Hugosson, 2022 | MRI TBx | ✓ | ✓ | ✓ | ? | NA | ? | ✓ | ✓ | ✓ | ✓ | ✓ | ? | NA | Some concern |
| Clinically significant PCa, PI-RADS≥3 | 10-12 core TRUS systematic bx | ✓ | ✓ | ✓ | ? | NA | | | | | | | | | |
| Eldred-Evans, 2023 | MRI TBx | ✗ | ? | ? | ✓ | NA | ? | ? | ✓ | ✓ | ? | ? | ✓ | NA | Some concern |
| Clinically significant PCa, PI-RADS≥3 | 12-core systematic bx | ✗ | ✓ | ✓ | ✓ | NA | | | | | | | | | |
| **Clinically significant PCa, subgroups** | | | | | | | | | | | | | | | |
| Alberts, 2018 | MRI TBx | ✓ | ✓ | ? | ? | NA | ? | ✓ | ✓ | ✓ | ✓ | ? | ✓ | NA | Some concern |
| Clinically significant PCa, PI-RADS≥4 | 12-core TRUS-Bx | ✓ | ✓ | ? | ? | NA | | | | | | | | | |
| Eldred-Evans, 2023 | MRI TBx | ✗ | ? | ? | ✓ | NA | ? | ? | ✓ | ✓ | ? | ? | ✓ | NA | Some concern |
| Clinically significant PCa, PI-RADS≥4 | 12-core systematic bx | ✗ | ✓ | ✓ | ✓ | NA | | | | | | | | | |
| Stanik, 2025 | MRI TBx | ✗ | ✓ | ? | ? | NA | ? | ✓ | ✓ | ✓ | ✓ | ✓ | ✗ | NA | High risk |
| Clinically significant PCa, PI-RADS≥4 | 12-core systematic bx | ✗ | ✓ | ? | ? | NA | | | | | | | | | |

| Study<br>Outcome | Biopsy type in each arm (i.e., sequential screening and PSA-based screening) | Risk of bias (QUADAS-2) P | I | R | FT | O | Applicability concerns (QUADAS-2) P | I | R | Risk of bias (QUADAS-C) P | I | R | FT | O | Overall RoB |
|---|---|---|---|---|---|---|---|---|---|---|---|---|---|---|---|
| Alberts, 2018 | MRI TBx and 12-core TRUS-Bx | ✓ | ✓ | ? | ? | NA | ? | ✓ | ✓ | ✓ | ✓ | ? | ✓ | NA | Some concern |
| Clinically significant PCa, PI-RADS≥3 | 12-core TRUS-Bx | ✓ | ✓ | ? | ? | NA | | | | | | | | | |
| Eklund, 2021 | MRI TBx and 10-12-core TRUS-Bx | ✓ | ✓ | ? | ? | NA | ✓ | ✓ | ✓ | ✓ | ✓ | ? | ✗ | NA | High risk |
| Clinically significant PCa, PI-RADS≥3 | 10-12 core TRUS bx | ✓ | ✓ | ? | ✗ | NA | | | | | | | | | |
| Eldred-Evans, 2021 | MRI TBx and 12-core systematic bx | ✗ | ? | ? | ✓ | NA | ? | ? | ? | ✓ | ? | ✓ | ✓ | NA | Some concern |
| Clinically significant PCa, PI-RADS≥3 | 12-core systematic bx | ✗ | ✓ | ✓ | ✓ | NA | | | | | | | | | |
| Al-Monajjed, 2025 | MRI TBx and 12-18 core systematic bx | ? | ✗ | ? | ✗ | NA | ✓ | ✓ | ✓ | ✓ | ✓ | ✓ | ✗ | NA | High risk |
| Clinically significant PCa, PI-RADS≥3 | 12-18 core systematic bx | ? | ✓ | ? | ✗ | NA | | | | | | | | | |
| **Clinically insignificant PCa** | | | | | | | | | | | | | | | |
| Alberts, 2018 | MRI TBx | ✓ | ✓ | ? | ? | NA | ? | ✓ | ✓ | ✓ | ✓ | ? | ✓ | NA | Some concern |
| Clinically insignificant PCa, PI-RADS≥3 | 12-core TRUS-Bx | ✓ | ✓ | ? | ? | NA | | | | | | | | | |
| Eklund, 2021 | MRI TBx | ✓ | ✓ | ? | ? | NA | ✓ | ✓ | ✓ | ✓ | ✓ | ? | ✗ | NA | High risk |
| Clinically insignificant PCa, PI-RADS≥3 | 10-12 core TRUS bx | ✓ | ✓ | ? | ✗ | NA | | | | | | | | | |
| Eldred-Evans, 2023 | MRI TBx | ✗ | ? | ? | ✓ | NA | ? | ? | ✓ | ✓ | ? | ? | ✓ | NA | Some concern |
| Clinically insignificant PCa, PI-RADS≥3 | 12-core systematic bx | ✗ | ✓ | ✓ | ✓ | NA | | | | | | | | | |
| Grenabo Bergdahl, 2017 | MRI TBx | ✓ | ✓ | ? | ? | NA | ? | ✓ | ✓ | ✓ | ✓ | ✓ | ? | NA | Some concern |
| Clinically insignificant PCa, PI-RADS≥3 | 10-core TRUS systematic bx | ✓ | ✓ | ? | ? | NA | | | | | | | | | |
| Hugosson, 2022 | MRI TBx | ✓ | ✓ | ✓ | ? | NA | ? | ✓ | ✓ | ✓ | ✓ | ✓ | ? | NA | Some concern |

| Study<br>Outcome | Biopsy type in each arm (i.e., sequential screening and PSA-based screening) | Risk of bias (QUADAS-2) | | | | | Applicability concerns (QUADAS-2) | | | Risk of bias (QUADAS-C) | | | | | Overall RoB |
|---|---|---|---|---|---|---|---|---|---|---|---|---|---|---|---|
| | | P | I | R | FT | O | P | I | R | P | I | R | FT | O | |
| Clinically insignificant PCa, PI-RADS≥3 | 10-12 core TRUS systematic bx | ✓ | ✓ | ✓ | ? | NA | | | | | | | | | |
| **Clinically insignificant PCa, subgroups** | | | | | | | | | | | | | | | |
| Alberts, 2018 | MRI TBx | ✓ | ✓ | ? | ? | NA | ? | ✓ | ✓ | ✓ | ✓ | ? | ✓ | NA | Some concern |
| Clinically insignificant PCa, PI-RADS≥4 | 12-core TRUS-Bx | ✓ | ✓ | ? | ? | NA | | | | | | | | | |
| Eldred-Evans, 2023 | MRI TBx | ✗ | ? | ? | ✓ | NA | ? | ? | ✓ | ✓ | ? | ? | ✓ | NA | Some concern |
| Clinically insignificant PCa, PI-RADS≥4 | 12-core systematic bx | ✗ | ✓ | ✓ | ✓ | NA | | | | | | | | | |
| Stanik, 2025 | MRI TBx | ✗ | ✓ | ? | ? | NA | ? | ✓ | ✓ | ✓ | ✓ | ✓ | ✗ | NA | High risk |
| Clinically insignificant PCa, PI-RADS≥4 | 12-core systematic bx | ✗ | ✓ | ? | ? | NA | | | | | | | | | |
| Alberts, 2018 | MRI TBx and 12-core TRUS-Bx | ✓ | ✓ | ? | ? | NA | ? | ✓ | ✓ | ✓ | ✓ | ? | ✓ | NA | Some concern |
| Clinically insignificant PCa, PI-RADS≥3 | 12-core TRUS-Bx | ✓ | ✓ | ? | ? | NA | | | | | | | | | |
| Eklund, 2021 | MRI TBx and 10-12-core TRUS-Bx | ✓ | ✓ | ? | ? | NA | ✓ | ✓ | ✓ | ✓ | ✓ | ? | ✗ | NA | High risk |
| Clinically insignificant PCa, PI-RADS≥3 | 10-12 core TRUS bx | ✓ | ✓ | ? | ✗ | NA | | | | | | | | | |
| Eldred-Evans, 2021 | MRI TBx and 12-core systematic bx | ✗ | ? | ✓ | ✓ | NA | ? | ? | ? | ✓ | ? | ✓ | ✓ | NA | Some concern |
| Clinically insignificant PCa, PI-RADS≥3 | 12-core systematic bx | ✗ | ✓ | ✓ | ✓ | NA | | | | | | | | | |
| Al-Monajjed, 2025 | MRI TBx and 12-18 core systematic bx | ? | ✗ | ? | ✗ | NA | ✓ | ✓ | ✓ | ✓ | ✓ | ✓ | ✗ | NA | High risk |
| Clinically insignificant PCa, PI-RADS≥3 | 12-18 core systematic bx | ? | ✓ | ? | ✗ | NA | | | | | | | | | |
| **False positive** | | | | | | | | | | | | | | | |
| Alberts, 2018 | MRI TBx | ✓ | ✓ | ? | ? | NA | ? | ✓ | ✓ | ✓ | ✓ | ? | ✓ | NA | Some concern |
| False positive, PI-RADS≥3 | 12-core TRUS-Bx | ✓ | ✓ | ? | ? | NA | | | | | | | | | |
| Eklund, 2021 | MRI TBx | ✓ | ✓ | ? | ? | NA | ✓ | ✓ | ✓ | ✓ | ✓ | ? | ✗ | NA | High risk |

| Study<br>Outcome | Biopsy type in each arm (i.e., sequential screening and PSA-based screening) | Risk of bias (QUADAS-2) | | | | | Applicability concerns (QUADAS-2) | | | Risk of bias (QUADAS-C) | | | | | Overall RoB |
|---|---|---|---|---|---|---|---|---|---|---|---|---|---|---|---|
| | | P | I | R | FT | O | P | I | R | P | I | R | FT | O | |
| False positive, PI-RADS≥3 | 10-12 core TRUS bx | ✓ | ✓ | ? | ✗ | NA | | | | | | | | | |
| Eldred-Evans, 2023 | MRI TBx | ✗ | ? | ? | ✓ | NA | ? | ? | ✓ | ✓ | ? | ? | ✓ | NA | Some concern |
| False positive, PI-RADS≥3 | 12-core systematic bx | ✗ | ✓ | ✓ | ✓ | NA | | | | | | | | | |
| Grenabo Bergdahl, 2017 | MRI TBx | ✓ | ✓ | ? | ? | NA | ? | ✓ | ✓ | ✓ | ✓ | ? | ? | NA | Some concern |
| False positive, PI-RADS≥3 | 10-core TRUS systematic bx | ✓ | ✓ | ? | ? | NA | | | | | | | | | |
| Hugosson, 2022 | MRI TBx | ✓ | ✓ | ✓ | ✓ | NA | ? | ✓ | ✓ | ✓ | ✓ | ✓ | ✗ | NA | High risk |
| False positive, PI-RADS≥3 | 10-12 core TRUS systematic bx | ✓ | ✓ | ✓ | ? | NA | | | | | | | | | |
| **False positive, subgroups** | | | | | | | | | | | | | | | |
| Alberts, 2018 | MRI TBx | ✓ | ✓ | ? | ? | NA | ? | ✓ | ✓ | ✓ | ✓ | ? | ✓ | NA | Some concern |
| False positive, PI-RADS≥4 | 12-core TRUS-Bx | ✓ | ✓ | ? | ? | NA | | | | | | | | | |
| Stanik, 2025 | MRI TBx | ✗ | ✓ | ? | ? | NA | ? | ✓ | ✓ | ✓ | ✓ | ✓ | ✗ | NA | High risk |
| False positive, PI-RADS≥4 | 12-core systematic bx | ✗ | ✓ | ? | ? | NA | | | | | | | | | |
| Alberts, 2018 | MRI TBx and 12-core TRUS-Bx | ✓ | ✓ | ? | ? | NA | ? | ✓ | ✓ | ✓ | ✓ | ? | ✓ | NA | Some concern |
| False positive, PI-RADS≥3 | 12-core TRUS-Bx | ✓ | ✓ | ? | ? | NA | | | | | | | | | |
| Eklund, 2021 | MRI TBx and 10-12-core TRUS-Bx | ✓ | ✓ | ? | ? | NA | ✓ | ✓ | ✓ | ✓ | ✓ | ? | ✗ | NA | High risk |
| False positive, PI-RADS≥3 | 10-12 core TRUS bx | ✓ | ✓ | ? | ✗ | NA | | | | | | | | | |
| Eldred-Evans, 2021 | MRI TBx and 12-core systematic bx | ✗ | ? | ✓ | ✓ | NA | ? | ? | ? | ✓ | ? | ✓ | ✓ | NA | Some concern |
| False positive, PI-RADS≥3 | 12-core systematic bx | ✗ | ✓ | ✓ | ✓ | NA | | | | | | | | | |
| Al-Monajjed, 2025 | MRI TBx and 12-18 core systematic bx | ? | ✗ | ? | ✗ | NA | ✓ | ✓ | ✓ | ✓ | ✓ | ✓ | ✗ | NA | High risk |
| False positive, PI-RADS≥3 | 12-18 core systematic bx | ? | ✓ | ? | ✗ | NA | | | | | | | | | |
| **Complications: Infection** | | | | | | | | | | | | | | | |

**Screening for prostate cancer using PSA with and without MRI: systematic reviews with meta-analysis**

Jennifer Pillay, Lindsay Gaudet, Sholeh Rahman, Roland Grad, Guylène Thériault, Philipp Dahm, Keith J Todd, Gail McCartney, Brett Thombs, Sabrina Saba, Lisa Hartling

| Study<br>Outcome | Biopsy type in each arm (i.e., sequential screening and PSA-based screening) | Risk of bias (QUADAS-2) | | | | | Applicability concerns (QUADAS-2) | | | Risk of bias (QUADAS-C) | | | | | Overall RoB |
|---|---|---|---|---|---|---|---|---|---|---|---|---|---|---|---|
| | | P | I | R | FT | O | P | I | R | P | I | R | FT | O | |
| Eklund, 2021<br>Complications: Infection | MRI TBx and 10-12-core TRUS-Bx | ✓ | NA | NA | NA | ✓ | ✓ | NA | ✓ | ✓ | NA | NA | NA | ✓ | Low risk |
| | 10-12 core TRUS bx | ✓ | NA | NA | NA | ✓ | | | | | | | | | |
| Hugosson, 2022<br>Complications: Infection | MRI TBx and 10-12 core systematic bx | ✓ | NA | NA | NA | ✓ | ? | NA | ✓ | ✓ | NA | NA | NA | ✓ | Low risk |
| | 10-12 core TRUS systematic bx | ✓ | NA | NA | NA | ✓ | | | | | | | | | |
| Stanik, 2025<br>Complications: infection, serious adverse events, mortality* | MRI TBx | ✗ | ✓ | NA | NA | ? | ? | ✓ | ✓ | ✓ | ✓ | NA | NA | ✓ | High risk |
| | 12-core systematic bx | ✗ | ✓ | NA | NA | ? | | | | | | | | | |
| **Complications: Serious adverse events (AEs)** | | | | | | | | | | | | | | | |
| Eklund, 2021<br>Complications: Serious AEs (i.e., hospitalization) | MRI TBx and 10-12-core TRUS-Bx | ✓ | NA | NA | NA | ✓ | ✓ | NA | ✓ | ✓ | NA | NA | NA | ✓ | Low risk |
| | 10-12 core TRUS bx | ✓ | NA | NA | NA | ✓ | | | | | | | | | |
| Eldred-Evans, 2021<br>Complications: Serious AEs | MRI TBx and 12-core systematic bx | ✗ | NA | NA | NA | ? | ? | NA | ? | ✓ | NA | NA | NA | ✓ | Low risk |
| | 12-core systematic bx | ✗ | NA | NA | NA | ? | | | | | | | | | |
| Hugosson, 2022<br>Complications: Serious AEs (i.e., hospitalization) | MRI TBx and 10-12 core systematic bx | ✓ | NA | NA | NA | ✓ | ? | NA | ✓ | ✓ | NA | NA | NA | ✓ | Low risk |
| | 10-12 core TRUS systematic bx | ✓ | NA | NA | NA | ✓ | | | | | | | | | |
| **Complications: Mortality** | | | | | | | | | | | | | | | |
| Eklund, 2021<br>Complications: mortality | MRI TBx and 10-12-core TRUS-Bx | ✓ | NA | NA | NA | ✓ | ✓ | NA | ✓ | ✓ | NA | NA | NA | ✓ | Low risk |
| | 10-12 core TRUS bx | ✓ | NA | NA | NA | ✓ | | | | | | | | | |
| Hugosson, 2022<br>Complications: mortality | MRI TBx and 10-12 core systematic bx | ✓ | NA | NA | NA | ✓ | ? | NA | ✓ | ✓ | NA | NA | NA | ✓ | Low risk |

**Screening for prostate cancer using PSA with and without MRI: systematic reviews with meta-analysis**
Jennifer Pillay, Lindsay Gaudet, Sholeh Rahman, Roland Grad, Guylène Thériault, Philipp Dahm, Keith J Todd, Gail McCartney, Brett Thombs, Sabrina Saba, Lisa Hartling

| Study / Outcome | Biopsy type in each arm (i.e., sequential screening and PSA-based screening) | Risk of bias (QUADAS-2) | | | | | Applicability concerns (QUADAS-2) | | | Risk of bias (QUADAS-C) | | | | | Overall RoB |
|---|---|---|---|---|---|---|---|---|---|---|---|---|---|---|---|
| | | P | I | R | FT | O | P | I | R | P | I | R | FT | O | |
| | 10-12 core TRUS systematic bx | ✓ | NA | NA | NA | ✓ | | | | | | | | | |

P = patient selection; I = index test; R = reference standard; FT = flow and timing; O = outcome definition (this domain was only rated for complications); Bx = biopsy; TBx = targeted biopsy; TRUS = transrectal ultrasound; NA = not applicable; PCa = prostate cancer; RoB = risk of bias
✓ indicates low risk, ✗ indicates high risk, and ? indicates unclear risk in the risk of bias sections. ✓ indicates no concern, and ? indicates some concern in the applicability concerns section.
* The review team assumed that "adverse events" reported by the study authors applied to all three categories of complications in our review.

#### Forest plots for intention-to-screen (ITS) and per protocol (PP) effects

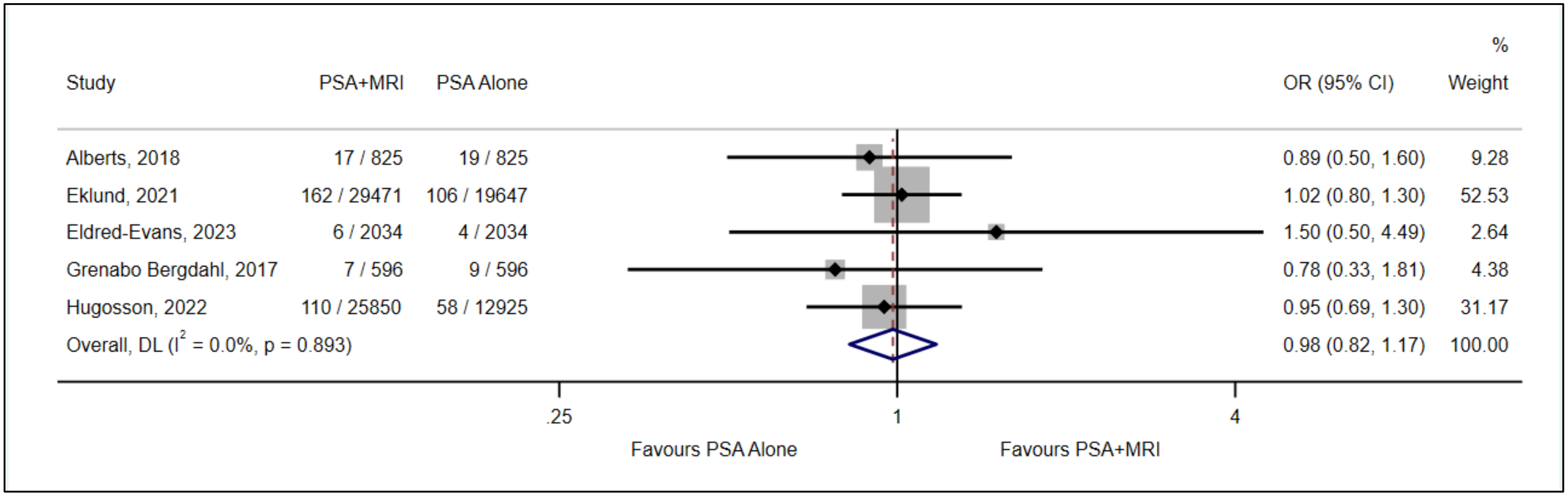


**Fig 1.** Clinically significant prostate cancer, comparison between MRI followed by targeted biopsy after PSA-based screening versus PSA-based screening alone, based on PI-RADS≥3 MRI cutoff for cancer detection (ITT)

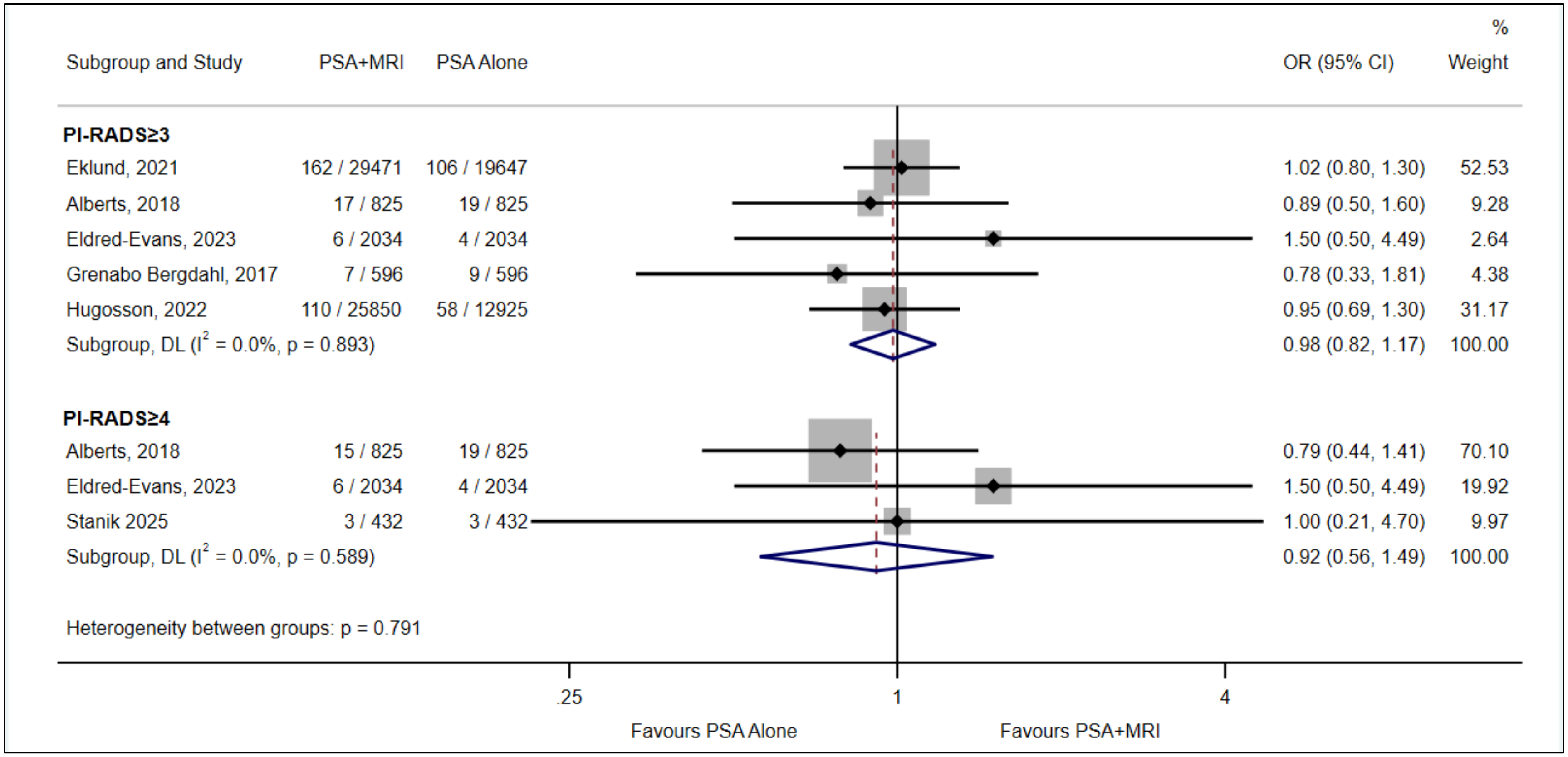


**Fig 2.** Clinically significant prostate cancer, comparison between MRI followed by targeted biopsy after PSA-based screening versus PSA-based screening alone, subgroup analysis based on PI-RADS thresholds (ITT)

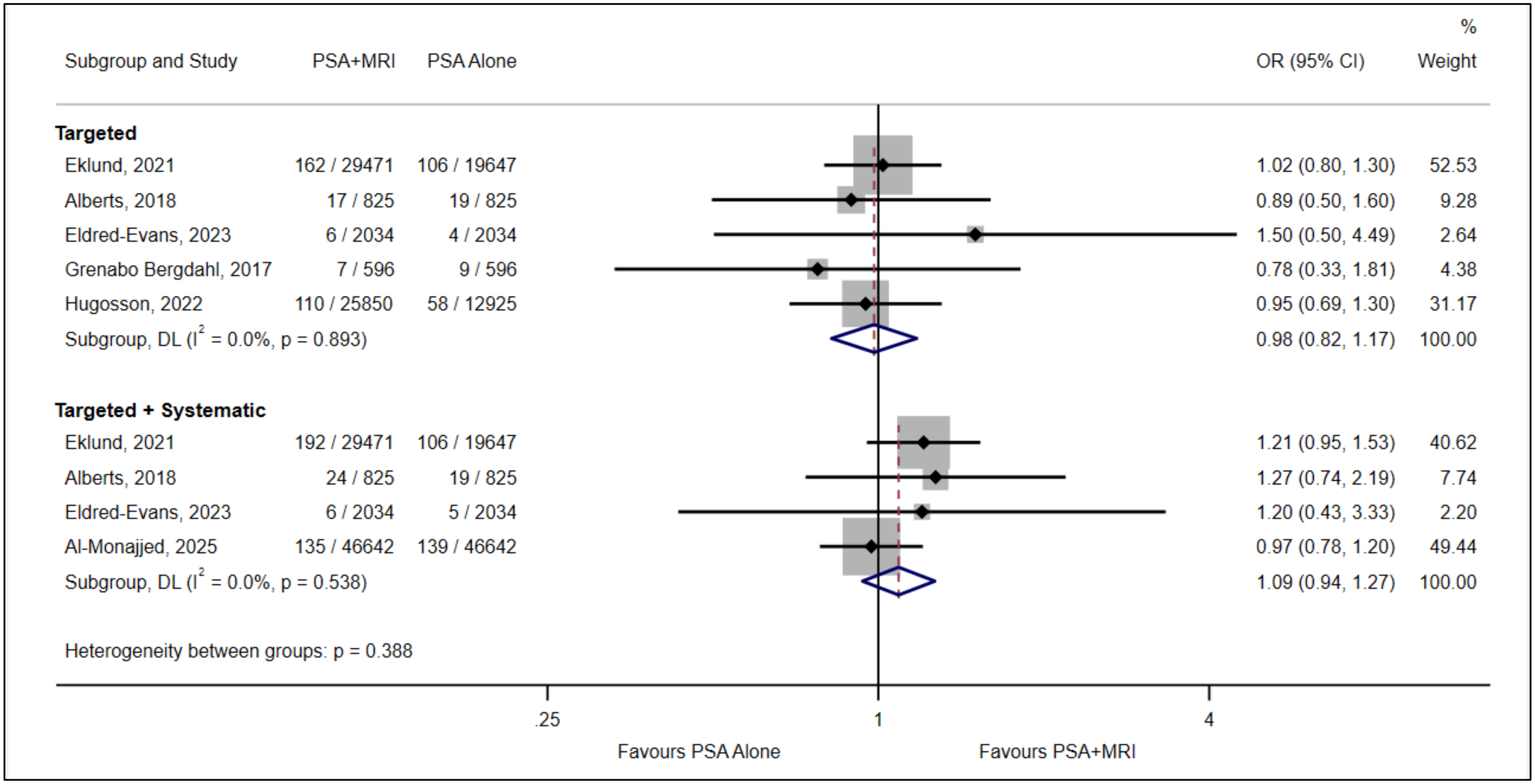


**Fig 3.** Clinically significant prostate cancer, subgroup analysis for PI-RADS≥3 MRI cutoff based on comprehensiveness of biopsy in the MRI arm (ITT)

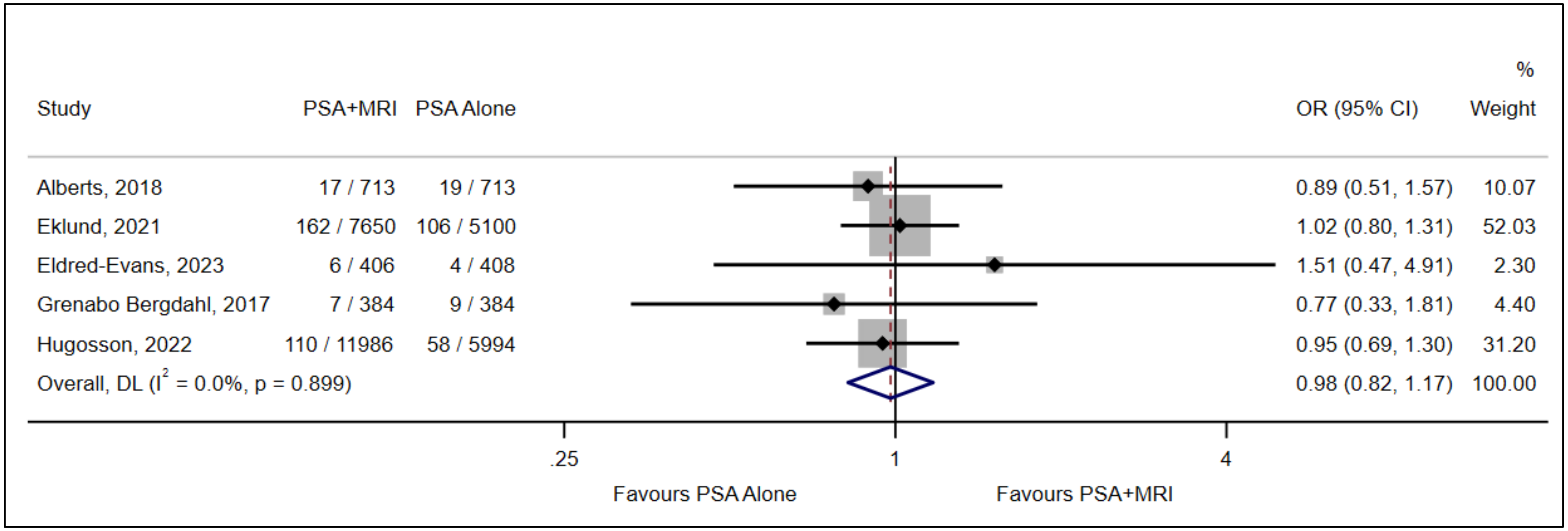


**Fig 4.** Clinically significant prostate cancer, comparison between MRI followed by targeted biopsy after PSA-based screening versus PSA-based screening alone, based on PI-RADS≥3 MRI cutoff for cancer detection (PP)

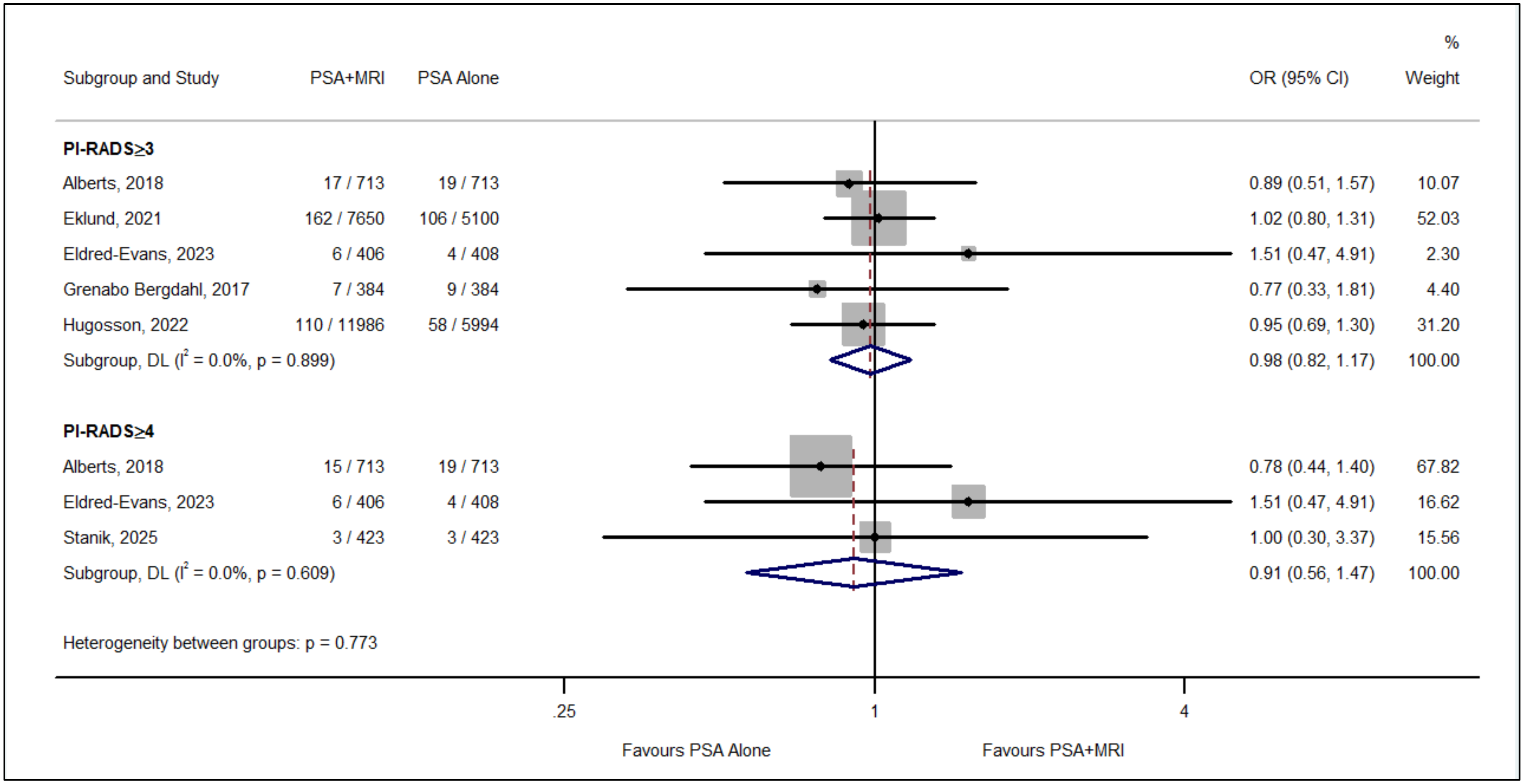


**Fig 5.** Clinically significant prostate cancer, comparison between MRI followed by targeted biopsy after PSA-based screening versus PSA-based screening alone, subgroup analysis based on PI-RADS thresholds (PP)

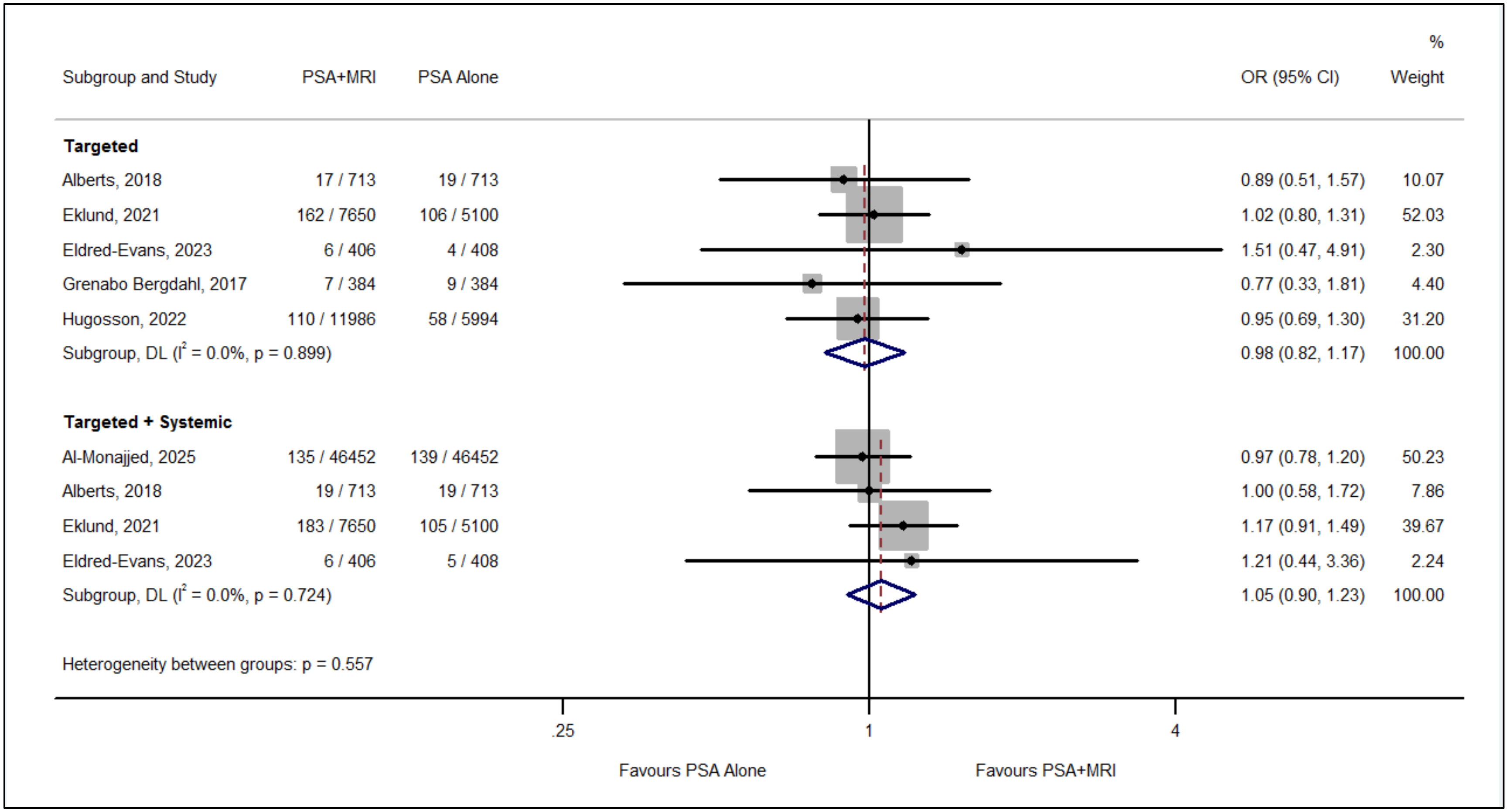


**Fig 6.** Clinically significant prostate cancer, subgroup analysis for PI-RADS≥3 MRI cutoff based on comprehensiveness of biopsy in the MRI arm (PP)

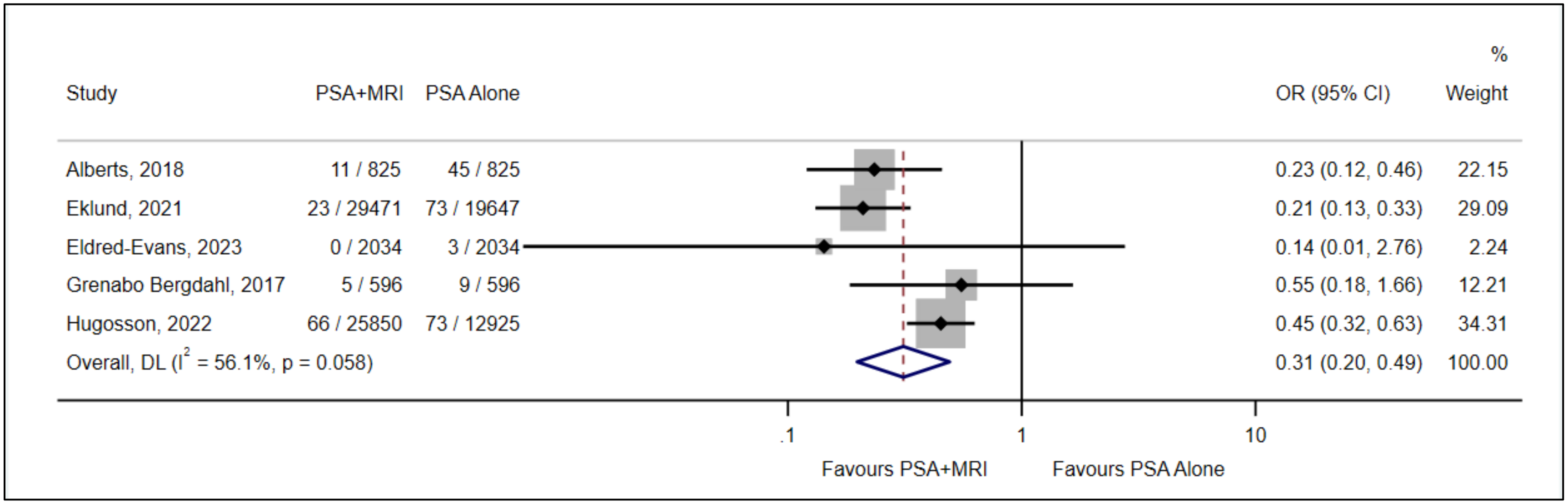


**Fig 7.** Clinically insignificant prostate cancer, comparison between MRI followed by targeted biopsy after PSA-based screening versus PSA-based screening alone, based on PI-RADS≥3 MRI cutoff for cancer detection (ITT)

**Screening for prostate cancer using PSA with and without MRI: systematic reviews with meta-analysis**
Jennifer Pillay, Lindsay Gaudet, Sholeh Rahman, Roland Grad, Guylène Thériault, Philipp Dahm, Keith J Todd, Gail McCartney, Brett Thombs, Sabrina Saba, Lisa Hartling

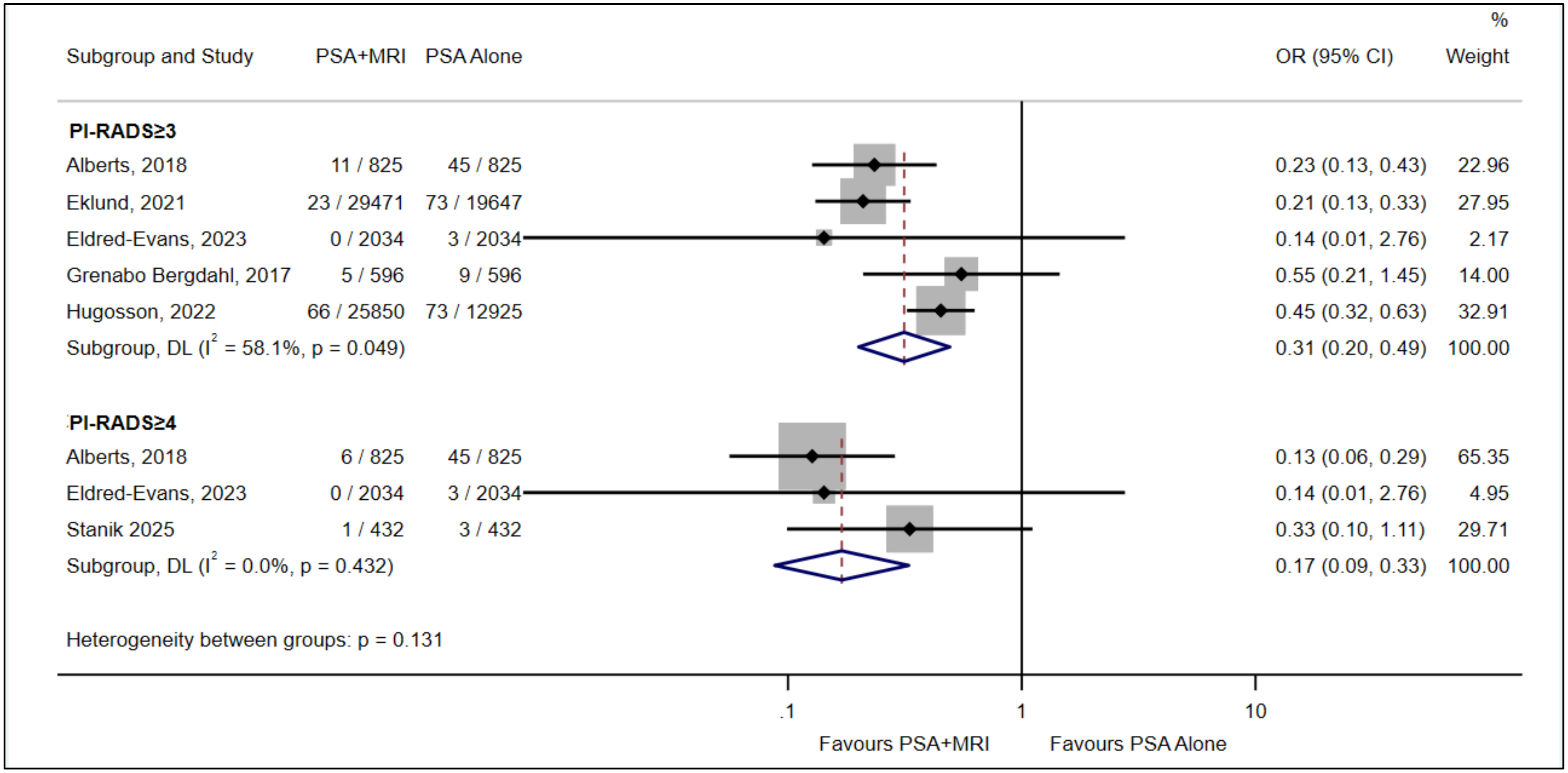


**Fig 8.** Clinically insignificant prostate cancer, comparison between MRI followed by targeted biopsy after PSA-based screening versus PSA-based screening alone, subgroup analysis based on PI-RADS thresholds (ITT)

**Screening for prostate cancer using PSA with and without MRI: systematic reviews with meta-analysis**
Jennifer Pillay, Lindsay Gaudet, Sholeh Rahman, Roland Grad, Guylène Thériault, Philipp Dahm, Keith J Todd, Gail McCartney, Brett Thombs, Sabrina Saba, Lisa Hartling

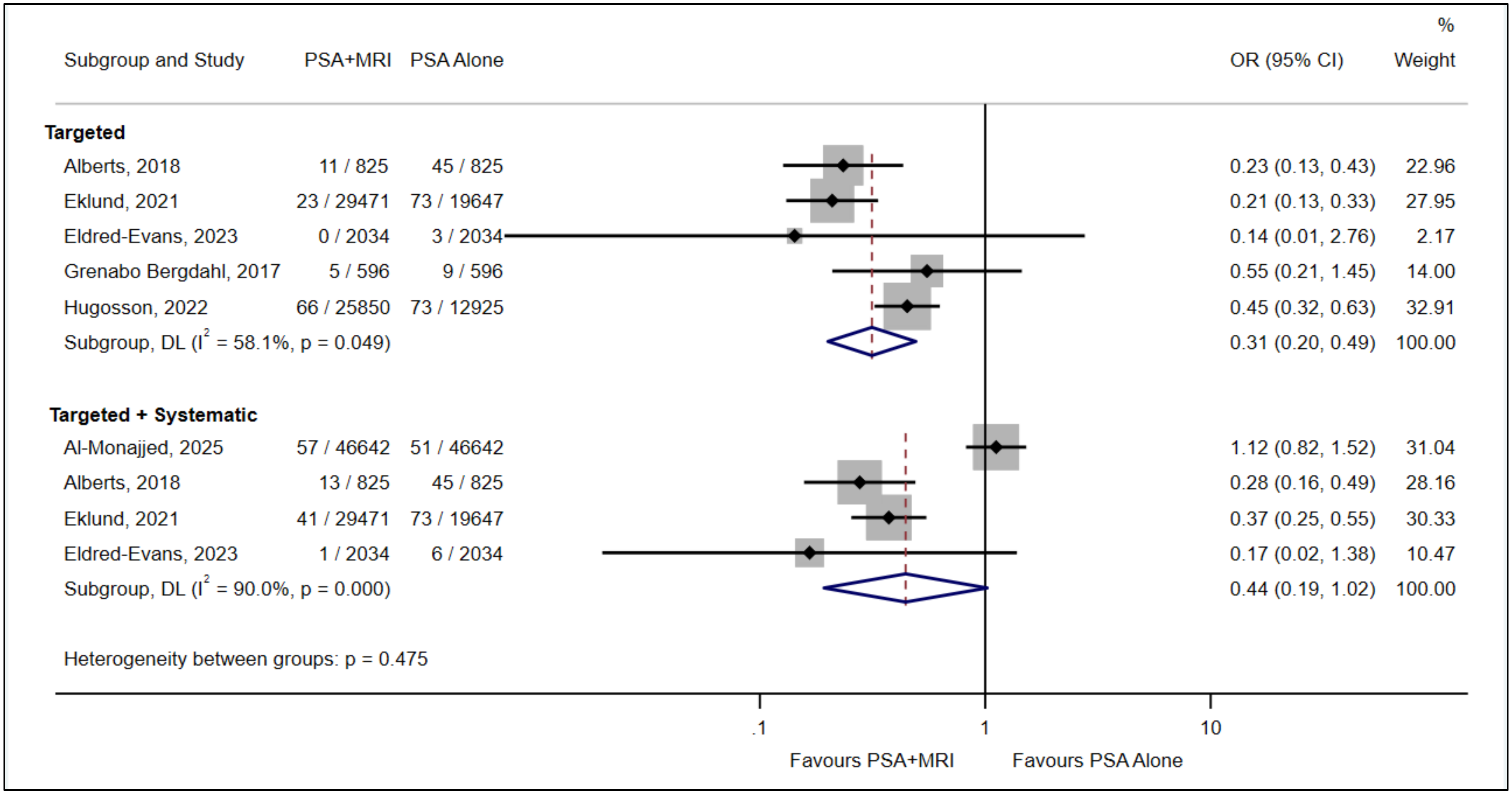


**Fig 9.** Clinically insignificant prostate cancer, subgroup analysis for PI-RADS≥3 MRI cutoff based on comprehensiveness of biopsy in the MRI arm (ITT)

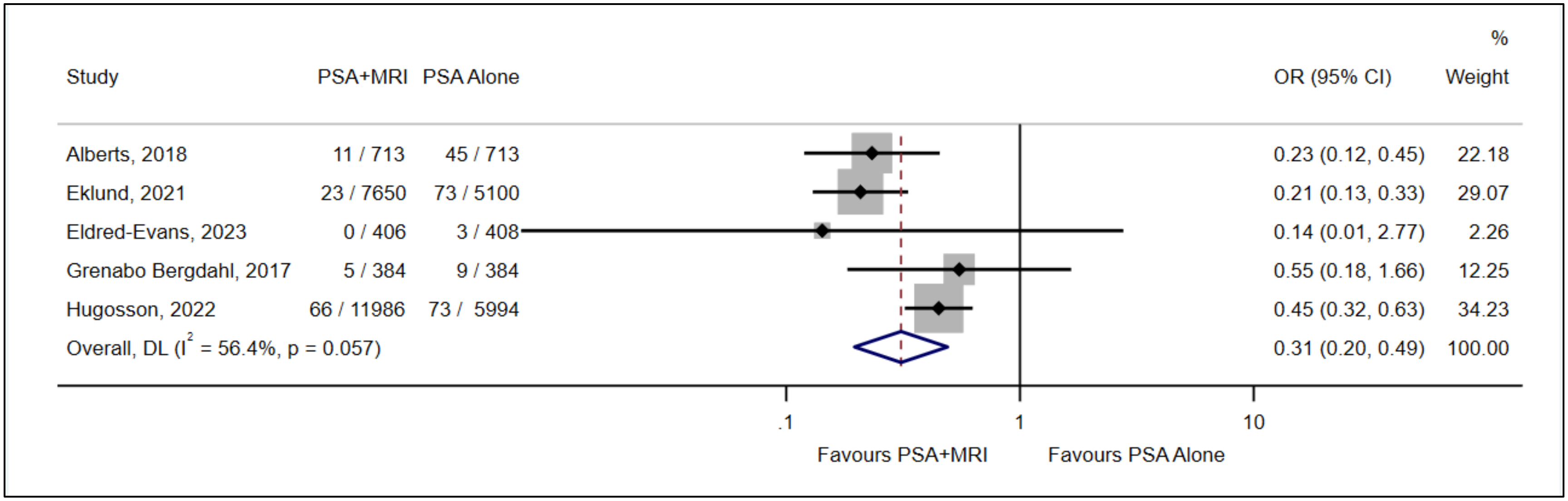


**Fig 10.** Clinically insignificant prostate cancer, comparison between MRI followed by targeted biopsy after PSA-based screening versus PSA-based screening alone, based on PI-RADS≥3 MRI cutoff for cancer detection (PP)

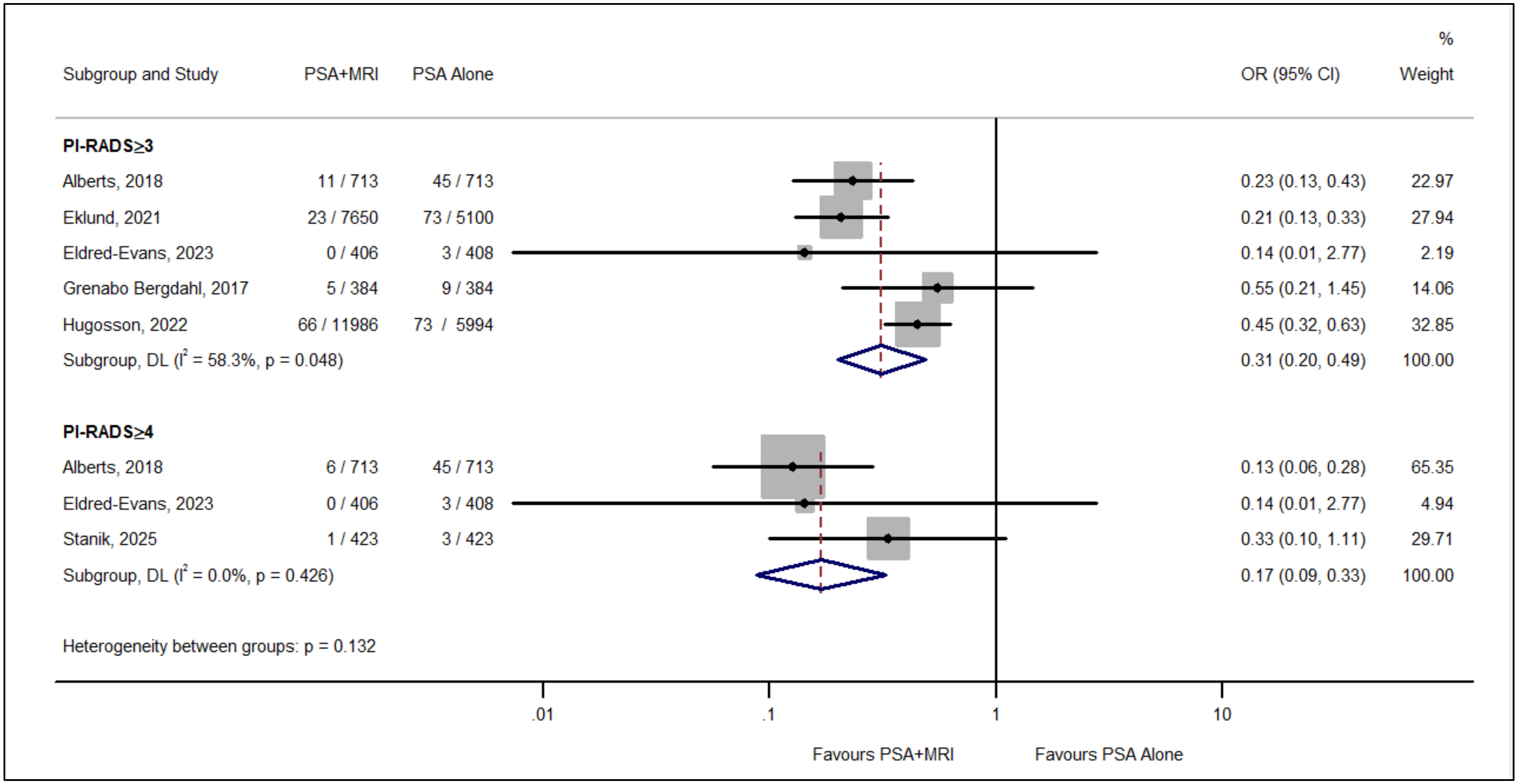


**Fig 11.** Clinically insignificant prostate cancer, comparison between MRI followed by targeted biopsy after PSA-based screening versus PSA-based screening alone, subgroup analysis based on PI-RADS thresholds (PP)

**Screening for prostate cancer using PSA with and without MRI: systematic reviews with meta-analysis**
Jennifer Pillay, Lindsay Gaudet, Sholeh Rahman, Roland Grad, Guylène Thériault, Philipp Dahm, Keith J Todd, Gail McCartney, Brett Thombs, Sabrina Saba, Lisa Hartling

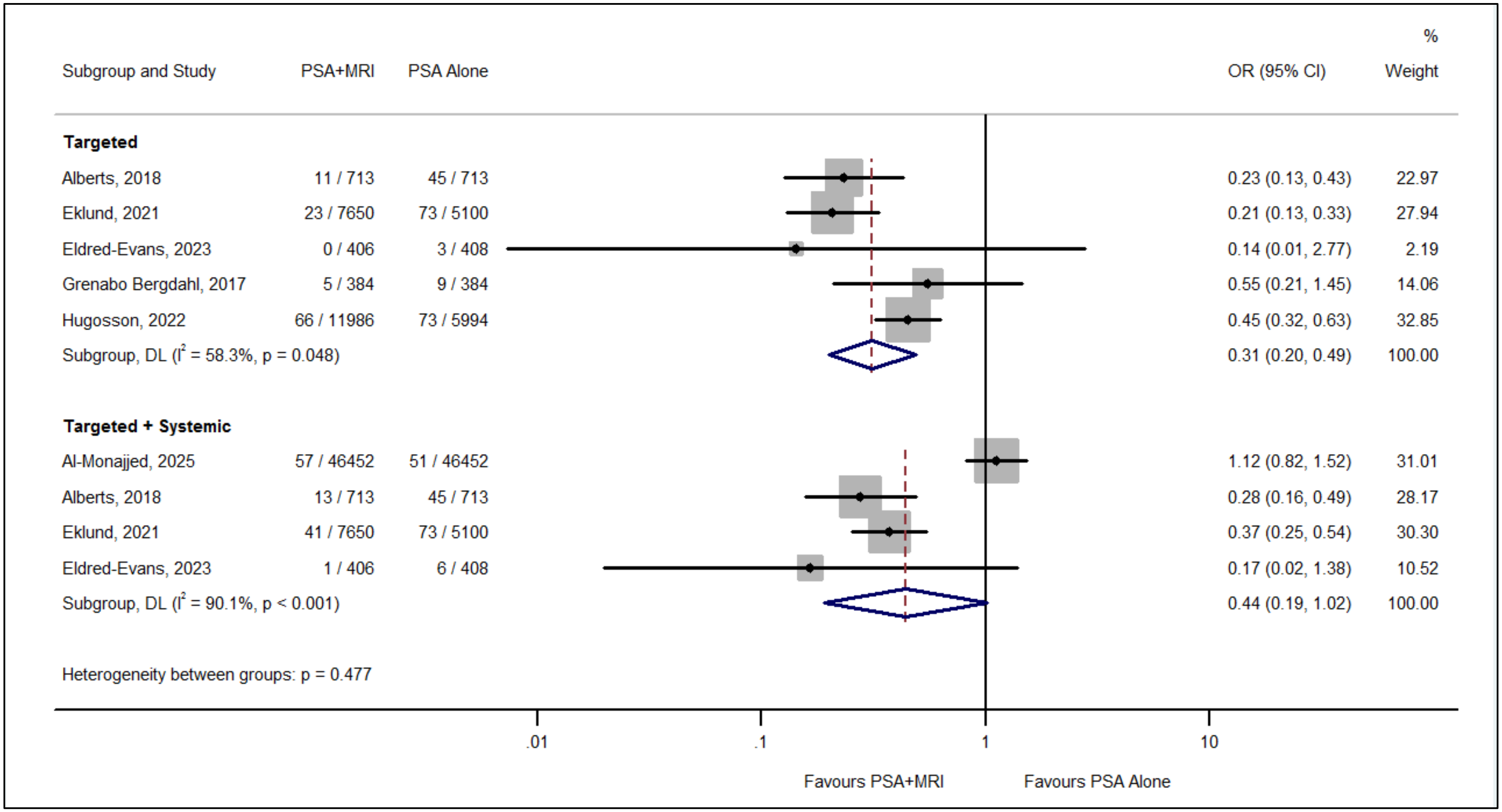


**Fig 12.** Clinically insignificant prostate cancer, subgroup analysis for PI-RADS≥3 MRI cutoff based on comprehensiveness of biopsy in the MRI arm (PP)

**Screening for prostate cancer using PSA with and without MRI: systematic reviews with meta-analysis**
Jennifer Pillay, Lindsay Gaudet, Sholeh Rahman, Roland Grad, Guylène Thériault, Philipp Dahm, Keith J Todd, Gail McCartney, Brett Thombs, Sabrina Saba, Lisa Hartling

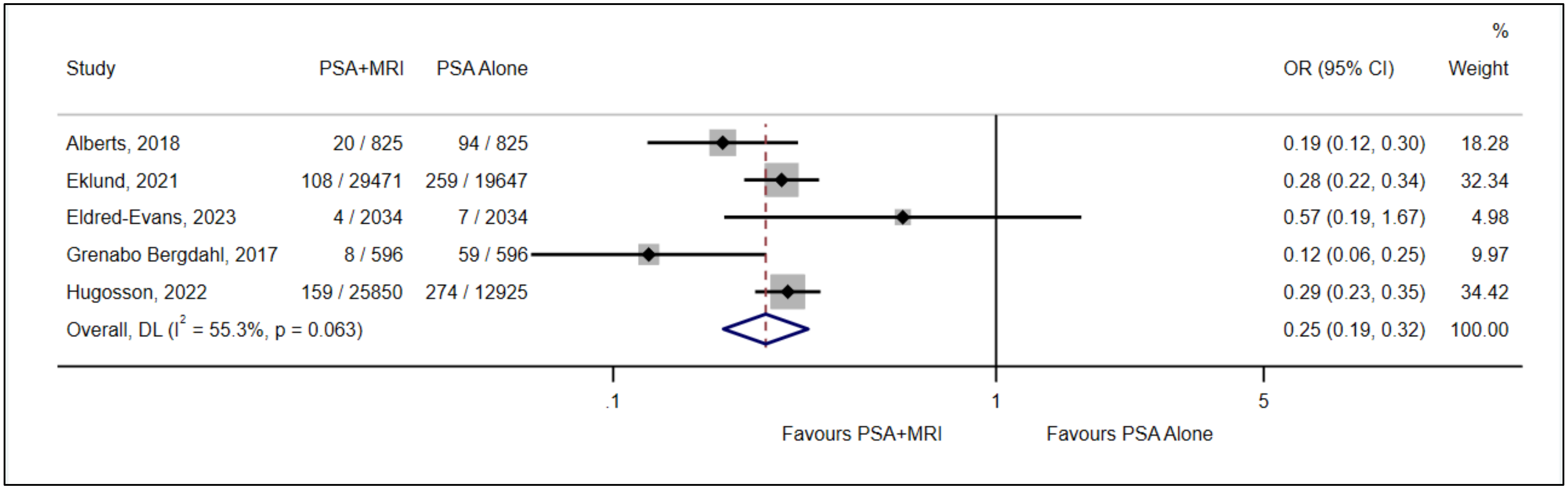


**Fig 13.** False positives, comparison between MRI followed by targeted biopsy after PSA-based screening versus PSA-based screening alone, based on PI-RADS≥3 MRI cutoff for cancer detection (ITT)

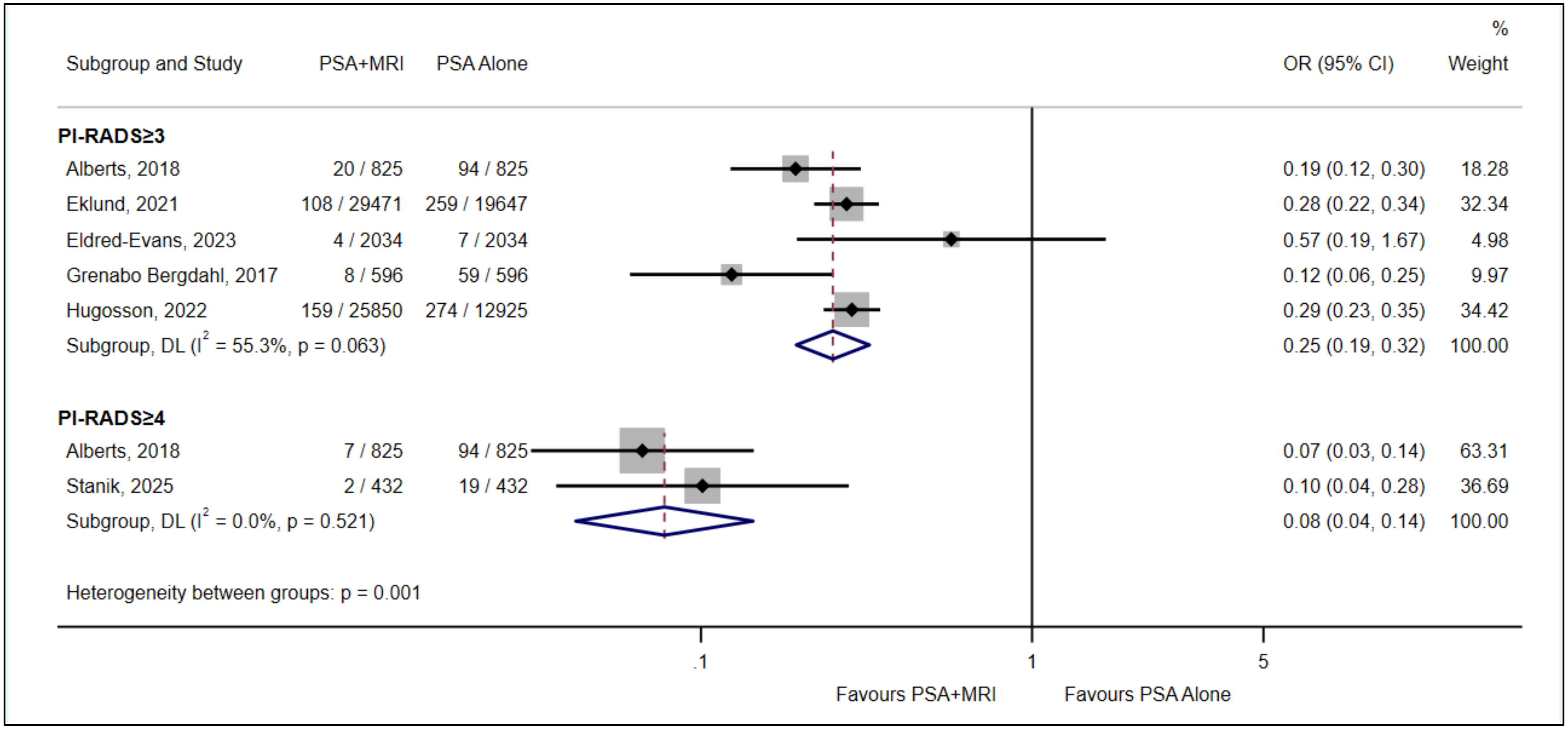


**Fig 14.** False positives, comparison between MRI followed by targeted biopsy after PSA-based screening versus PSA-based screening alone, subgroup analysis based on PI-RADS thresholds (ITT)

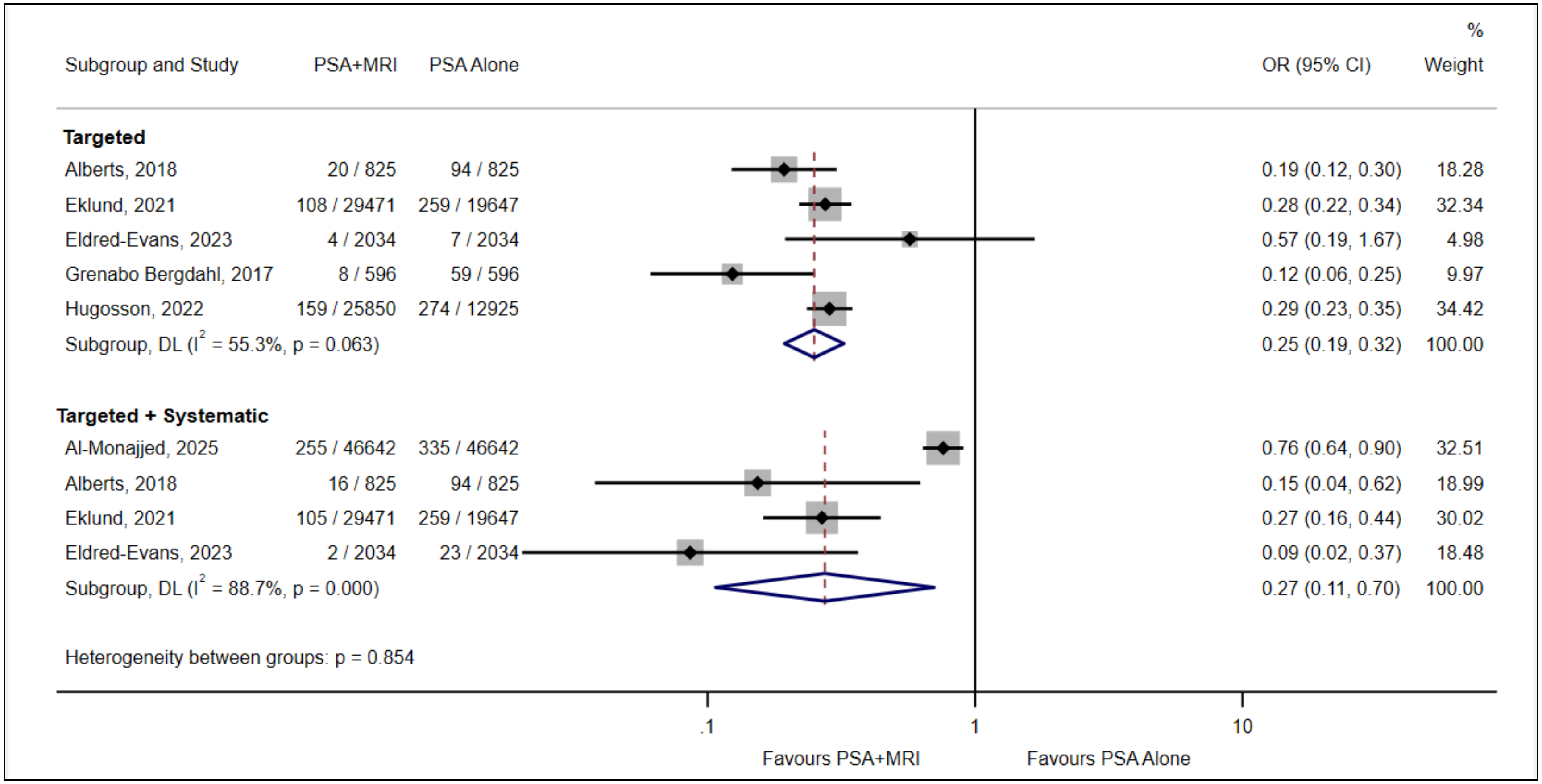


**Fig 15.** False positives, subgroup analysis for PI-RADS≥3 MRI cutoff based on comprehensiveness of biopsy in the MRI arm (ITT)

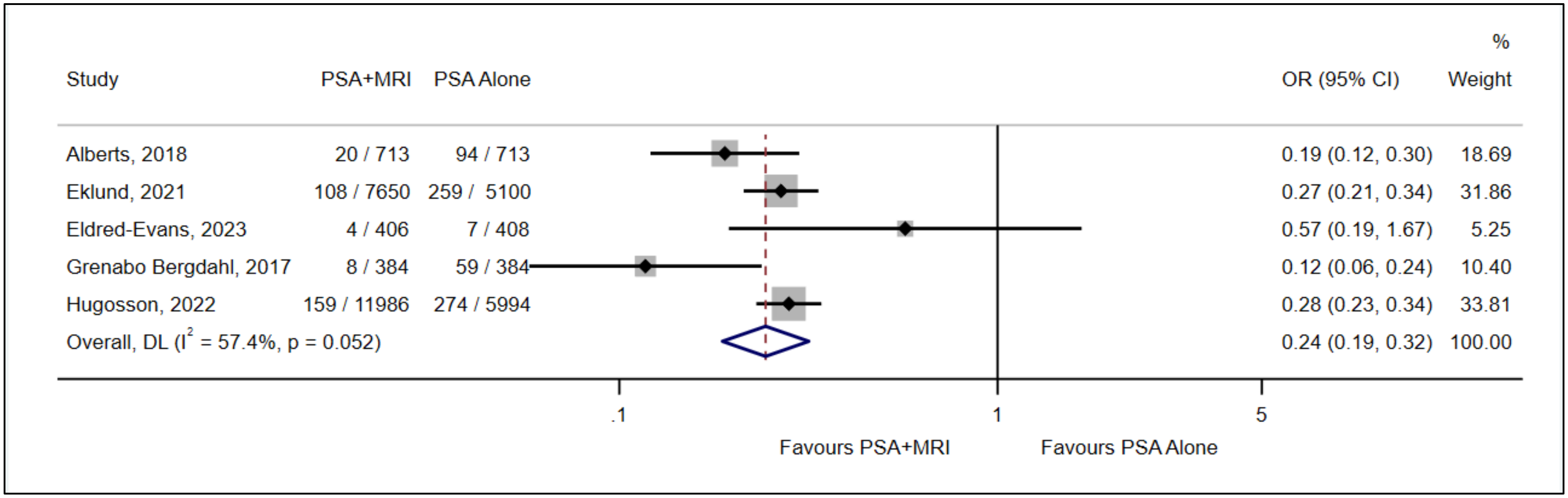


**Fig 16.** False positives, comparison between MRI followed by targeted biopsy after PSA-based screening versus PSA-based screening alone, based on PI-RADS≥3 MRI cutoff for cancer detection (PP)

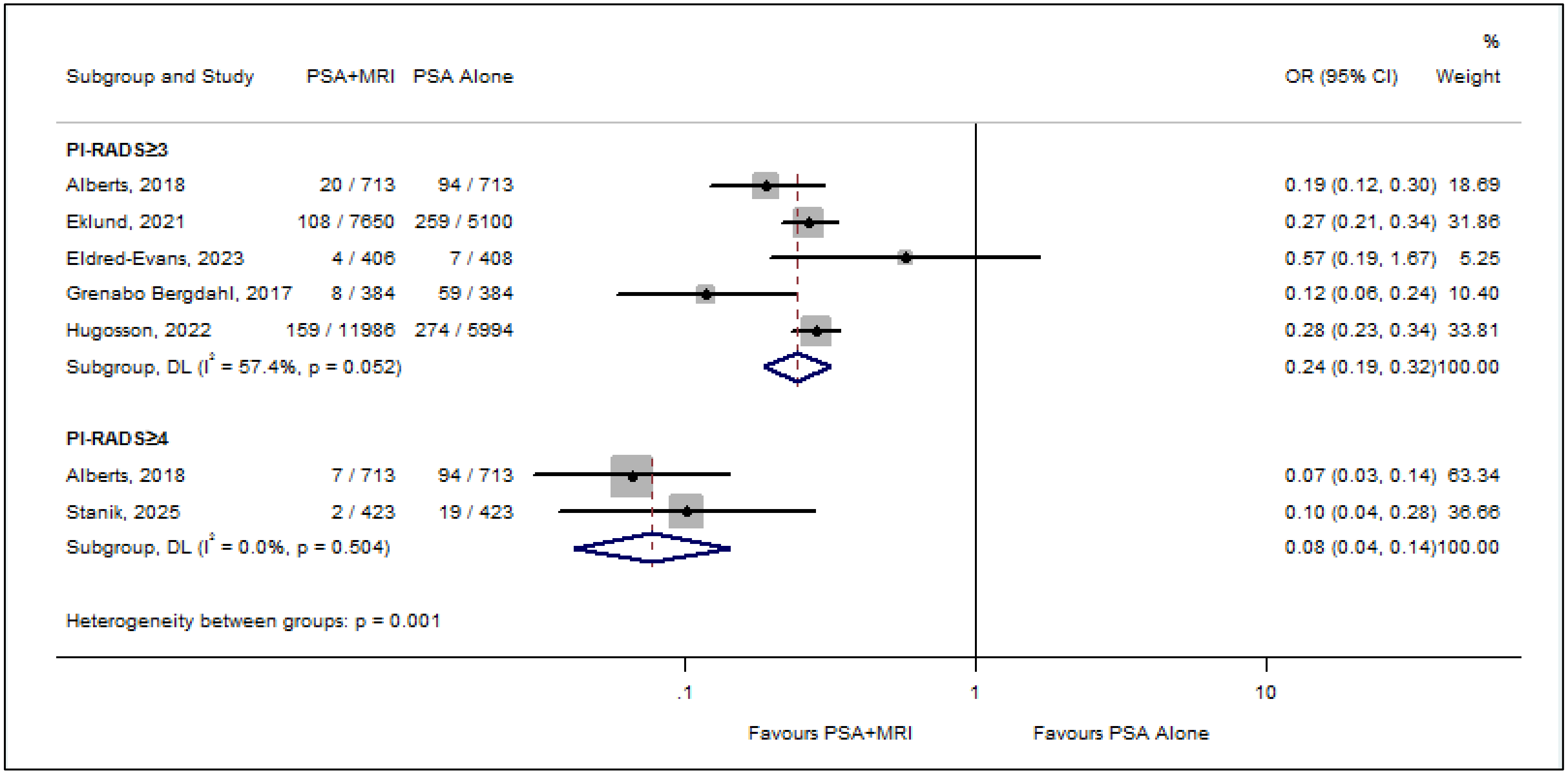


**Fig 17.** False positives, comparison between MRI followed by targeted biopsy after PSA-based screening versus PSA-based screening alone, subgroup analysis based on PI-RADS thresholds (PP)

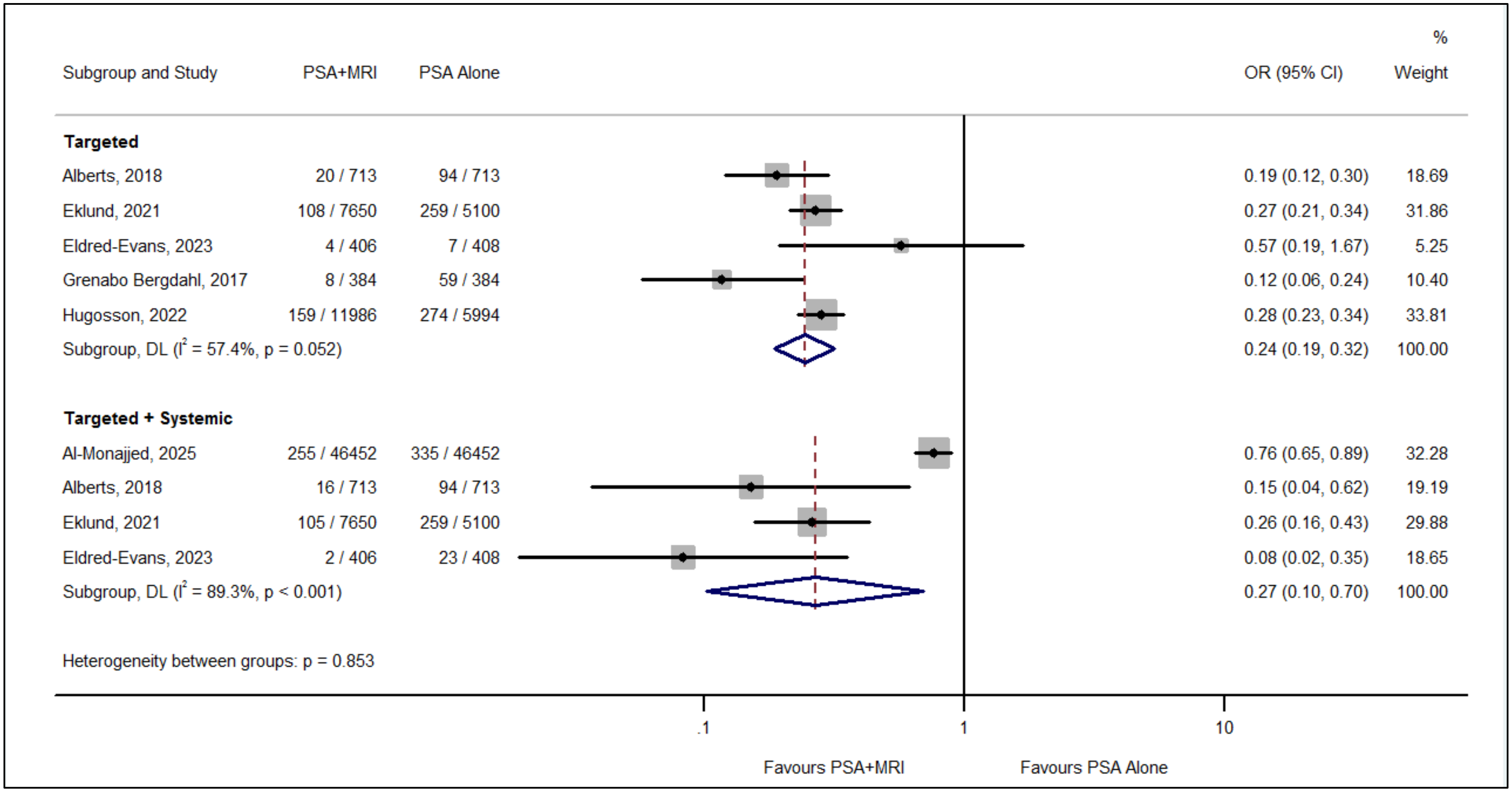


**Fig 18.** False positives, subgroup analysis for PI-RADS≥3 MRI cutoff based on comprehensiveness of biopsy in the MRI arm (PP)

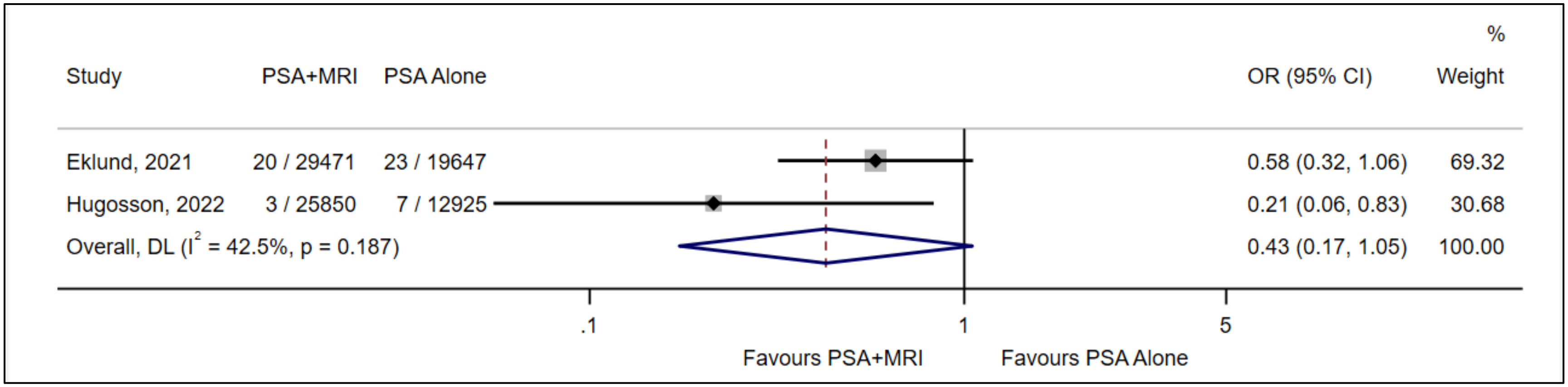


**Fig 19.** Complications (infections), comparison between MRI followed by targeted biopsy after PSA-based screening versus PSA-based screening alone, based on PI-RADS≥3 MRI cutoff for cancer detection (ITT)

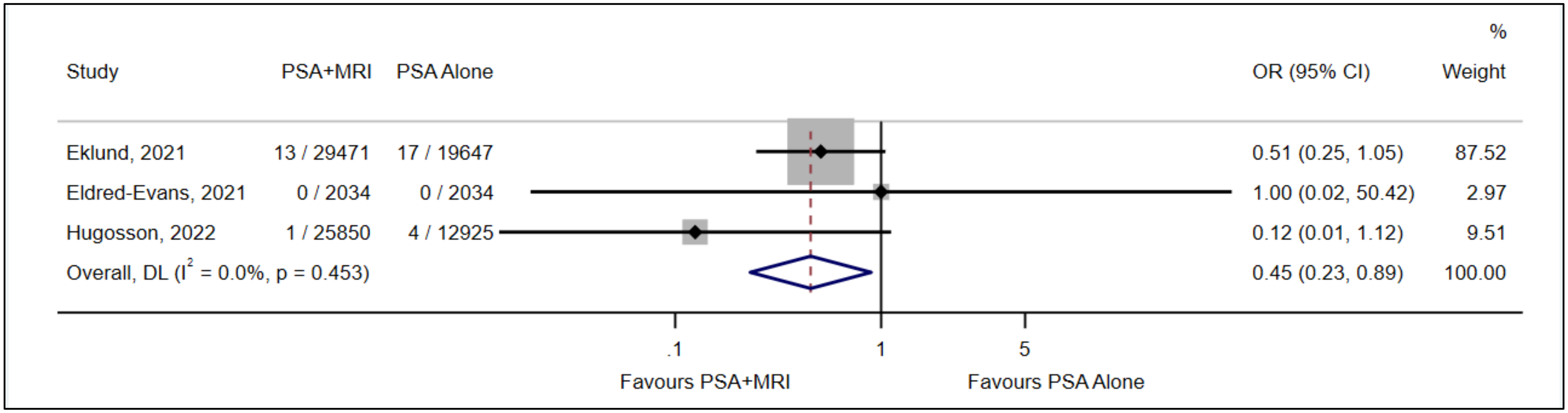


**Fig 20.** Complications (hospitalized after biopsy in 2 largest), comparison between MRI followed by targeted biopsy after PSA-based screening versus PSA-based screening alone, based on PI-RADS≥3 MRI cutoff for cancer detection (ITT)

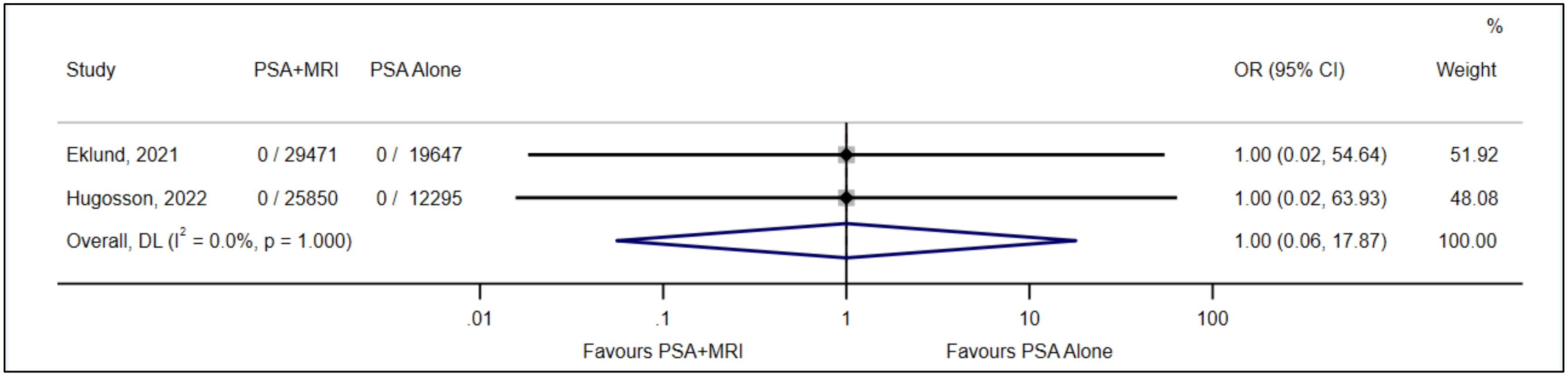


**Fig 21.** Complications (mortality following biopsy), comparison between MRI followed by targeted and systematic biopsy after PSA-based screening versus PSA-based screening alone, based on PI-RADS≥3 MRI cutoff for cancer detection (ITT)

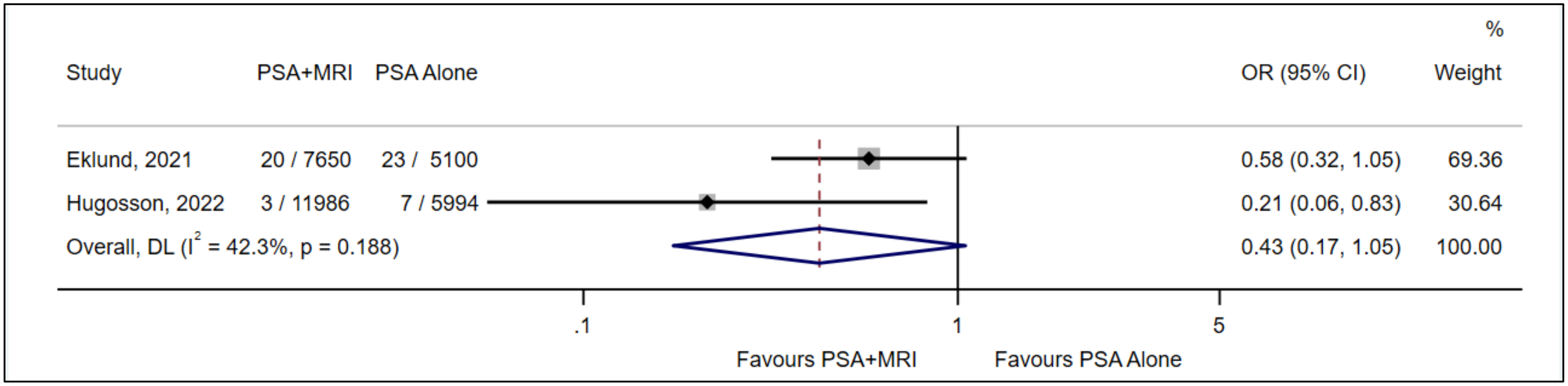


**Fig 22.** Complications (infections), comparison between MRI followed by targeted and systematic biopsy after PSA-based screening versus PSA-based screening alone, based on PI-RADS≥3 MRI cutoff for cancer detection (PP)

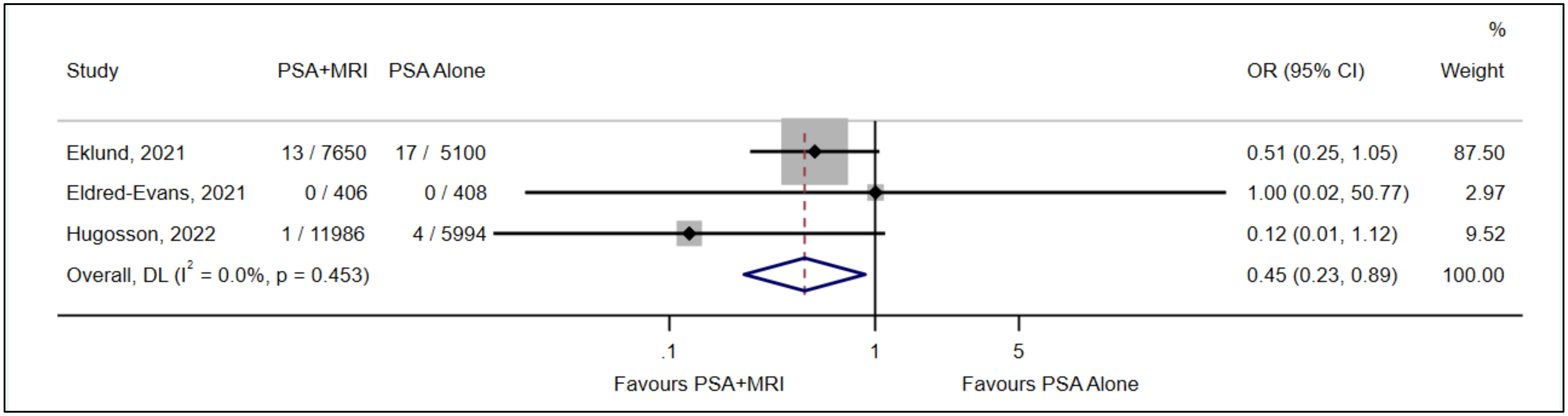


**Fig 23.** Complications (hospitalization), comparison between MRI followed by targeted biopsy after PSA-based screening versus PSA-based screening alone, based on PI-RADS≥3 MRI cutoff for cancer detection (PP)

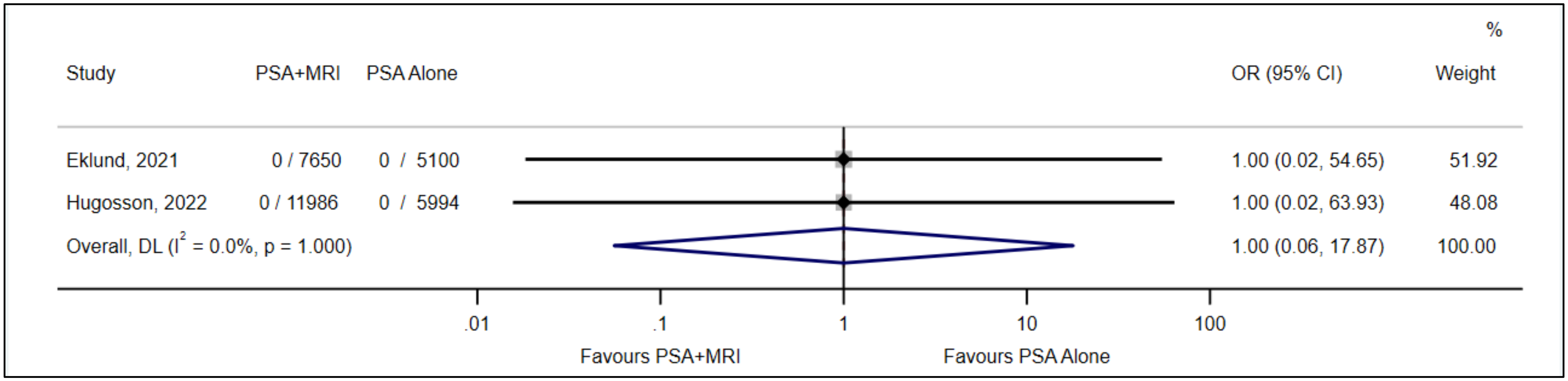


**Fig 24.** Complications (mortality following biopsy), comparison between MRI followed by targeted and systematic biopsy after PSA-based screening versus PSA-based screening alone, based on PI-RADS≥3 MRI cutoff for cancer detection (PP)

**Screening for prostate cancer using PSA with and without MRI: systematic reviews with meta-analysis**
Jennifer Pillay, Lindsay Gaudet, Sholeh Rahman, Roland Grad, Guylène Thériault, Philipp Dahm, Keith J Todd, Gail McCartney, Brett Thombs, Sabrina Saba, Lisa Hartling

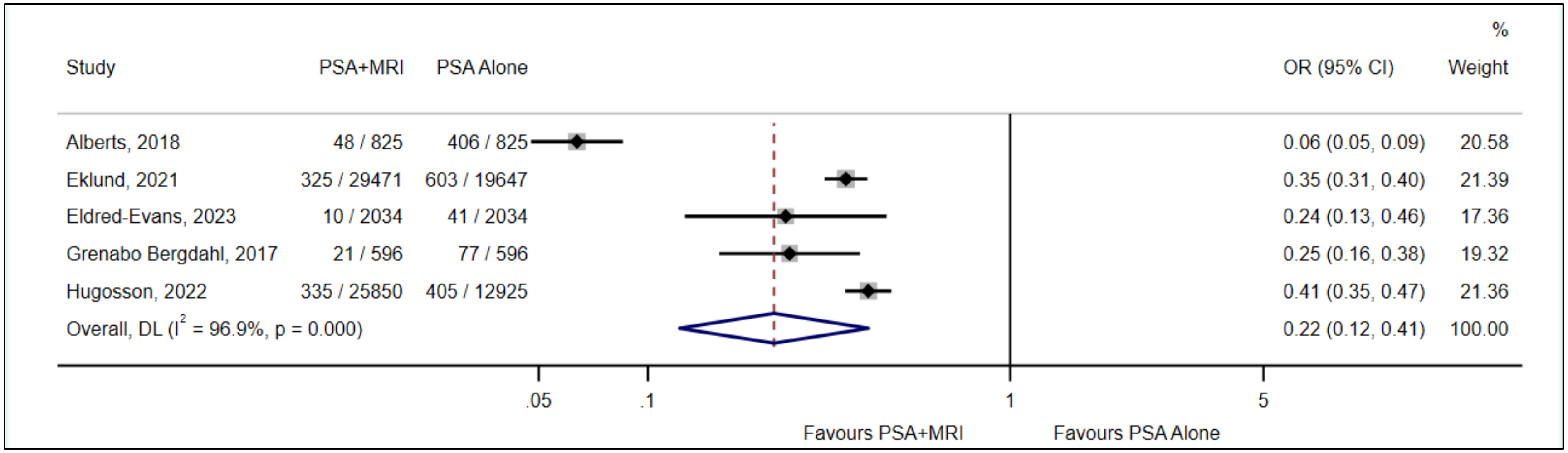


**Fig 25.** Biopsy referral, comparison between MRI followed by targeted biopsy after PSA-based screening versus PSA-based screening alone, based on PI-RADS≥3 MRI cutoff for cancer detection (ITT)

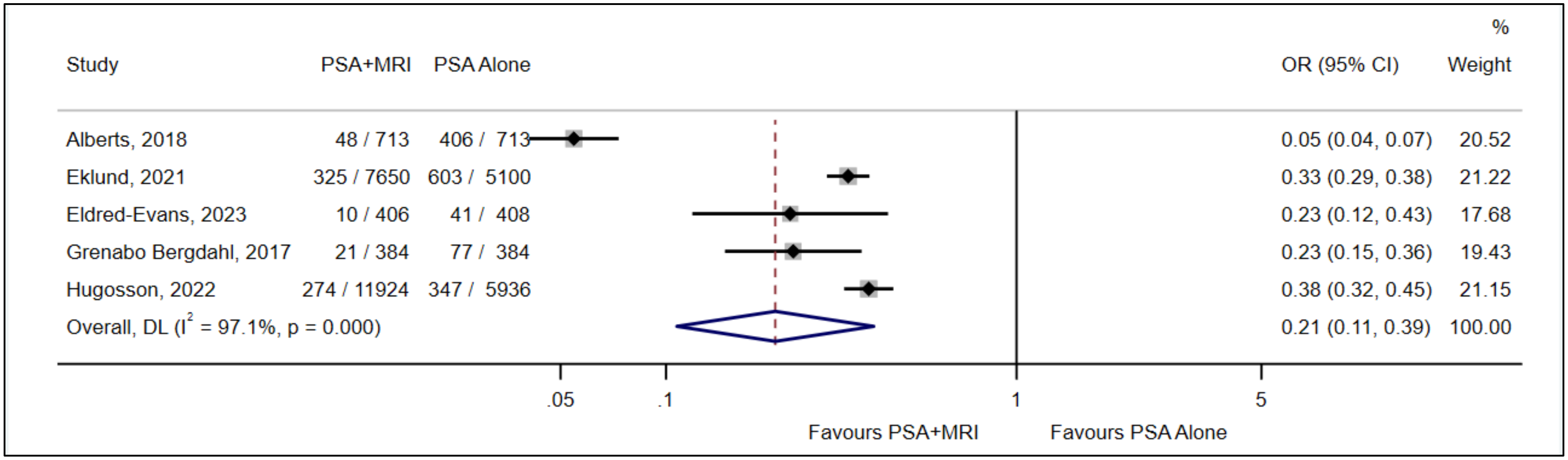


**Fig 26.** Biopsy referral, comparison between MRI followed by targeted biopsy after PSA-based screening versus PSA-based screening alone, based on PI-RADS≥3 MRI cutoff

**Screening for prostate cancer using PSA with and without MRI: systematic reviews with meta-analysis**

Jennifer Pillay, Lindsay Gaudet, Sholeh Rahman, Roland Grad, Guylène Thériault, Philipp Dahm, Keith J Todd, Gail McCartney, Brett Thombs, Sabrina Saba, Lisa Hartling

#### Appendix 4. Methods to estimate burden of management harms among overdiagnosed cases of low-risk prostate cancer

#### DRAFT

Methods

In order to estimate harms attributable to medical management of over-diagnosed cases (due to a screening program; i.e., "overtreatment"), we searched for reports of harms due to common management strategies in patients with low-risk prostate cancers. We focused on overdiagnosed cases as these men are more likely to not benefit from being diagnosed and thus bear the largest burden of harm. Management strategies of interest included: active surveillance (AS), radical prostatectomy (RP), and radiation therapy (RT; external beam radiation therapy or brachytherapy). AS, RP, and RT account for initial management of the vast majority of localized prostate cancers.[1] We did not include other therapies (e.g., hormone therapy, focal ultrasound, etc.) in our assessment of management harms because they are infrequently utilized for low-risk prostate cancers. Outcomes (harms) of interest were erectile dysfunction (ED), urinary incontinence (UI), and bowel incontinence (BI). We focused on physical harms attributable to medical management and not psychological harms that may also be the result of receiving a cancer diagnosis.

One author (LG) identified relevant publications from a range of sources including screening trials identified as part of the systematic review, studies included in relevant systematic reviews and guidelines, and records from a database search previously developed to identify comparative reports of prostate cancer management strategies. Reports in men diagnosed with low-risk prostate cancer of management choice (in those diagnosed since 2015) or patient-reported harms over a minimum of five years of follow-up were considered.

Estimation took a 3-stage approach. First, we gathered information to estimate the proportion of men with low-risk prostate cancer who underwent the management strategies of interest (AS, RP, RT); these were choices at diagnosis, recognizing that men may change management approaches (e.g., many will switch at some point from AS to surgery or radiation). We looked for large (n ≥ ~500 per treatment) trials or prospective cohort studies reporting on initial management strategies undergone by men diagnosed with low-risk prostate cancer, determined using any available risk stratification tool (e.g., D'Amico score, NCCN guidelines). Because management patterns have changed substantially in the last 20 years (i.e., use of AS has doubled in the US since 2014)[2], we relied on data from studies reporting management choices by men diagnosed since ~2015.

Second, we estimated the proportion of men choosing each management strategy who experienced a harm of interest during follow-up, regardless of what treatment they received over time. Under the assumption that the rate of harms (e.g., ED, UI, FI) have not substantially changed, there were no limits on year of diagnosis for data on the patient reported harms. In order to examine how the rate of harms may change with age, we focused on reports of our chosen harms over at least five years after diagnosis.

Finally, we applied the proportions of men receiving each management strategy and the men undergoing each management strategy and experiencing the harms of interest to the number of overdiagnosed cases estimated in KQ1 (i.e., 24 overdiagnosed cases per 1000 men screened), assuming that nearly all (i.e., >95) overdiagnosed cases are low (or very low) risk cancers (see Table 2 in text). For each time point after diagnoses, we subtracted the baseline rate, as the occurrence of our harms of interest at baseline would not be attributable to medical management.

Management choice data

Proportion estimates of management choice were based on reports from four prospective cohorts in three countries. Three publications of the ERSPC trial reported on patient choice among men diagnosed with low-risk prostate cancer, but ultimately did not inform our management strategy estimates because nearly all of the diagnoses occurred prior to 2015.[3-5]

**Table A. Proportion of men diagnosed with low-risk prostate cancer undergoing common management strategies**

| Study<br>Cohort | Country<br>Years of diagnosis | Active surveillance | Radical prostatectomy | Radiation therapy |
|---|---|---|---|---|
| Botejue 2019[6]<br>PURC | US<br>2015-2017 | 57% | 37% | 6% |
| Tiruye 2022[7]<br>SA-PCCOC registry | Australia<br>2010-2019 | 51% | 37% | 12% |
| Xu 2022[1]<br>TOPCS | US<br>2014-2017 | 57% | 23% | 17% |
| Stroomberg 2024[8]<br>DanProst | Denmark<br>2016-2021 | 63% | 14% | 2% |
| Mean | | 57% | 28% | 9% |
| Median | | 57% | 30% | 9% |
| Number used in analyses: | | 60% | 30% | 10% |

*Note: some studies reported small proportions of patients receiving other treatments not included in this evaluation so proportions within study may not add to 100%.*

Management harms data

For our assessment of management harms, estimates were informed by long-term findings of the ProTECT trial,[9] which randomized men diagnosed with localized cancer to AS, RP or RT. In ProTECT, patient reported-outcomes were measured using the EPIC questionnaire at baseline/before curative treatment, at 6 months, and yearly from 1-12 years after diagnosis (only data for baseline, 1 yr, 5 yrs, and 10 yrs are used here). When the proportion during follow-up was lower than baseline, we defaulted to using the baseline proportion as we assume this was due to variance in measurement between timepoints (e.g., cases reporting the harm at baseline not completing a follow-up survey) and not due to spontaneous recovery of function. Although the US SEER database also reported management harms after 10 years of follow-up, we were unable to incorporate this data into our estimates because the SEER database does not adequately distinguish between AS (observation with intent to switch to curative treatment as needed) and watchful waiting (observation with intent to switch to palliative treatment).[10]

**Table B. Frequency of erectile dysfunction, urinary incontinence, and bowel incontinence over time by prostate cancer management strategy**

| | Active surveillance, all | Active surveillance, no curative treatment | Radical prostatectomy | Radiation therapy |
|---|---|---|---|---|
| *Erectile dysfunction*[1] | | | | |
| Baseline | 32% | 38% | 33% | 31% |
| 1 yr | 44% | 50% | 84% | 59% |
| 5 yrs | 61% | 58% | 75% | 70% |
| 10 yrs | 76% | 72% | 82% | 82% |
| *Urinary incontinence*[2] | | | | |
| Baseline | 0% | 0% | 1% | 0% |
| 1 yr | 4% | 1% | 24% | 3% |
| 5 yrs | 5% | 2% | 14% | 2% |
| 10 yrs | 7% | 4% | 20% | 5% |
| *Bowel incontinence*[3] | | | | |
| Baseline | 4% | NR | 3% | 1% |
| 1 yr | 1%* | NR | 2%* | 12% |
| 5 yrs | 4% | NR | 2%* | 6% |
| 10 yrs | 4% | NR | 3% | 10% |

Footnotes: [1]Erections not firm enough for intercourse; [2]One or more pads per day in past 4 weeks; [3]Fecal leakage once per week or more often. *proportion replaced with baseline measure in burden calculations.

<u>Burden of management harms attributed to prostate-cancer screening</u>

The estimated burdens of management harms from treatment of overdiagnosed low-risk prostate cancers are presented in Table C.

**Table C. Burden of treatment harms among overdiagnosed cases, reported as number of per 1000 men screened (bold text indicates numbers used in Figure 4 of text)**

| | Active surveillance, all | Active surveillance, no curative treatment | Radical prostatectomy | Radiation therapy | Attributable to overtreatment (i.e., minus baseline rate) |
|---|---|---|---|---|---|
| | *Of 1000 men screened, 24 will be overdiagnosed with low-risk prostate cancer, 14.4 (60%) of whom will choose AS for primary management.*<br><br>*Of the 14.4 cases choosing AS, 6.5 (45%) will not undergo curative/radical treatment within 10 yrs.*[11] | | *Of 1000 men screened, 24 will be overdiagnosed with low-risk prostate cancer, 7.2 (30%) of whom will choose RP for primary management.* | *Of 1000 men screened, 24 will be overdiagnosed with low-risk prostate cancer, 2.4 (10%) of whom will choose RT for primary management.* | *NA* |
| *Erectile dysfunction*[1] | | | | | |
| Baseline | *4.6 of the 14.4 overdiagnosed cases managed with AS will report ED before treatment.* | | *2.4 of the 7.2 overdiagnosed cases treated with RP will report ED before treatment.* | *0.7 of the 2.4 overdiagnosed cases treated with RT will report ED before treatment.* | *Baseline rate: 7.7 men per 1000* |

| | | | | | |
|---|---|---|---|---|---|
| 1 yr | 6.3 men will report ED 1 yr after dx. | 3.2 men will report ED 1 yr after dx | 6.0 men will report ED 1 yr after dx. | 1.4 man will report ED 1 yr after dx. | 6.0 per 1000 |
| 5 yrs | 8.8 men will report ED 5 yrs after dx. | 3.8 men will report ED 5 yrs after dx | 5.4 men will report ED 5 yrs after dx. | 1.7 man will report ED 5 yrs after dx. | 8.2 per 1000 |
| 10 yrs | 10.9 men will report ED 10 yrs after dx. | 4.7 men will report ED 10 yrs after dx | 5.9 men will report ED 5 yrs after dx. | 2.0 men will report ED 10 yrs after dx. | **11.1 per 1000** |
| *Urinary incontinence[2]* | | | | | |
| Baseline | *0 of the 14.4 overdiagnosed cases managed with AS will report UI before treatment.* | | *0.1 of the 7.2 overdiagnosed cases treated with RP will report UI before treatment.* | *0 of the 2.4 overdiagnosed cases treated with RT will report UI before treatment.* | *Baseline rate: 0.1 per 1000* |
| 1 yr | 0.6 men will report UI 1 yr after dx. | 0.1 men will report UI 1 yr after dx. | 1.7 man will report UI 1 yr after dx. | 0.1 men will report UI 1 yr after dx. | 2.3 per 1000 |
| 5 yrs | 0.7 men will report UI 5 yrs after dx. | 0.1 men will report UI 5 yrs after dx. | 1.0 man will report UI 5 yrs after dx. | 0.1 men will report UI 5 yrs after dx. | 1.7 per 1000 |
| 10 yrs | 1.0 men will report UI 10 yrs after dx. | 0.3 men will report UI 10 yrs after dx. | 1.4 man will report UI 10 yrs after dx. | 0.1 men will report UI 10 yrs after dx. | **2.4 per 1000** |
| *Fecal incontinence[3]* | | | | | |
| Baseline | *0.6 of the 14.4 overdiagnosed cases managed with AS will report FI before treatment.* | | *0.2 of the 7.2 overdiagnosed cases treated with RP will report UI before treatment.* | *0.02 of the 2.4 overdiagnosed cases treated with RT will report UI before treatment.* | *Baseline rate: 0.8 per 1000* |
| 1 yr | 0.6* men will report FI 1 yr after dx. | No evidence available | 0.2* men will report FI 1 yr after dx. | 0.3 men will report FI 1 yr after dx. | 0.3 per 1000 |
| 5 yrs | 0.6 man will report FI 5 yrs after dx. | | 0.2* men will report FI 5 yrs after dx. | 0.1 men will report FI 5 yrs after dx. | 0.1 per 1000 |
| 10 yrs | 0.6 man will report FI 10 yrs after dx. | | 0.2 men will report FI 10 yrs after dx. | 0.2 men will report FI 10 yrs after dx. | **0.2 per 1000** |

Footnotes: [1]Erections not firm enough for intercourse; [2]One or more pads per day in past 4 weeks; [3]Fecal leakage once per week or more often. *baseline proportion used in burden calculations.
